# Gaps Between Willingness and Uptake of Influenza and COVID-19 Vaccines During the 2025-26 Respiratory Virus Season in a U.S. Adult Cohort

**DOI:** 10.64898/2026.07.17.26358235

**Authors:** Jenna Sanborn, McKaylee M Robertson, Kate Penrose, Madhura Rane, Rachael Piltch-Loeb, Angela M Parcesepe, Denis Nash

## Abstract

During the 2025-26 respiratory virus season, changes to COVID-19 vaccine eligibility, recommendations, and communication may have made vaccination follow-through especially challenging. Under-vaccination may reflect not only lack of willingness, but also breakdowns between willingness and uptake. We analyzed data from 3,390 adults in the CHASING COVID Cohort who completed assessments in August 2025 and March 2026 to examine gaps between vaccine willingness and subsequent influenza and COVID-19 vaccination. Vaccine willingness was defined using prior-season vaccination and stated intention to vaccinate during the 2025-26 respiratory virus season. In August 2025, 73% of participants were influenza vaccine willing and 68% were COVID-19 vaccine willing. Among vaccine-willing participants, 17% and 39% were unvaccinated for influenza and COVID-19, respectively, by March 2026. Absence of prior-season vaccination was the strongest predictor of not vaccinating for influenza and COVID-19, respectively (aRR [95% CI]: 4.04 [3.44-4.74]; 3.01 [2.72-3.34]). Non-vaccination was also associated with food insecurity (1.99 [1.66-2.37]; 1.47 [1.33-1.63]), any healthcare barrier (1.89 [1.57-2.28]; 1.51 [1.36-1.67]), and being not at all confident in vaccine safety (2.95 [2.15-4.05]; 2.21 [1.88-2.59]). Trajectory analyses suggested willingness-uptake gaps reflected incomplete follow-through on intentions and discontinuation among some prior vaccinators. Commonly reported reasons among vaccine-willing non-vaccinators included difficulty finding a convenient time, place, or appointment and, for COVID-19, lack of healthcare provider recommendation. Findings among non-vaccinated adults with prior or stated openness to vaccinate highlight missed opportunities and suggest avenues to improve coverage through strategies that reinforce vaccine confidence, reduce access and logistical barriers, and make vaccination easier to complete.

## Introduction

Adequate vaccination coverage against SARS-CoV-2 and influenza remains important for reducing the risk of infection, emergency medical visits, and hospitalization.^1^ However, uptake of both COVID-19 and influenza vaccines has declined in recent years in the United States.^2–5^ During the 2025-26 season, an estimated 43.9% of adults aged 18+ received an influenza vaccine while only 16.1% received a COVID-19 vaccine.^6^ Research examining vaccine uptake has largely focused on vaccine hesitancy and willingness to vaccinate, often treating stated intentions as proxies for subsequent behavior.^7,8^ However, intending to be vaccinated does not necessarily translate into subsequent vaccine receipt, and under-vaccination may reflect not only vaccine refusal, but also breakdowns in the pathway from willingness to action.

Even among individuals who initially express willingness to vaccinate, structural barriers, competing demands, evolving risk perceptions, changing vaccine recommendations, and vaccine-related mis/disinformation may disrupt the pathway from intention to uptake.^9–14^ In addition, individuals who previously engaged in vaccination may no longer intend to vaccinate in subsequent seasons, reflecting erosion of vaccination intentions over time. Improving follow-through and maintenance of vaccination behavior among individuals who have already demonstrated vaccine acceptance may represent a particularly important and actionable target for intervention.^15^ Examining vaccination trajectories across prior vaccination history, current intentions, and subsequent uptake can help identify where breakdowns occur, including incomplete follow-through among those who intend to vaccinate and discontinuation among those with prior vaccination history. Understanding these patterns and the reasons for non-uptake may help identify missed opportunities and inform strategies aimed at preventing declines in vaccination rates.

These dynamics were particularly relevant during the 2025-2026 respiratory virus season, which occurred amid substantial and unprecedented upheaval in the U.S. public health and vaccine landscape.^16^ This period was characterized by ideological changes to vaccine recommendations and communication practices, disruptions to federal public health infrastructure and workforce capacity, and alarming policy changes affecting healthcare access and insurance coverage.^17–19^ At the same time, delayed and conflicting guidance surrounding vaccine recommendations may have undermined informed decision-making by individuals and providers, contributing to uncertainty regarding vaccine eligibility, insurance coverage, availability, accessibility, and timing.^20–26^ Such disruptions may influence not only whether individuals intend to vaccinate, but also whether individuals who historically tended to get vaccinated ultimately follow through and maintain vaccination behaviors over time.^27^

We used longitudinal data from a U.S. cohort to examine how vaccination history and stated intentions to vaccinate related to subsequent COVID-19 and influenza vaccine uptake. In this paper, we 1) characterize the extent of non-vaccination among vaccine-willing participants; 2) examine trajectories from 2022-23 vaccination history to 2024-25 vaccination uptake, August 2025 intention, and subsequent uptake during the 2025-26 respiratory virus season; 3) identify predictors of non-vaccination despite previous vaccination or stated intention; and 4) describe and compare reasons for non-uptake among vaccine-willing and non-willing participants.

## Methods

### Study population

The CHASING COVID Cohort is a national, community-based prospective study of adults aged ≥18 recruited in 2020 and followed approximately every three months through 2023, with additional follow-up assessments conducted in August 2025 and March 2026. Participants reside in all 50 U.S. states, Washington D.C., Puerto Rico, and Guam and represent a socio-demographically diverse population. Recruitment methods and follow-up procedures have been described previously,^28^ and study materials are publicly available.^29^

The August 2025 and March 2026 follow-up assessments were designed to capture vaccine attitudes, intentions, and uptake during a period marked by substantial vaccine mis/disinformation, changes to vaccine guidelines, and disruptions to the public health systems involved in vaccine delivery. The August 2025 assessment collected information on health characteristics, vaccine confidence, trusted sources of vaccine information, intentions to receive COVID-19 and influenza vaccinations during the 2025-26 respiratory virus season, and vaccination receipt during the previous 12 months. The March 2026 assessment measured influenza and COVID-19 vaccination uptake occurring between September 2025 and March 2026.

This study was approved by the Institutional Review Board at the City University of New York Graduate School for Public Health and Health Policy, and participants could voluntarily discontinue participation at any time.

### Analytic Sample

The analytic sample included participants who completed both the August 2025 and March 2026 assessments. We additionally excluded participants who reported receiving an influenza or COVID-19 vaccine in August 2025 before completing the August assessment, because their 2025-26 vaccination preceded the assessment of baseline vaccination intentions (n=25). The final analytic sample included 3,390 participants.

### Study Measures

Details on survey tools, response options, and categorizations for study measures are provided in Supplemental Table S1.

#### Sociodemographic and Health Characteristics

Sociodemographic characteristics assessed at enrollment included age, gender, race/ethnicity, and education level. Time-updated characteristics assessed in August 2025 included annual household income, employment status, food insecurity, housing insecurity, parent/guardian status for a child under age 18, disability status, pregnancy status, relationship status, underlying health conditions, and barriers to healthcare. Participants were classified as having an underlying health condition if they reported ever being diagnosed with one or more chronic conditions identified by the Centers for Disease Control and Prevention (CDC) as increasing risk for severe COVID-19.^30^ These included chronic obstructive pulmonary disease, emphysema, chronic bronchitis, angina or coronary heart disease, hypertension, history of myocardial infarction, current asthma, type 2 diabetes, kidney disease, rheumatic conditions, lupus, or HIV/AIDS. Healthcare access barriers included lack of health insurance coverage, inability to see a doctor within the past year due to cost, and not having a regular primary care provider.

#### Vaccine confidence and trusted information sources

At the August 2025 assessment, participants reported their trust in various sources of vaccine information and general confidence in vaccines. Trust in vaccine information sources, including healthcare providers, local health departments, the CDC, and pharmaceutical companies, was assessed using Likert-scale responses ranging from “none at all” to “a lot.” Participants additionally reported confidence in the safety of influenza and COVID-19 vaccines, federal agencies’ ability to ensure vaccine safety and effectiveness, and mRNA vaccines, using four response options: not at all confident, somewhat confident, very confident, and don’t know/not sure.

#### Vaccine willingness

We use the term “vaccine-willing” to refer to participants with behavioral or stated evidence of openness to vaccination before the start of the 2025-26 respiratory virus season. Vaccine willingness was defined using two indicators assessed at the August 2025 assessment: vaccination during the prior respiratory virus season and prospective intention to vaccinate during the upcoming 2025-26 season. Prior-season vaccination was defined separately influenza and COVID-19 as self-reported receipt of that vaccine during the 12 months before the August 2025 assessment, representing vaccination during the 2024-25 season. Prospective intention was defined based on participants’ reported plans in August 2025 to receive the corresponding vaccine during the 2025-26 respiratory virus season. For influenza vaccination, participants were classified as vaccine-willing if they reported prior-season influenza vaccination and/or reported plans to receive an influenza vaccine during the 2025-26 season. For COVID-19 vaccination, participants were classified as vaccine-willing if they reported prior-season COVID-19 vaccination and/or reported that they would definitely or probably receive an updated COVID-19 vaccine during the 2025-26 respiratory virus season.

#### Prior vaccination history

To characterize longer-term vaccination patterns, we examined prior vaccination history during the 2022-23 respiratory virus season. For COVID-19 vaccination, participants reported dates of prior vaccine doses across follow-up assessments administered from May 2021 to December 2023; receipt during the 2022-23 season was defined as any reported COVID-19 vaccination between September 2022 and August 2023. For influenza vaccination, receipt during the 2022-23 season was defined using reports of influenza vaccination in the past 12 months from the April 2023 assessment, supplemented with responses to the same item from the September 2023 assessment when April 2023 data were missing.

#### 2025-26 vaccine uptake

Vaccination uptake during the 2025-26 respiratory virus season was measured in March 2026 and defined as self-reported receipt of the influenza or COVID-19 vaccine between September 1, 2025, and the March 2026 assessment. For regression analyses, the outcome was non-vaccination during this period.

#### Reasons for non-vaccination

Participants who did not report receiving the corresponding vaccine since September 1, 2025 were asked to select reasons for non-vaccination. Response options included reasons related to vaccine safety concerns, perceived need for vaccination, healthcare provider recommendation, logistical barriers, and healthcare access barriers. Participants could select multiple reasons and could also provide open-ended responses, which were reviewed and grouped into thematic categories. Reasons for non-vaccination were summarized separately for vaccine-willing and non-willing participants for each vaccine.

### Statistical Analyses

Because influenza and COVID-19 vaccination may reflect different histories of routine uptake, recommendation clarity, and public attitudes, all analyses were conducted separately for each vaccine.^31,32^

We first summarized participant characteristics and vaccination-related measures overall and by vaccine-willing status using frequencies, percentages and chi-square tests. Then, to quantify gaps between vaccine willingness and uptake, we calculated the proportion of participants who did not receive the corresponding 2025-26 vaccine among those classified as vaccine willing in August 2025. To describe vaccination patterns across seasons and during the 2025-26 policy and communication context, we used Sankey diagrams to visualize trajectories from 2022-23 vaccination history, 2024-25 vaccination, August 2025 intention, and subsequent 2025-26 vaccination among participants with complete data at all four trajectory time points. As a supplementary analysis, we repeated the vaccination trajectory analyses among adults aged ≥65 years, who are at greater risk of severe respiratory illness and were subject to clearer COVID-19 vaccine recommendations during the 2025-26 season.^33,34^

To identify factors associated with breakdowns between vaccine willingness and uptake, we restricted analyses to vaccine-willing participants. We estimated the prevalence of non-vaccination among vaccine-willing participants and examined predictors of non-vaccination using Poisson regression models with robust standard errors to estimate age- and sex-adjusted risk ratios (aRRs) and 95% confidence intervals. Predictors included sociodemographic characteristics, healthcare access barriers, 2022-23 vaccination history, 2024-25 prior-season vaccination, vaccine confidence, and trusted sources of vaccine information.

Among participants who did not receive the corresponding vaccine during the 2025-26 respiratory virus season, we summarized self-reported reasons for non-vaccination overall and by vaccine-willing status in August 2025.

Analyses were conducted in R, version 4.3.3.

## Results

### Participant characteristics and vaccine willingness

At the August 2025 assessment, 2,484 of 3,390 participants (73%) were classified as influenza vaccine willing, and 2,300 (68%) were classified as COVID-19 vaccine willing (Table 1). Vaccine willingness for both vaccines was more common among older adults, men, White non-Hispanic participants, those with higher educational attainment and household income, participants with underlying health conditions, and those reporting fewer material or healthcare access barriers. Vaccine-willing participants also reported substantially greater vaccine confidence and trust in healthcare providers, local health departments, the CDC, and pharmaceutical companies for vaccine information. During the 2025-26 respiratory virus season, 2,118 participants (62%) reported receiving the influenza vaccine and 1,423 (42%) reported receiving the COVID-19 vaccine.

**Table 1.**
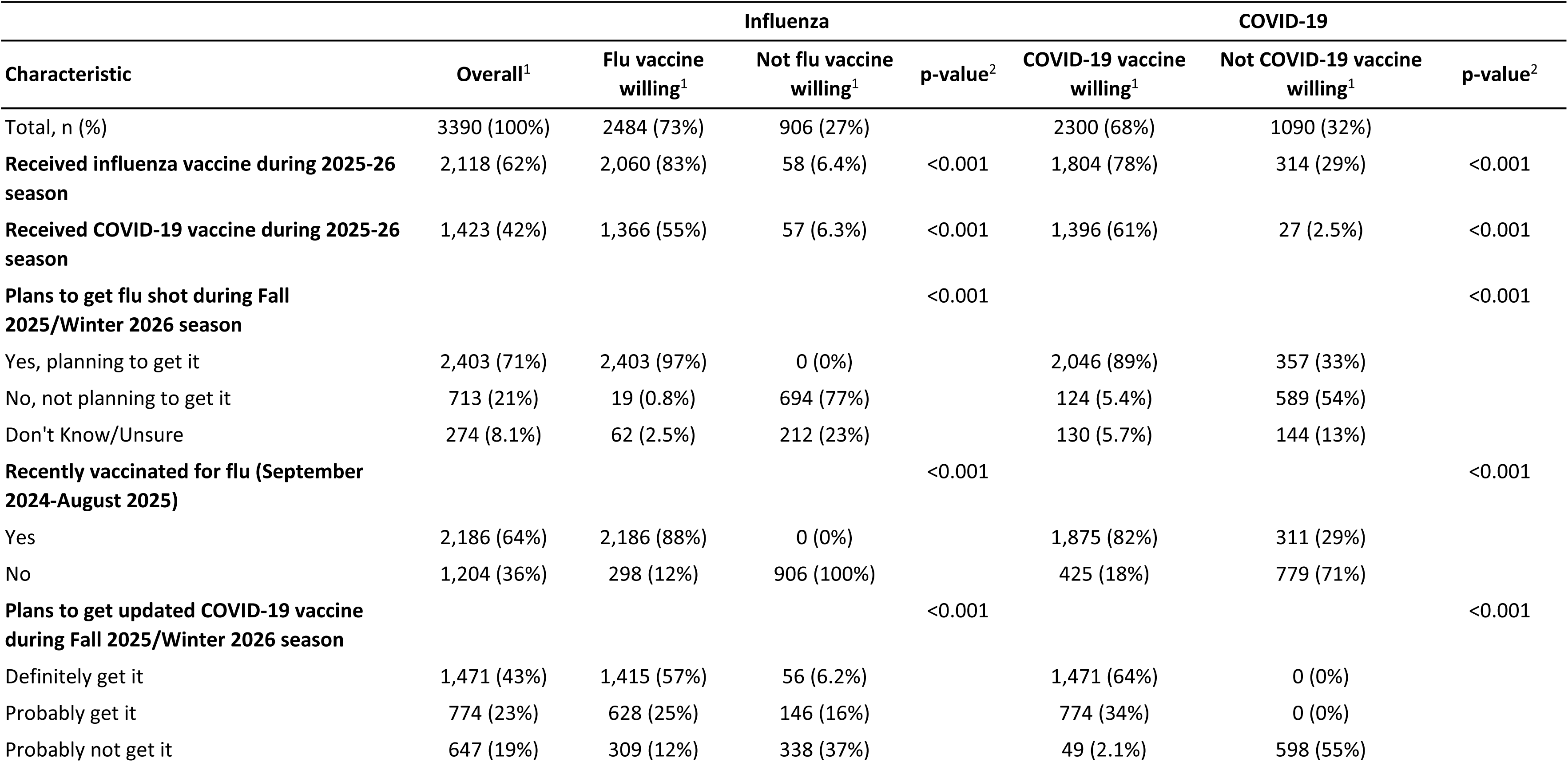
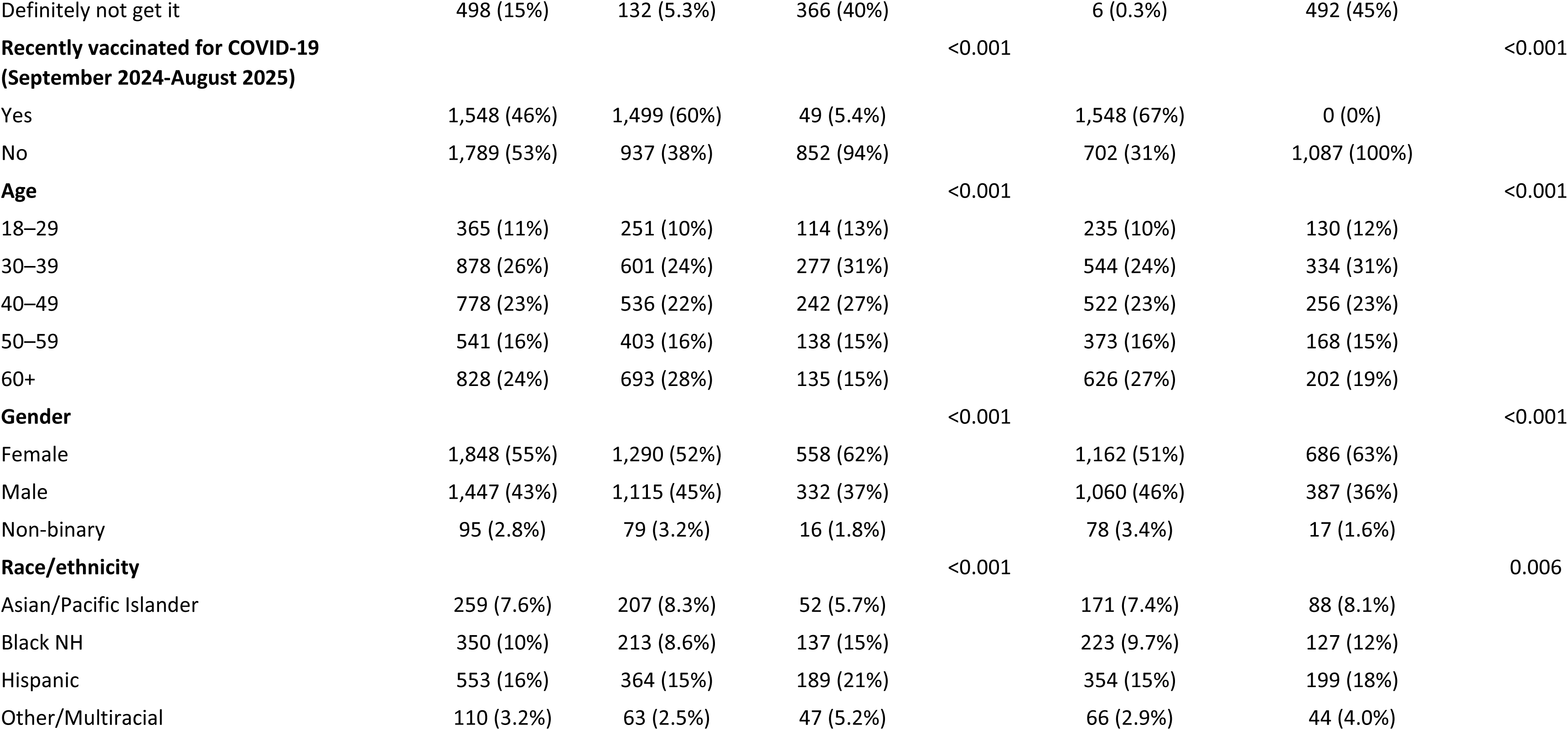
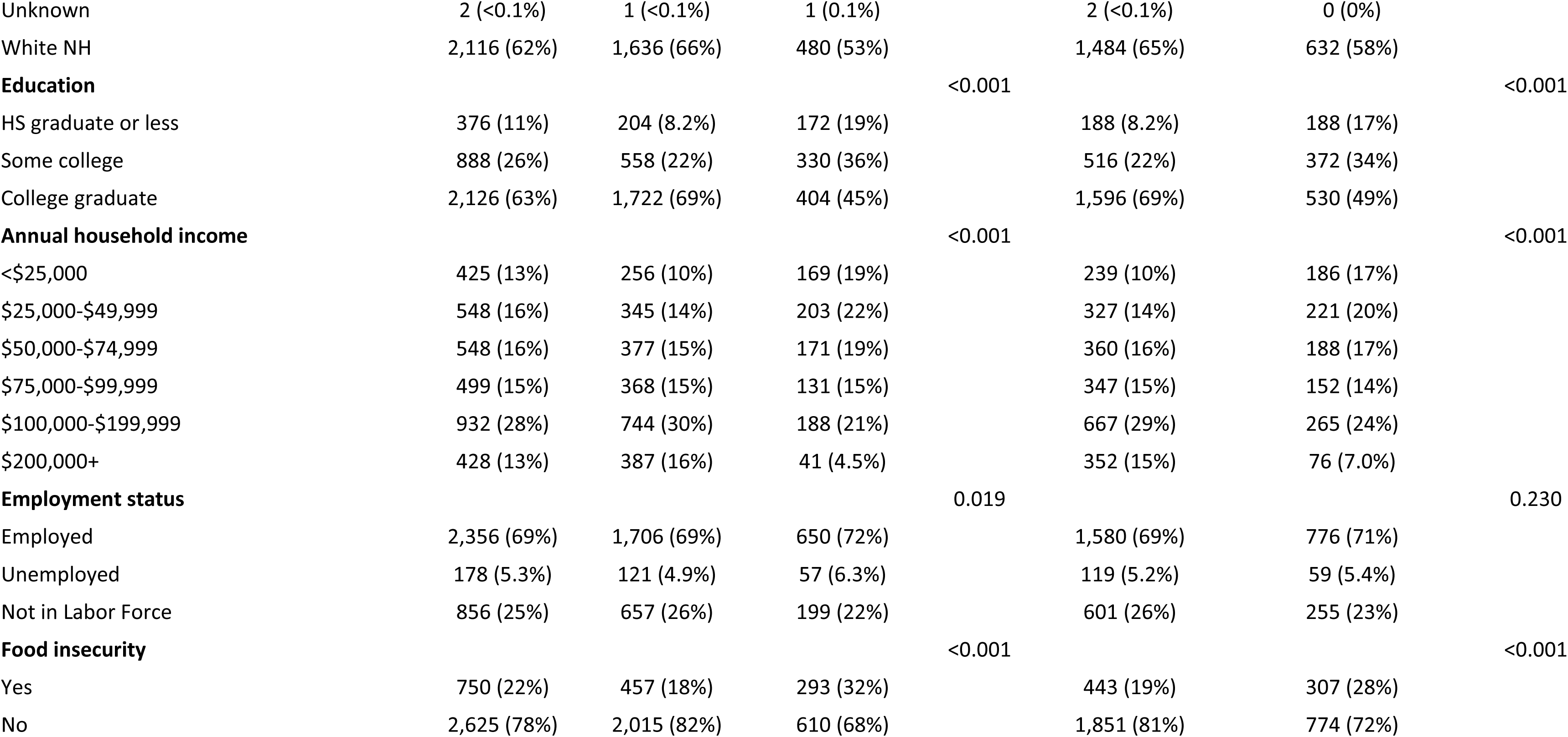
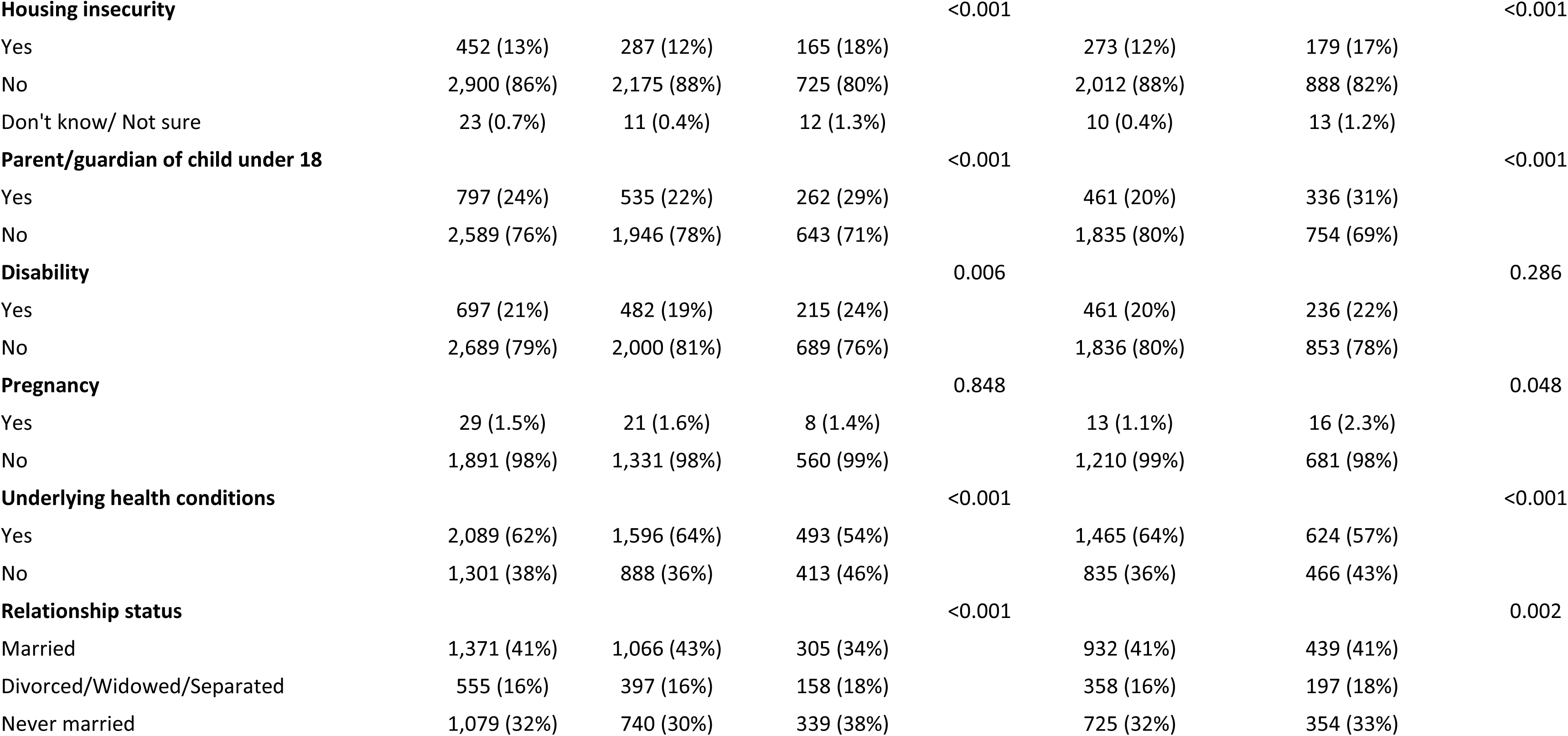
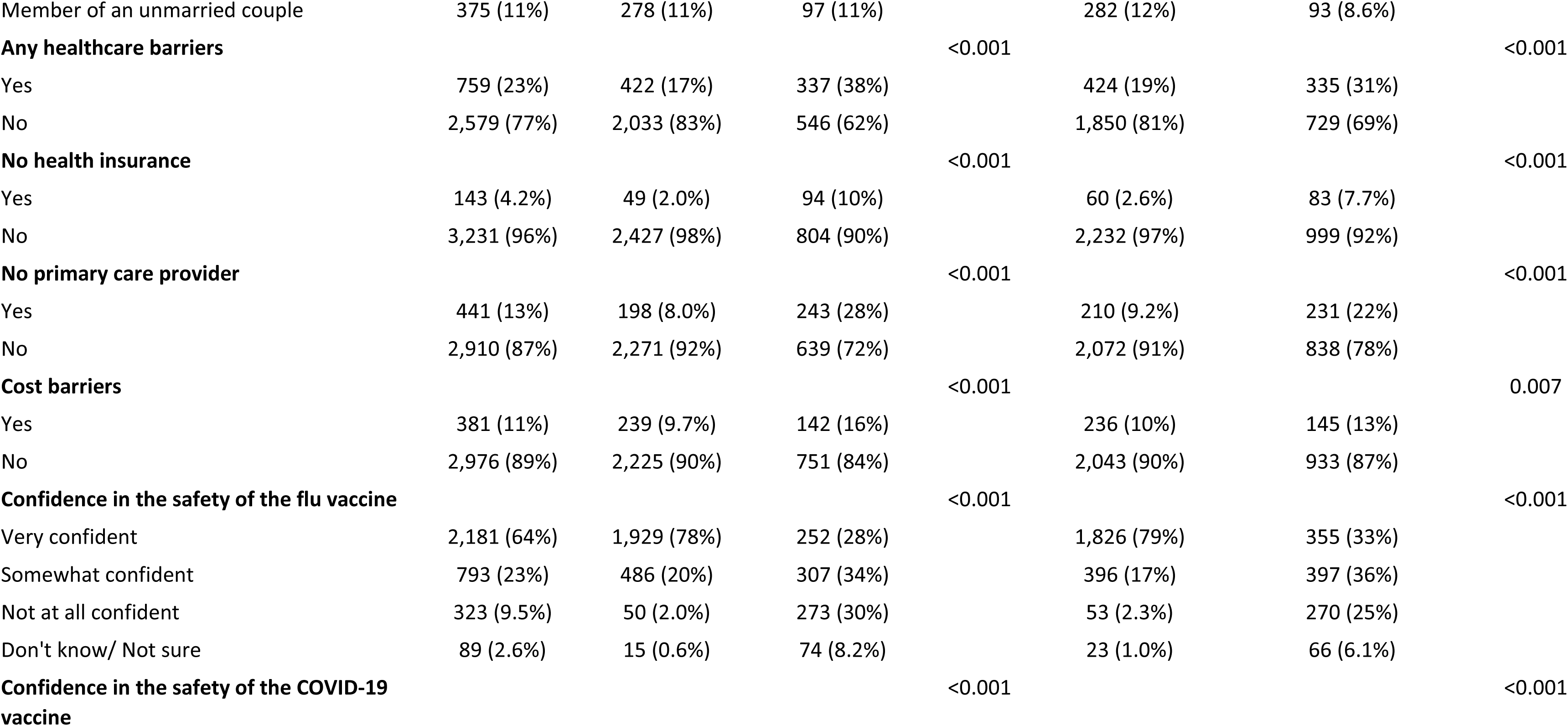
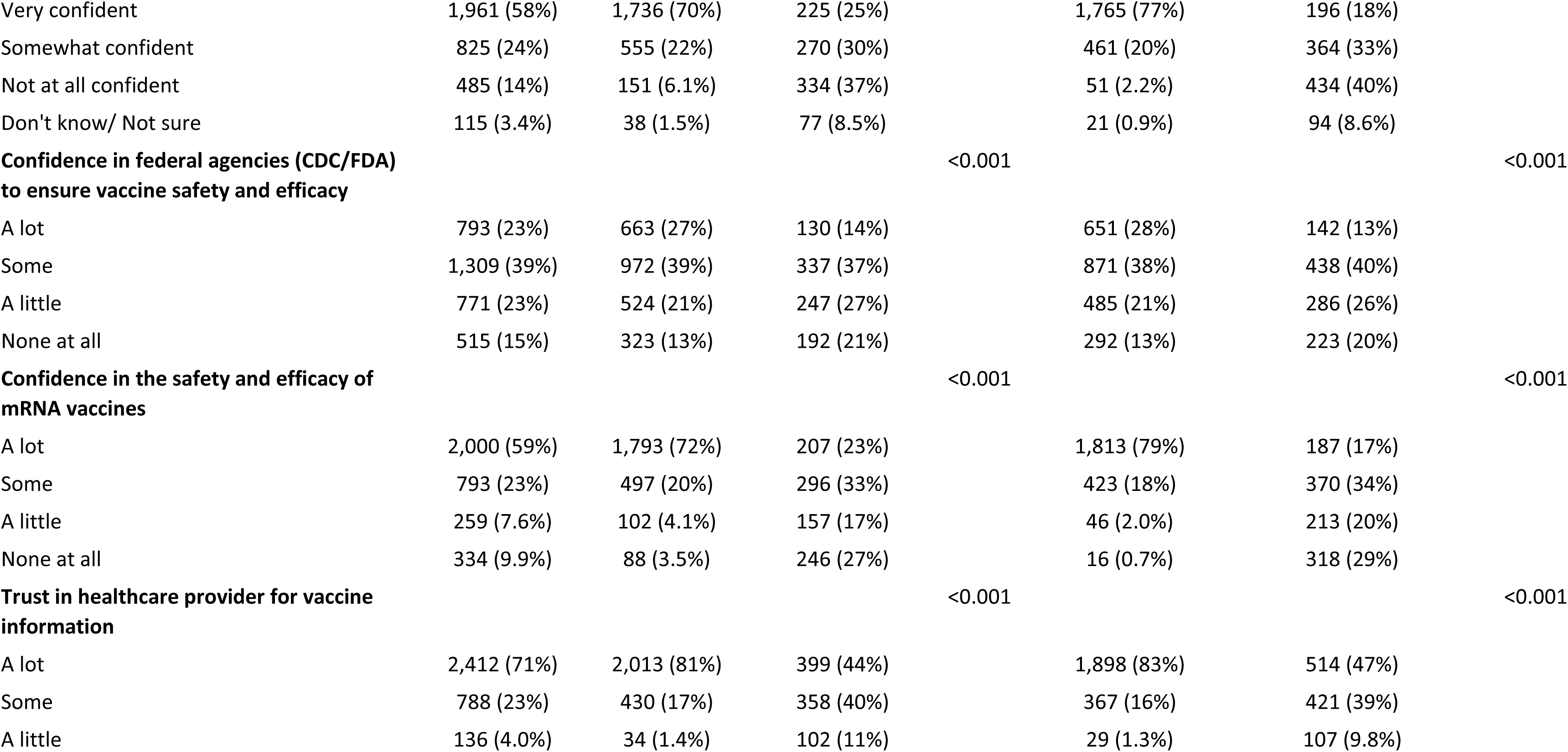
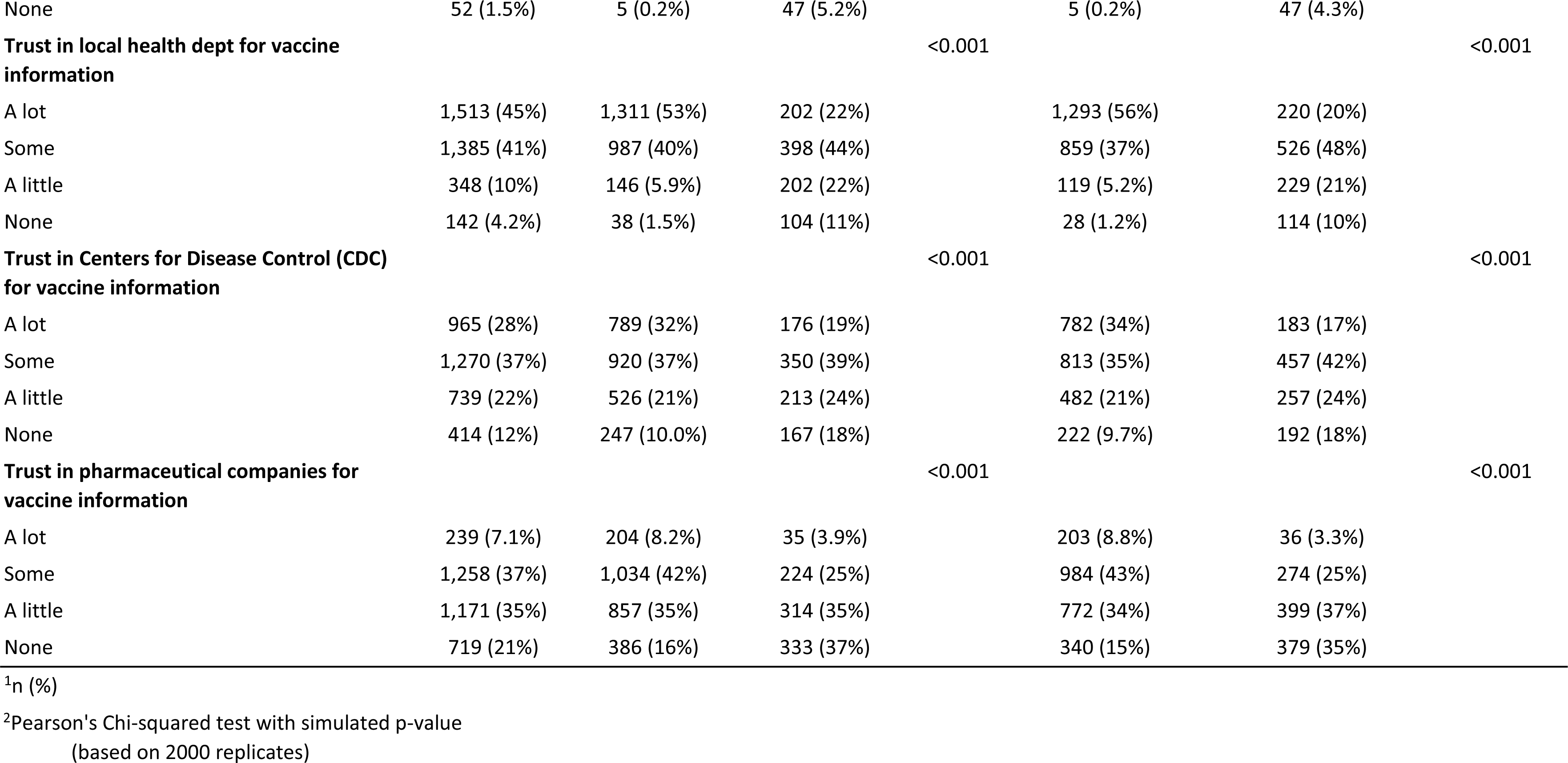
Characteristics of the analytic sample overall and by vaccine willing status, CHASING COVID Cohort (N=3,390)

### Willingness-uptake gaps and vaccination trajectories

Figure 1 shows non-vaccination during the 2025-26 respiratory virus season among participants classified as vaccine-willing in August 2025 and among the prior vaccination and intention groups used to define vaccine willingness. Among participants classified as vaccine-willing, 17% did not receive the influenza vaccine and 39% did not receive the COVID-19 vaccine by March 2026. For influenza, 16% of participants who planned to receive the vaccine and 12% of those vaccinated in the prior season did not receive the 2025-26 influenza vaccine. For COVID-19, non-vaccination varied by strength of intention: 24% of participants who definitely planned to receive the COVID-19 vaccine did not do so, compared with 66% of those who probably planned to receive it. Among participants vaccinated against COVID-19 in the prior season, 23% did not receive the 2025-26 COVID-19 vaccine.

**Figure 1.**
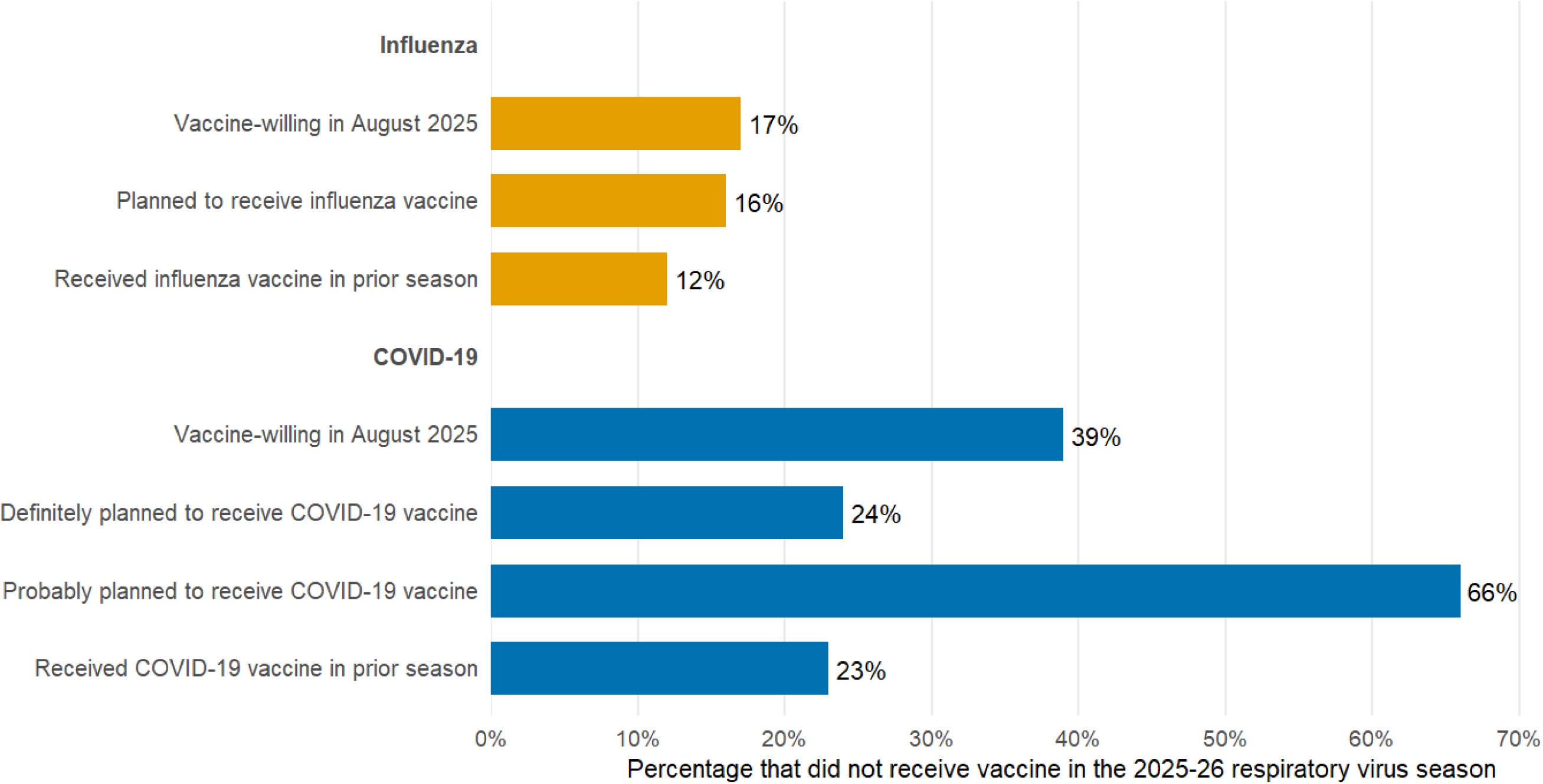
Non-vaccination during the 2025-2026 respiratory virus season among vaccine-willing participants. ***Note.** Bars show the percentage of participants in each group who did not receive the corresponding vaccine during the 2025-26 season. Denominators were 2,484 for influenza vaccine-willing participants, 2,403 for planned influenza vaccination, 2,186 for prior-season influenza vaccination, 2,300 for COVID-19 vaccine-willing participants, 1,471 for definite COVID-19 vaccination intention, 774 for probable COVID-19 vaccination intention, and 1,548 for prior-season COVID-19 vaccination.*

Figure 2 shows participant trajectories across prior vaccination history (2022-23), prior-season vaccination (2024-25), 2025-26 vaccination intention, and subsequent 2025-26 uptake. Among participants with complete influenza trajectory data (n=3,346), consistent vaccination was the dominant trajectory: 1,645 participants (49.2%) were vaccinated in both prior periods, planned to receive the 2025-26 influenza vaccine, and received it. The next largest trajectory was consistent non-vaccination: 608 participants (18.2%) were not vaccinated in either prior period, did not plan to vaccinate, and remained unvaccinated in 2025-26. An additional 373 participants (11.1%) planned to vaccinate but did not follow through, of whom 214 (57.4%) had received the influenza vaccine in 2024-25 (Figure 2; Table S2).

**Figure 2.**
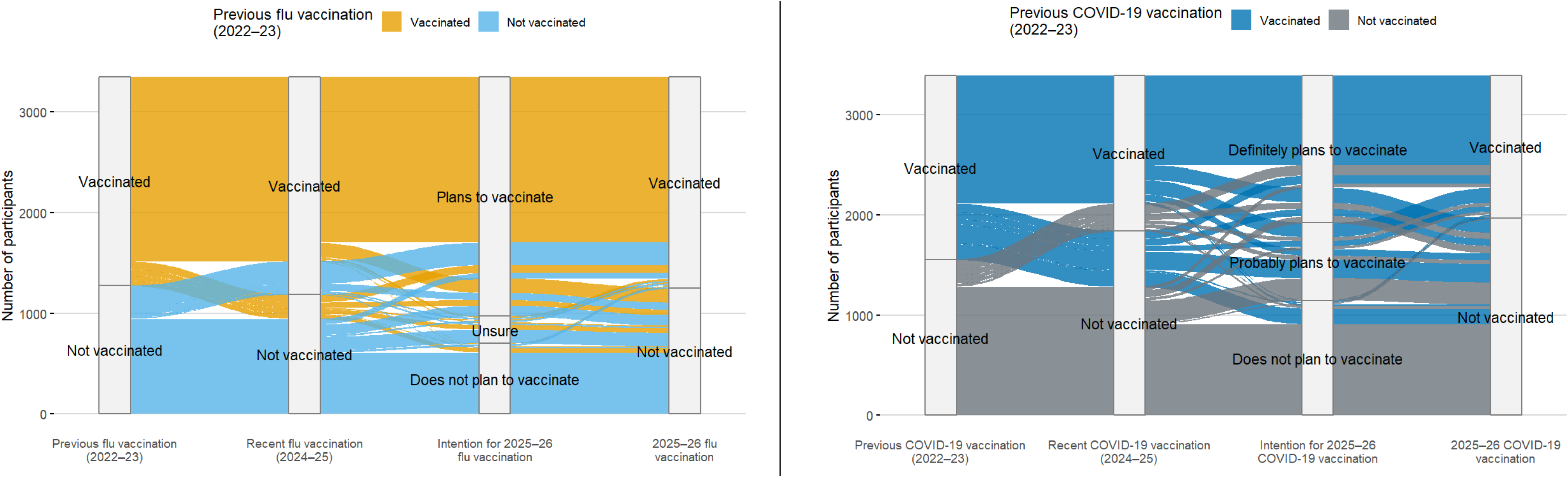
Participant trajectories across vaccination history, intention, and Fall 2025-Winter 2026 vaccination, CHASING COVID Cohort (N=3,390)

Among participants with complete COVID-19 trajectory data (n=3,390), trajectories were more evenly split between consistent vaccination and consistent non-vaccination than observed for influenza: 891 participants (26.3%) were consistently vaccinated and 907 (26.8%) were consistently unvaccinated. Among the next largest COVID-19 trajectories were several groups who reported definite or probable plans to vaccinate but remained unvaccinated in 2025-26, including participants who had not been vaccinated in either prior period but probably planned to vaccinate (n=222; 6.5%), those vaccinated in 2022-23 but not in 2024-25 who probably planned to vaccinate (n=183; 5.4%), and those vaccinated in both prior periods who definitely planned to vaccinate (n=148; 4.4%) (Figure 2; Table S3).

In supplementary analyses among adults aged ≥65 years, trajectories showed high consistent influenza vaccination but continued COVID-19 willingness-uptake gaps (Figure S1). Among older adults with complete influenza trajectory data (n=592), 417 (70.4%) were consistently vaccinated, 69 (11.7%) were consistently unvaccinated, and 31 (5.2%) planned to receive the 2025-26 influenza vaccine but did not do so (Table S4). Among older adults with complete COVID-19 trajectory data (n=595), 268 (45.0%) were consistently vaccinated, 117 (19.7%) were consistently unvaccinated, and 100 (16.8%) definitely or probably planned to receive the 2025-26 COVID-19 vaccine but remained unvaccinated (Table S5).

### Predictors of not vaccinating among vaccine-willing participants

Among vaccine-willing participants, 424 of 2,484 (17.1%) did not receive the 2025-26 influenza vaccine and 904 of 2,300 (39.3%) did not receive the 2025-26 COVID-19 vaccine (Table 2). The strongest predictor of not vaccinating was absence of prior-season vaccination: vaccine-willing participants who were not vaccinated in 2024-25 were more likely to remain unvaccinated in 2025-26 for both influenza (aRR=4.04, 95% CI: 3.44, 4.74) and COVID-19 (aRR=3.01, 95% CI: 2.72, 3.34). Non-vaccination was also higher among younger adults, participants with lower educational attainment and lower household income, and Hispanic and Black non-Hispanic participants compared with their respective reference groups; the largest demographic associations were observed for adults aged 18-29 years versus 60 years or older, household income less than $25,000 versus $200,000 or more, and high school education or less versus college graduate. Healthcare access barriers were consistently associated with lower uptake despite willingness, including lack of insurance, cost barriers, and no primary care provider; having no primary care provider was associated with a higher risk of not vaccinating for both influenza (aRR=1.96, 95% CI: 1.57, 2.45) and COVID-19 (aRR=1.49, 95% CI: 1.32, 1.69). Lower vaccine confidence was also associated lower uptake for both vaccines, with the strongest confidence-related associations observed among those not at all confident in the safety of the influenza vaccine (aRR=2.95, 95% CI: 2.15, 4.05) or COVID-19 vaccine (aRR=2.21, 95% CI: 1.88, 2.59), compared with those who were very confident. Associations with trusted sources of vaccine information were less consistent; greater trust in healthcare providers was associated with higher influenza vaccine uptake, while trust in local health departments and pharmaceutical companies was not clearly associated with uptake for either vaccine.

**Table 2.**
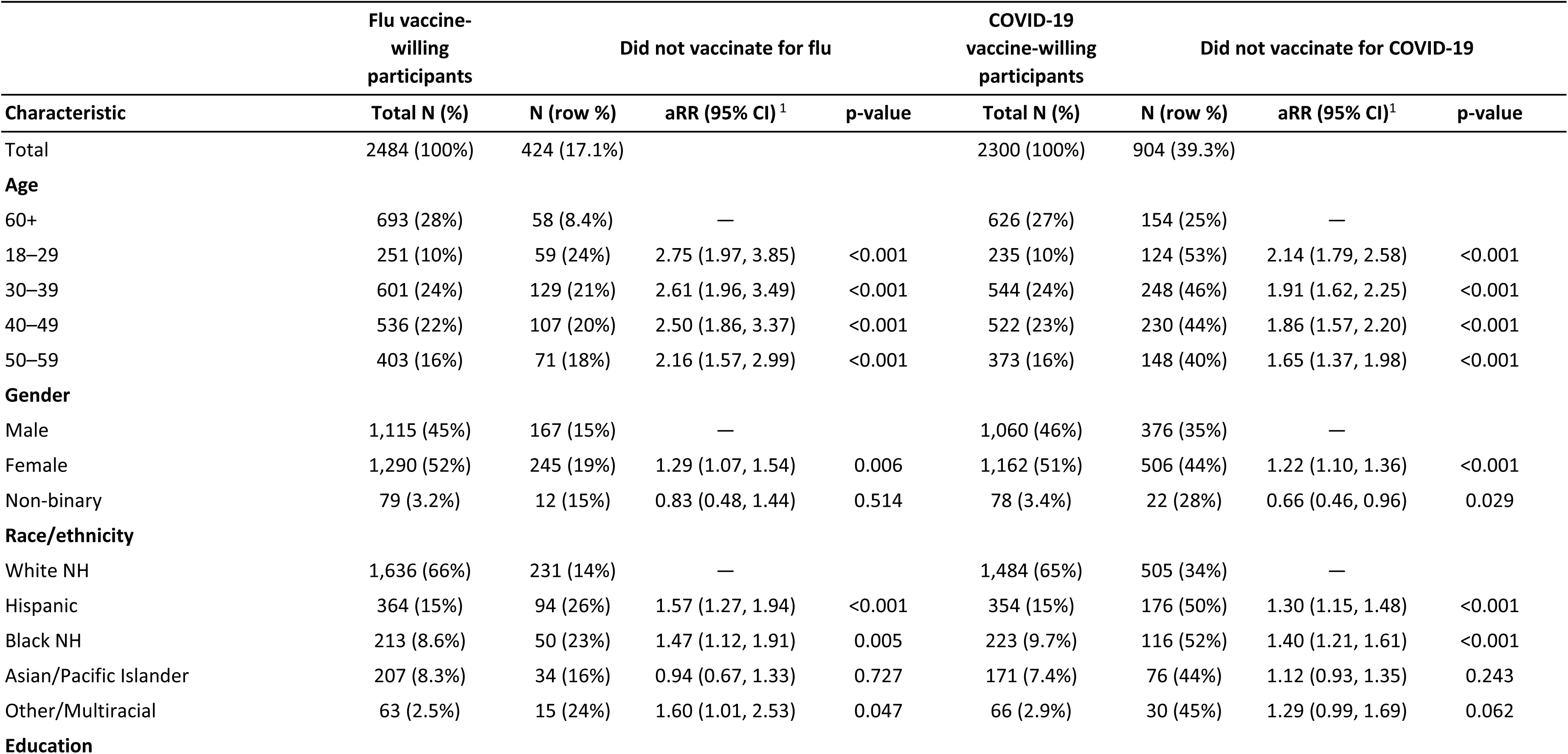
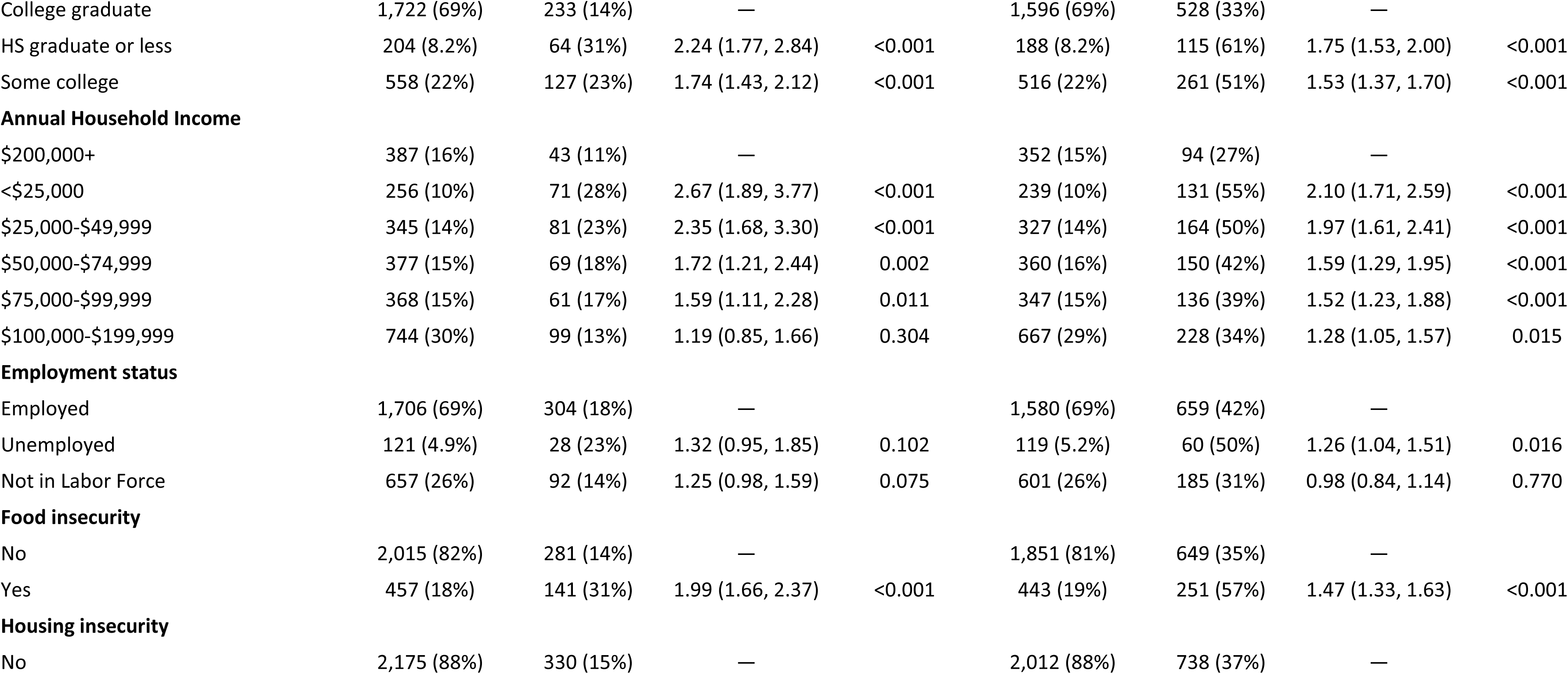
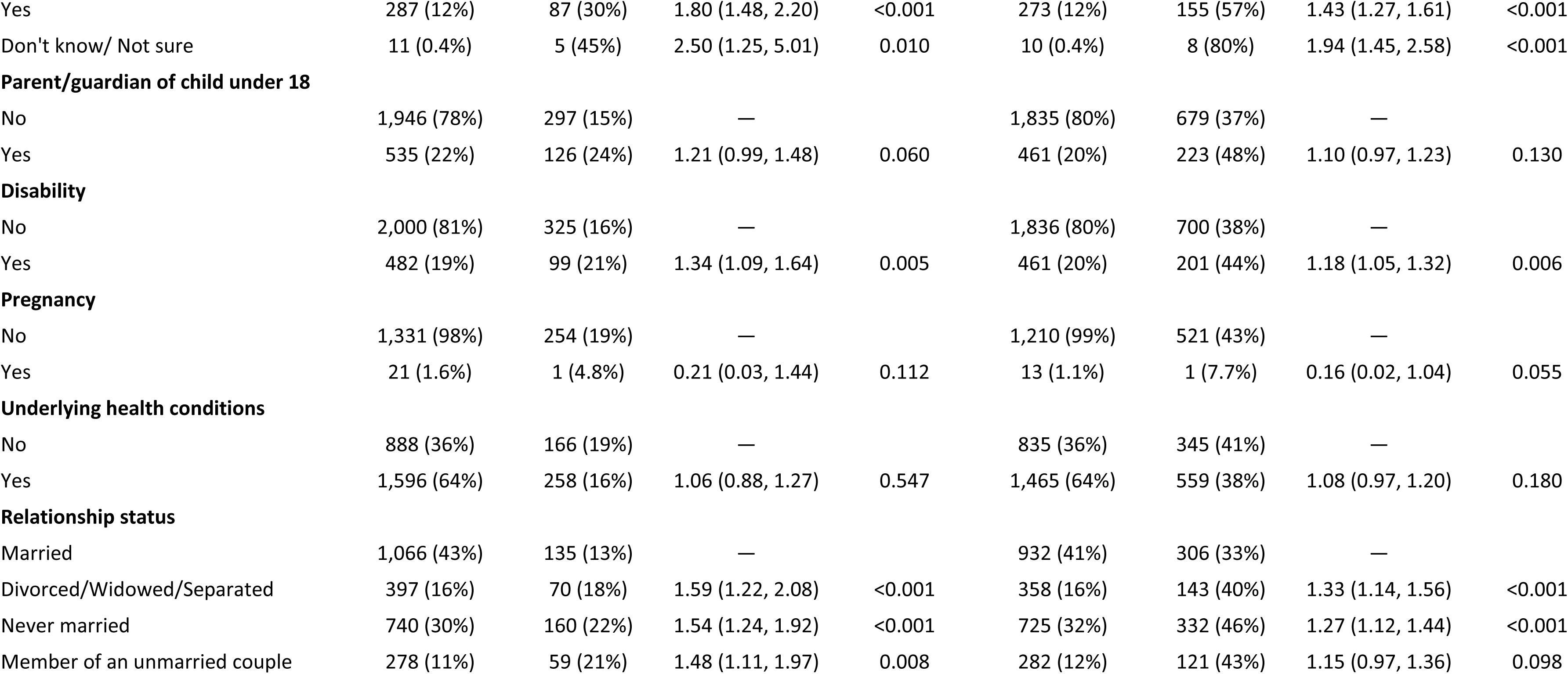
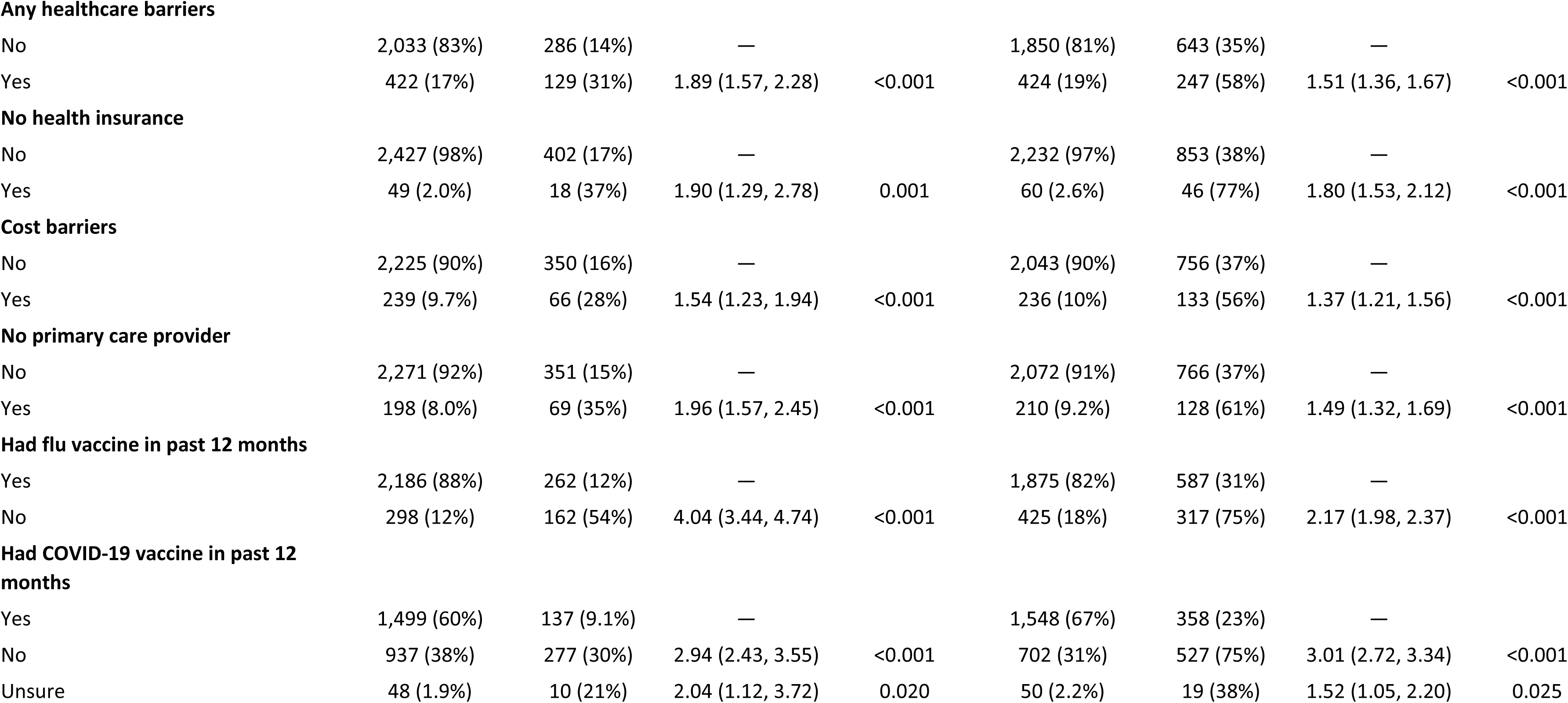
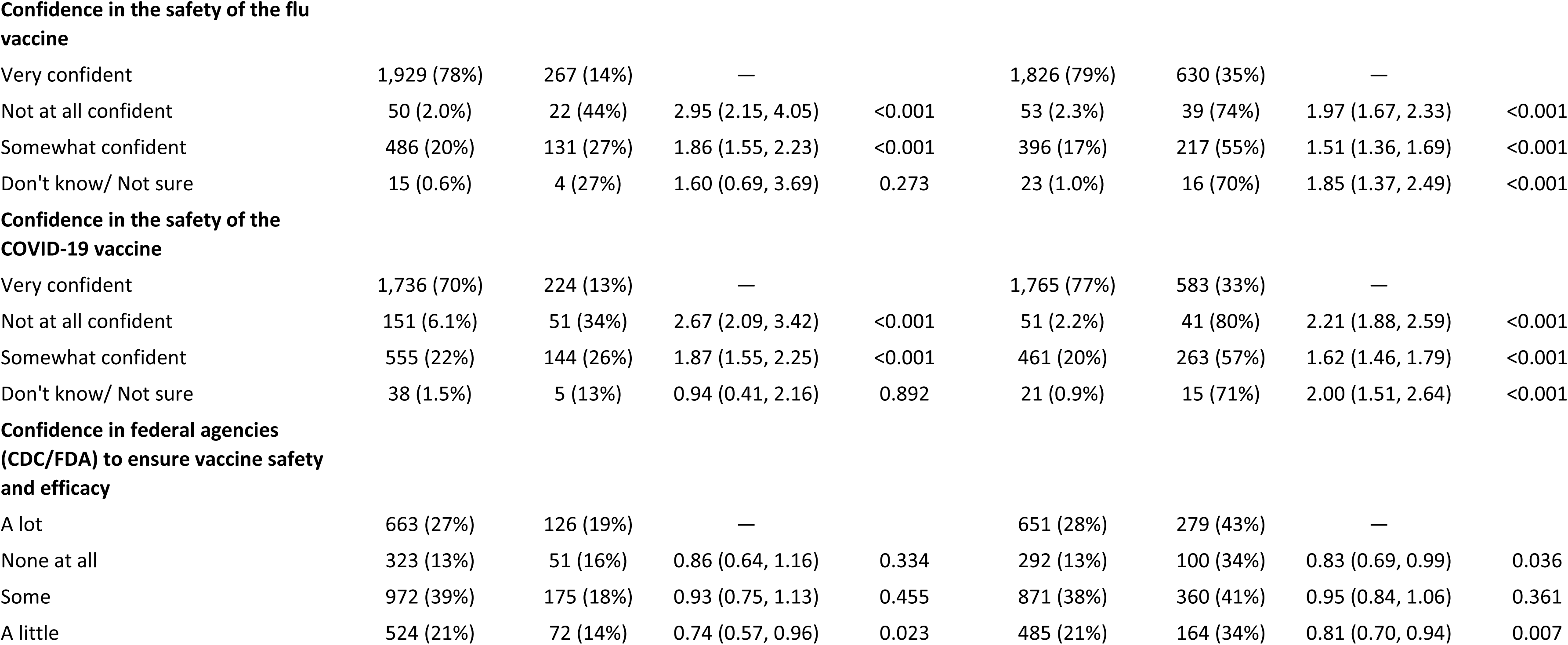
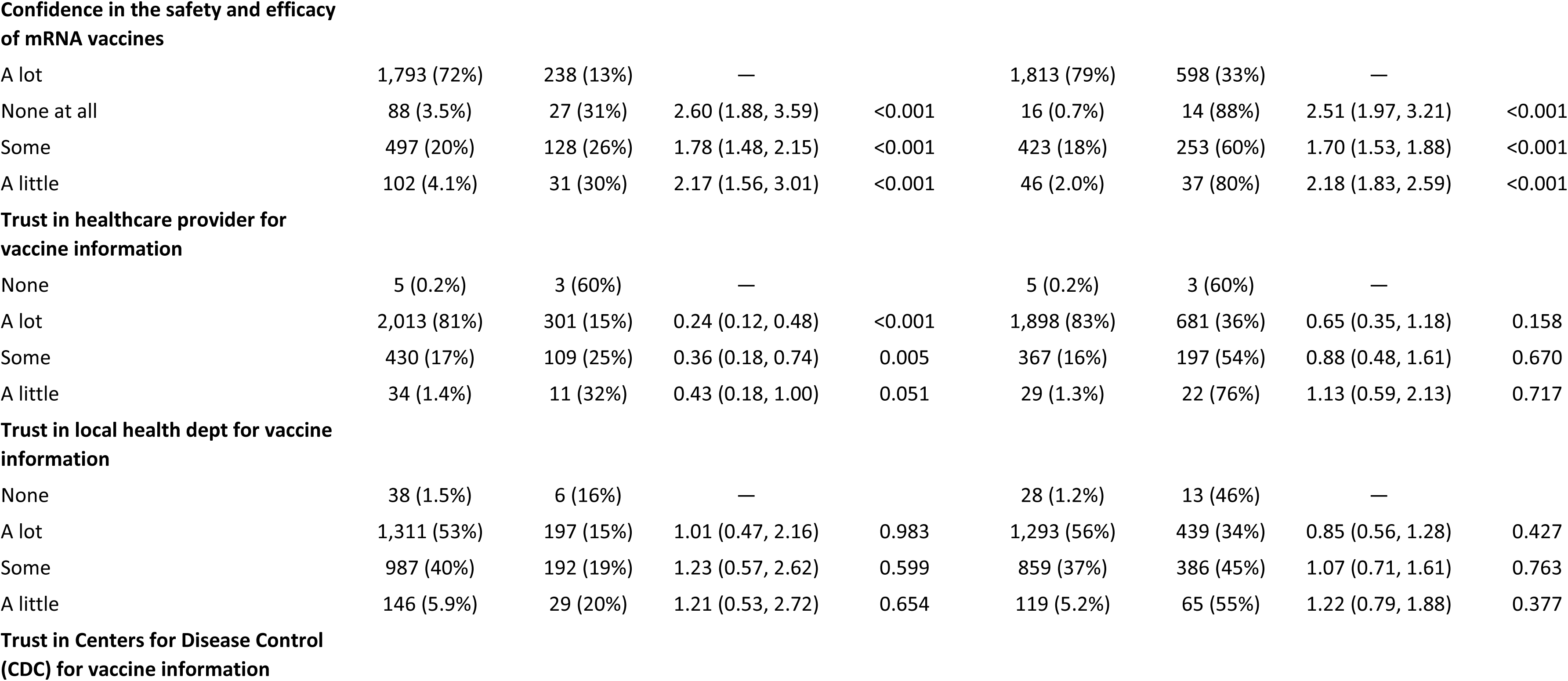
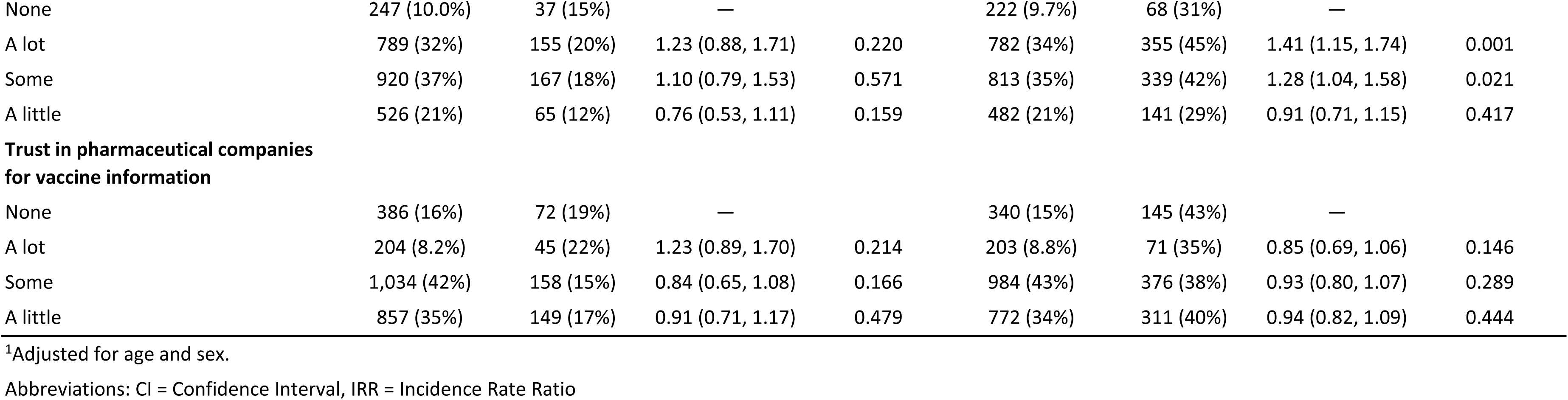
Predictors of 2025-26 vaccination non-uptake among vaccine-willing participants, CHASING COVID Cohort (N=3,390)

### Reasons for non-vaccination

Among participants who did not receive the 2025-26 influenza vaccine, reasons for non-vaccination differed by vaccine-willing status (Table 3A). The reason most endorsed among influenza vaccine-willing participants was inability to find a convenient time, place, or appointment, reported by 33.3% of vaccine-willing non-vaccinators compared with 7.5% of non-willing non-vaccinators. In contrast, non-willing participants more commonly reported concern about side effects (29.8% vs. 10.1% among vaccine-willing participants), believing they could stay healthy in other ways (25.8% vs. 8.3%), and lack of trust in vaccine safety (22.6% vs. 4.7%). For COVID-19 vaccination, reasons for non-vaccination also differed by vaccine-willing status (Table 3B). Compared with non-willing participants, COVID-19 vaccine-willing non-vaccinators more commonly reported inability to find a convenient time, place, or appointment (24.2% vs. 4.7%), lack of healthcare provider recommendation (16.7% vs. 9.9%), cost or insurance barriers (8.1% vs. 1.8%), and not knowing where to get the vaccine (7.4% vs. 1.6%). In contrast, non-willing participants more commonly reported concern about side effects (40.9% vs. 11.3% among vaccine-willing participants), lack of trust in vaccine safety (33.8% vs. 4.3%), and believing they could stay healthy in other ways (19.6% vs. 6.7%).

**Table 3A.**
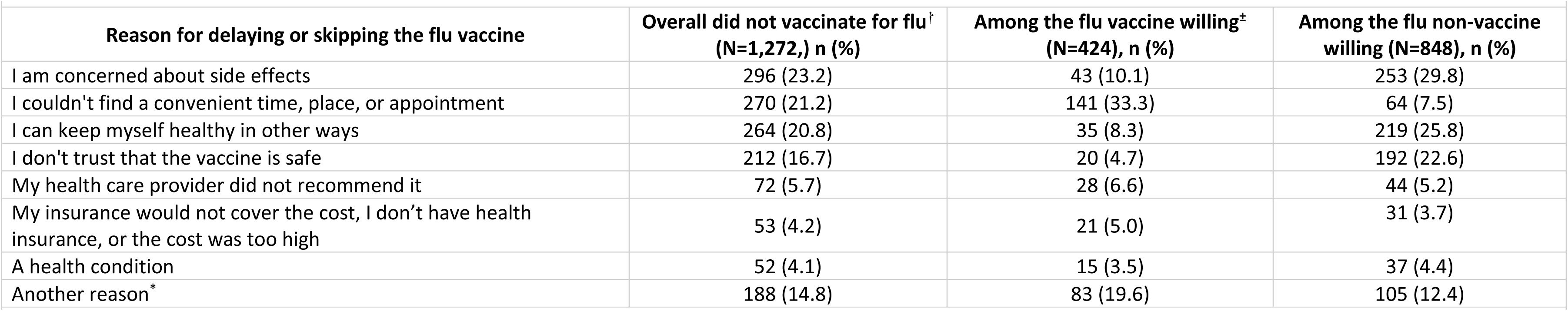
Reasons for not vaccinating for influenza among participants who did not receive influenza vaccination between September 2025 and March 2026, overall and among those who intended to vaccinate or had received influenza vaccine in the prior 12 months.

**Table 3B.**
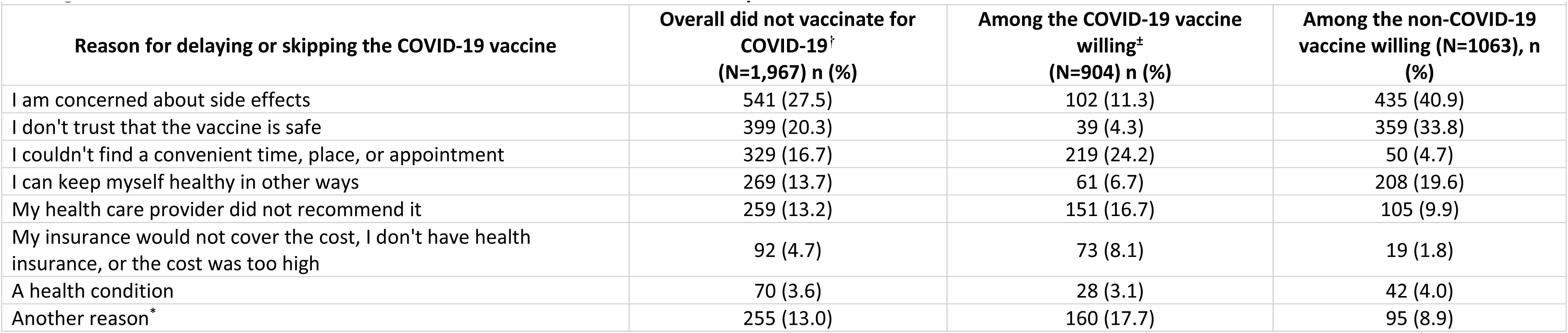
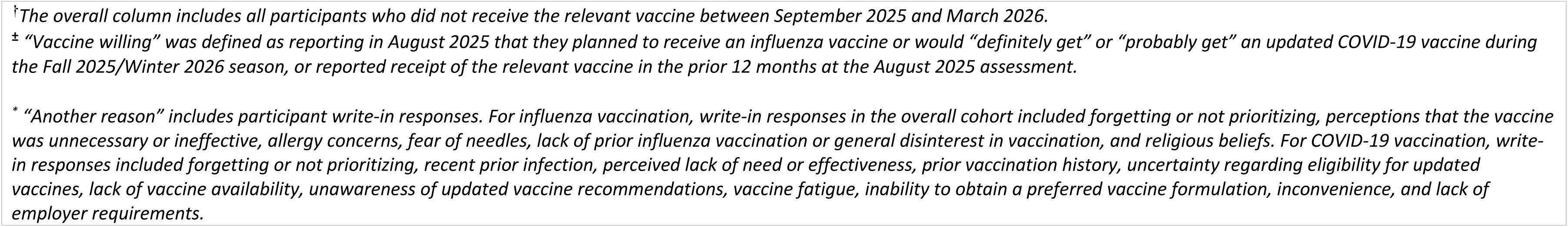
Reasons for not vaccinating for COVID-19 among participants who did not receive COVID-19 vaccination between September 2025 and March 2026, overall and among those who intended to vaccinate or had received COVID-19 vaccine in the prior 12 months.

Among participants who did not receive the respective vaccine, 185 (14.5%) influenza non-vaccinators and 251 (12.8%) COVID-19 non-vaccinators provided a write-in response for “another reason.” For both vaccines, write-in reasons included forgetting or not prioritizing vaccination and perceived lack of need or effectiveness for both vaccines. Influenza-specific write-in responses included allergy concerns, fear of needles, lack of prior influenza vaccination or general disinterest in vaccination, and religious beliefs. COVID-19-specific write-in responses included recent prior infection, prior vaccination history, uncertainty regarding eligibility for updated vaccines, lack of vaccine availability, unawareness of updated vaccine recommendations, vaccine fatigue, inability to obtain a preferred vaccine formulation, inconvenience, and lack of employer requirements.

## Discussion

In this national longitudinal cohort, we found notable gaps between vaccine willingness and vaccine uptake during the 2025-26 respiratory virus season, particularly for COVID-19 vaccination. Among participants classified as vaccine willing in August 2025, 17% remained unvaccinated for influenza and 39% remained unvaccinated for COVID-19 by March 2026. Uptake was especially low among participants without prior-season vaccination and those facing material hardship or healthcare access barriers. Reported reasons for non-vaccination among vaccine-willing participants suggest that many of these gaps reflected addressable barriers to follow-through, including difficulty finding a convenient time, place, or appointment and, for COVID-19, lack of healthcare provider recommendation. These findings align with prior work showing that health insurance coverage, provider recommendations, and convenience are important determinants of adult vaccination uptake.^35–41^ They also point to missed opportunities to improve uptake by reducing logistical and cost-related barriers, strengthening provider recommendations, and making vaccination easier to complete for adults already open to vaccination.

Among vaccine-willing participants, lack of prior-season vaccination was the strongest predictor of non-vaccination, consistent with existing literature.^42,43^ Participants who had not received the corresponding vaccine in 2024-25 were approximately four times as likely to remain unvaccinated for influenza and three times as likely to remain unvaccinated for COVID-19. Willingness may therefore be less likely to translate into uptake when vaccination is not already part of an established routine.^44^ Trajectory analyses further clarified where gaps emerged across the historical vaccination pathway. For influenza, sustained vaccination across seasons was the dominant trajectory, suggesting relatively stable routine vaccination behavior. For COVID-19, trajectories were more evenly divided between sustained vaccination and sustained non-vaccination, and several common pathways ended in non-vaccination despite stated intention, prior vaccination history, or both. This contrast may reflect, in part, the fact that COVID-19 vaccination is newer and less routinized than seasonal influenza vaccination and has been subject to more frequent changes in recommendations and public messaging.^31,45^ Importantly, gaps were also observed among adults aged ≥65 years, with 17% of those who definitely or probably planned to receive the COVID-19 vaccine remaining unvaccinated. This suggests missed opportunities for protection among a group for whom vaccination is particularly important, and is consistent with prior literature identifying cost, transportation, mobility, and scheduling barriers as constraints on vaccination follow-through among older adults.^46^

Non-vaccination among vaccine-willing participants was patterned by sociodemographic characteristics, material hardship, and healthcare access. Younger adults, participants with lower education and income, and Hispanic and Black non-Hispanic participants, and those reporting food insecurity, housing insecurity, lack of insurance, cost barriers, or no primary care provider had higher non-vaccination despite willingness. These findings are consistent with prior studies documenting socioeconomic and racial and ethnic disparities in adult vaccination uptake and suggest that willingness-uptake gaps may often reflect structural and logistical barriers.^47–49^ Reported reasons for non-vaccination further support this interpretation: vaccine-willing non-vaccinators more commonly reported difficulty finding a convenient time, place, or appointment and, for COVID-19 cost or insurance barriers and not knowing where to get vaccinated. Write-in responses, including forgetting or not prioritizing vaccination, perceived lack of need, and vaccine fatigue, also suggest that some non-uptake among vaccine-willing adults may reflect ambivalence or low urgency. By contrast, participants who were not vaccine willing more often cited side effect concerns, lack of trust in vaccine safety, and beliefs that they could remain healthy in other ways, consistent with prior research.^40,50,51^ These patterns suggest adults already open to vaccination may benefit from strategies that reduce logistical barriers and help translate openness into action, such as routine provider offers, community-, pharmacy-, and workplace-based vaccination opportunities, reminder systems, clearer information about eligibility and insurance coverage, and continued reinforcement of vaccine safety and effectiveness.^52–54^

The gaps observed in this cohort are notable given that participants had higher vaccination uptake than national estimates during the 2025-26 season: 62% received an influenza vaccine and 42% received a COVID-19 vaccine, compared with national estimates of 44% and 16%, respectively.^6^ Given their willingness to participate in health research, they are likely to be more engaged with health information and preventive care than the general U.S. adult population. The cohort therefore provides a relatively favorable setting in which to examine whether adults who appear open to vaccination ultimately follow through. Yet even in this comparatively health-engaged group, many participants who had previously vaccinated or intended to vaccinate did not receive the 2025-26 COVID-19 and influenza vaccines. The barriers identified here are therefore likely to remain relevant among adults in the broader population who are similarly open to vaccination but do not follow through.

During the shifting 2025-26 vaccine policy and communication environment, reasons reported by COVID-19 vaccine-willing non-vaccinators suggest that some non-uptake may have reflected confusion about recommendations and access. Participants reported uncertainty about eligibility, lack of vaccine availability, unawareness of updated recommendations, and inability to obtain a preferred vaccine formulation. Although we cannot determine whether contemporaneous policy or communication changes caused these barriers, these responses point to actionable breakdowns in the path from willingness to uptake, highlighting the need for clear information about eligibility, coverage, availability, and where and how to get vaccinated.

Strengths of this study include its prospective longitudinal design, diverse national community-based sample, and ability to link prior vaccination history, intention, subsequent uptake, and reasons for non-vaccination within the same cohort. Several limitations should also be considered. Vaccination status was self-reported and may be subject to recall error. Our definition of vaccine willingness intentionally combined prior-season vaccination behavior and stated intention to identify participants with evidence of openness to vaccination, but these groups may differ in the strength or stability of their willingness. Finally, although the cohort was geographically and socio-demographically diverse and provided an informative sample for examining follow-through among adults open to vaccination, findings may not be fully generalizable to all U.S. adults. Because assessments were completed online and in English, findings may underrepresent adults with limited internet access, lower digital literacy, or limited English proficiency.

Findings from this study underscore that vaccine willingness does not necessarily translate into action. During the 2025-26 respiratory virus season, non-vaccination among vaccine-willing participants was especially common for COVID-19 vaccination and was associated with prior non-vaccination, structural and healthcare access barriers, and lower confidence in vaccine safety. Some willingness-uptake gaps may reflect logistical or structural barriers, while others may reflect underlying ambivalence. That substantial gaps remained even in a cohort with relatively high vaccination uptake points to missed opportunities among adults with prior or stated openness to vaccination. Improving coverage may therefore require strategies that help vaccine-willing adults move from openness to completed vaccination by reducing access barriers, reinforcing confidence and perceived need, and providing clear guidance about when, where, and how to get vaccinated.

## Acknowledgements

We thank the participants of the Communities, Households, and SARS-CoV-2 Epidemiology COVID Cohort Study for their contribution to the advancement of science.

## CRediT Author Statement

**Jenna Sanborn:** Methodology, Software, Formal analysis, Investigation, Writing-Original Draft. **McKaylee Robertson:** Methodology, Writing-Review & Editing. **Kate Penrose**: Writing-Review & Editing. **Madhura Rane:** Writing-Review and Editing. **Rachael Piltch-Loeb:** Writing-Review and Editing. **Angela M Parcesepe**: Writing-Review & Editing. **Denis Nash**: Conceptualization, Methodology, Writing-Review & Editing, Funding acquisition.

All authors attest they meet the ICMJE criteria for authorship

## Data availability statement

The datasets generated and/or analyzed during the current study are not publicly available due to funder requirements, but are available from the authors upon reasonable request, subject to approval and available resources.

## Declaration of generative AI and AI-assisted technologies in the manuscript preparation process

During the preparation of this work, the authors used ChatGPT™ (OpenAI, San Francisco, CA, USA) to improve the readability and language of the work and to support statistical coding. The authors critically evaluated and revised all content and code and take full responsibility for the published work.

## SUPPLEMENTAL MATERIALS

**Supplemental Table S1.**
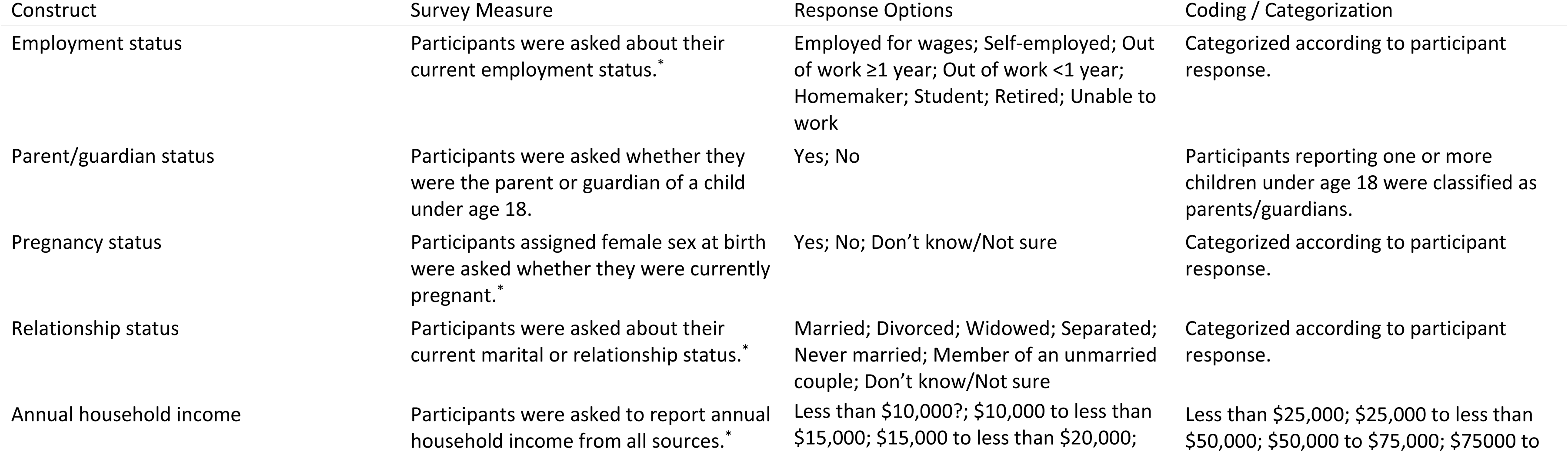
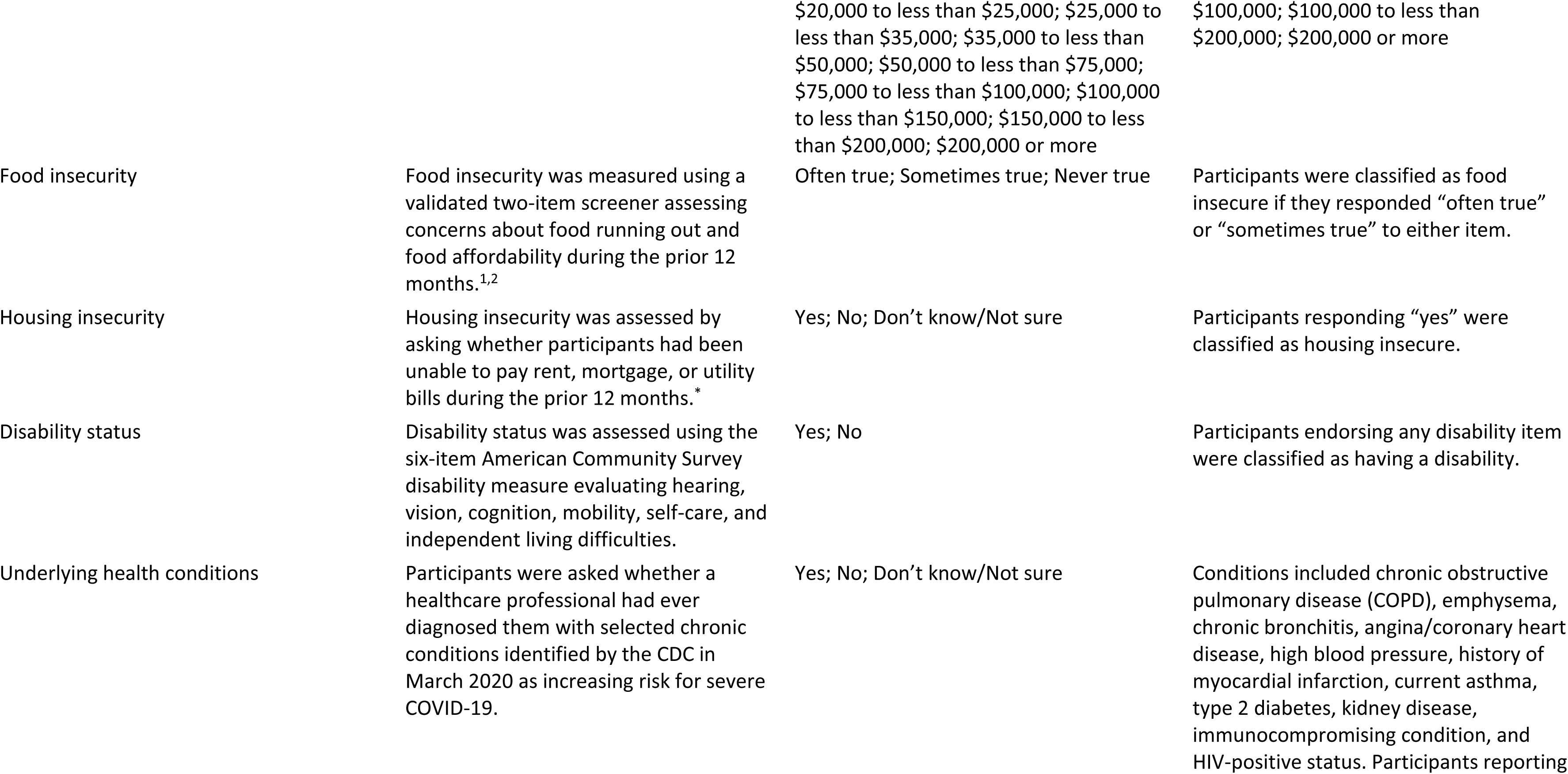
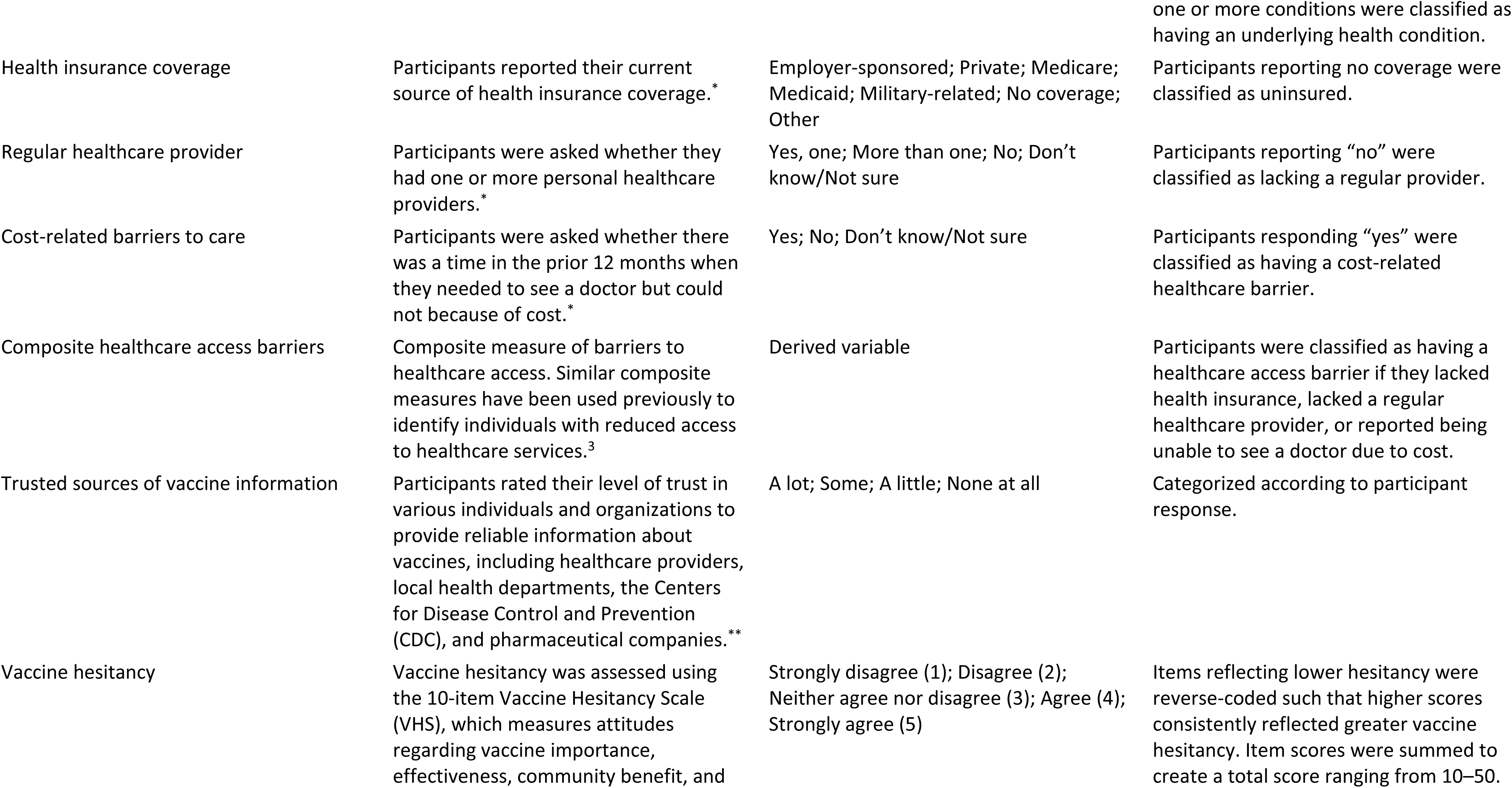
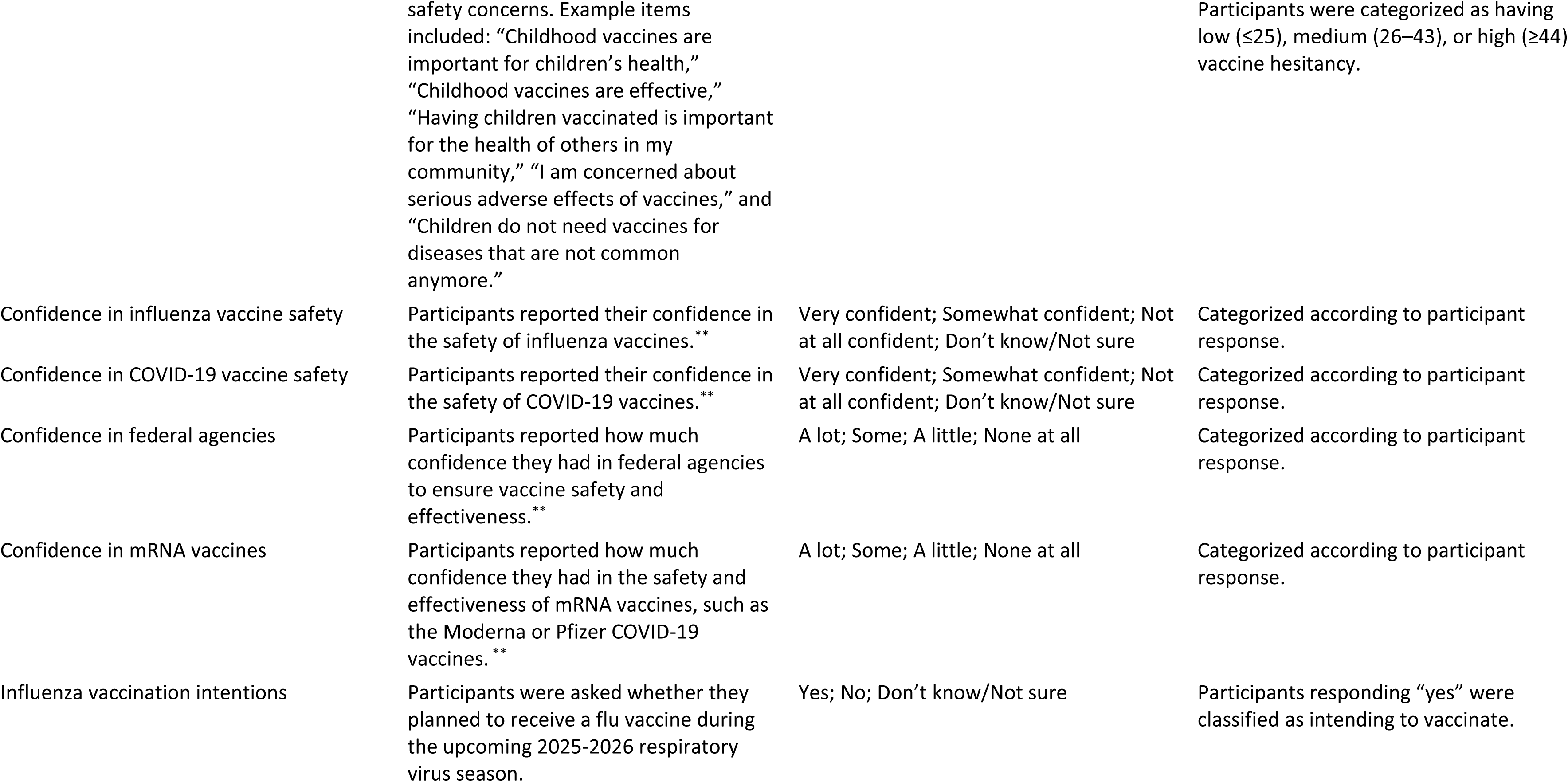
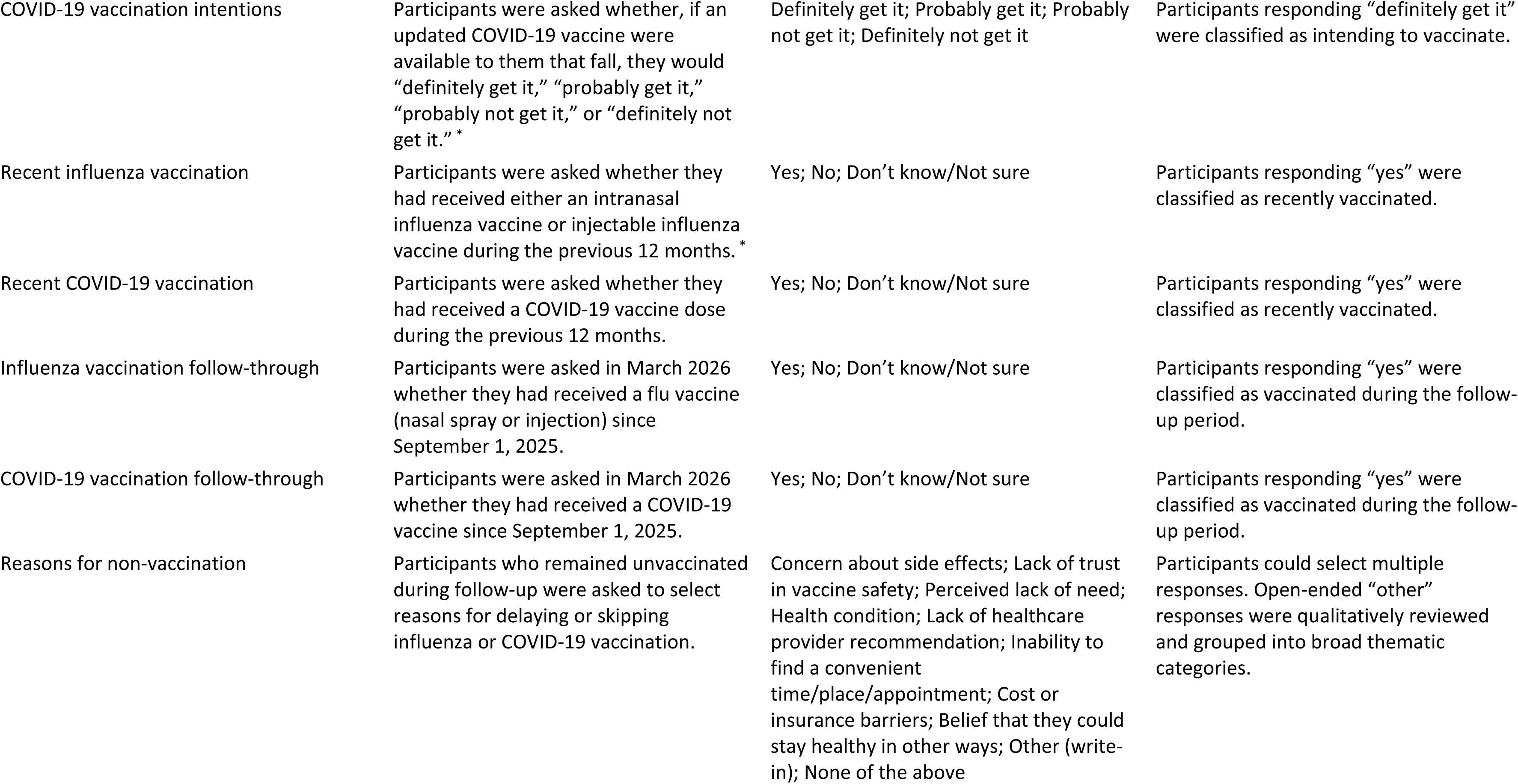
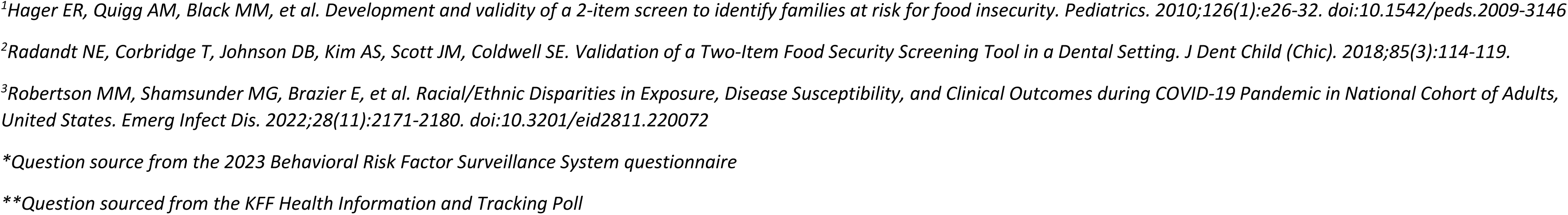
Vaccine-Related Survey Measures, Response Options, and Variable Definitions.

**Supplemental Table S2.**
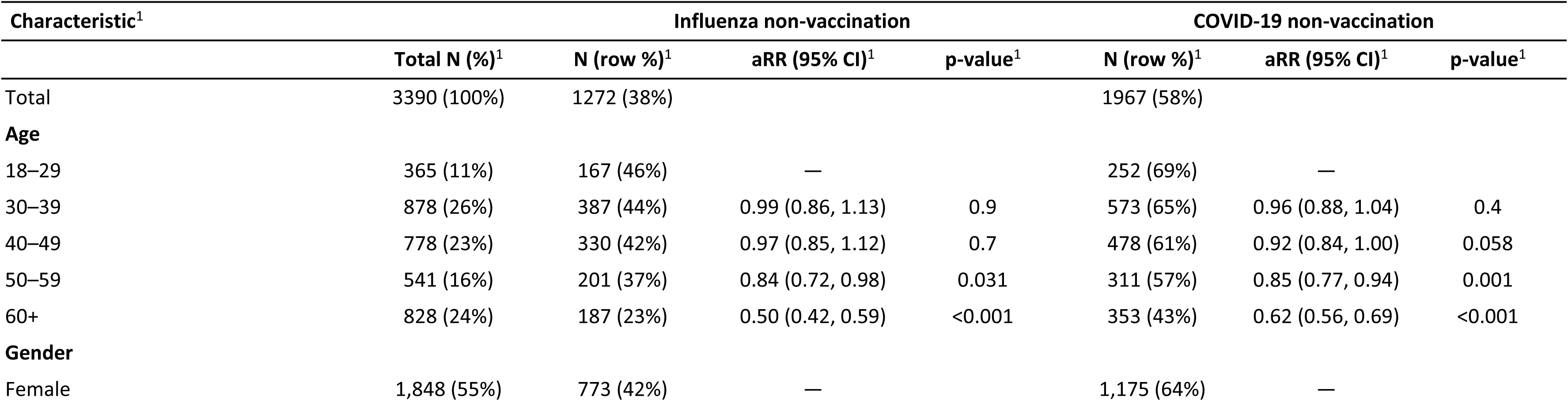
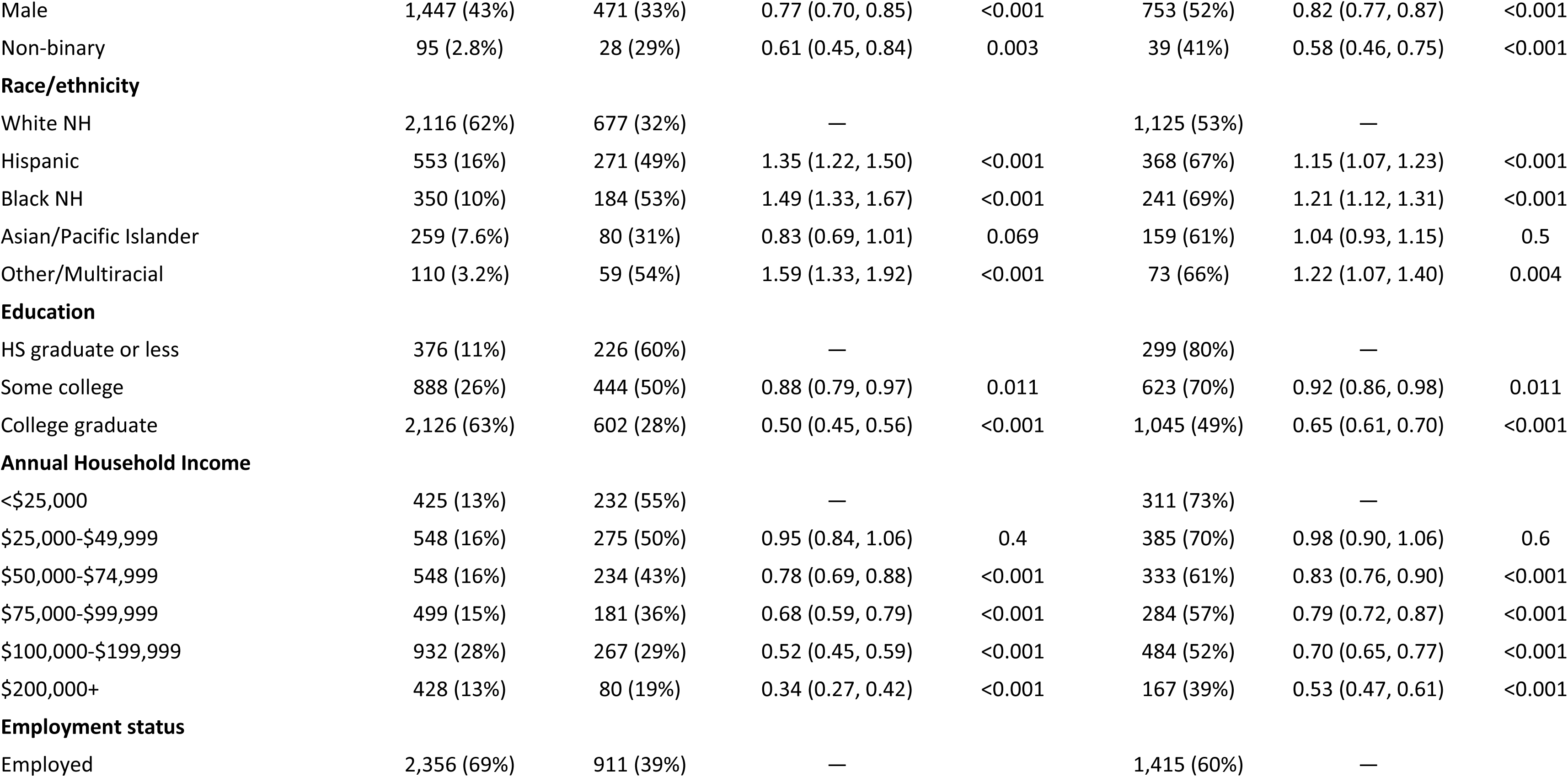
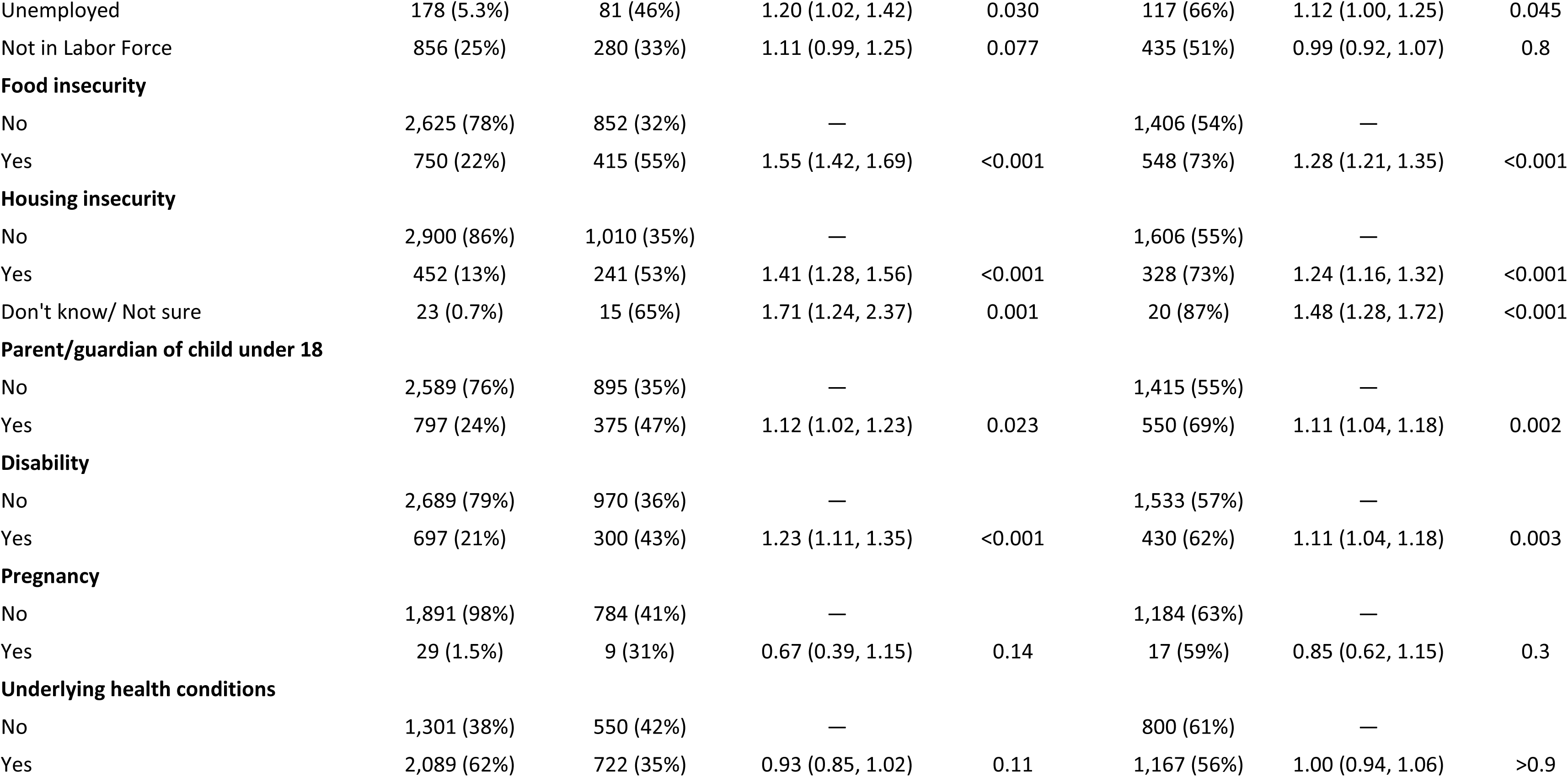
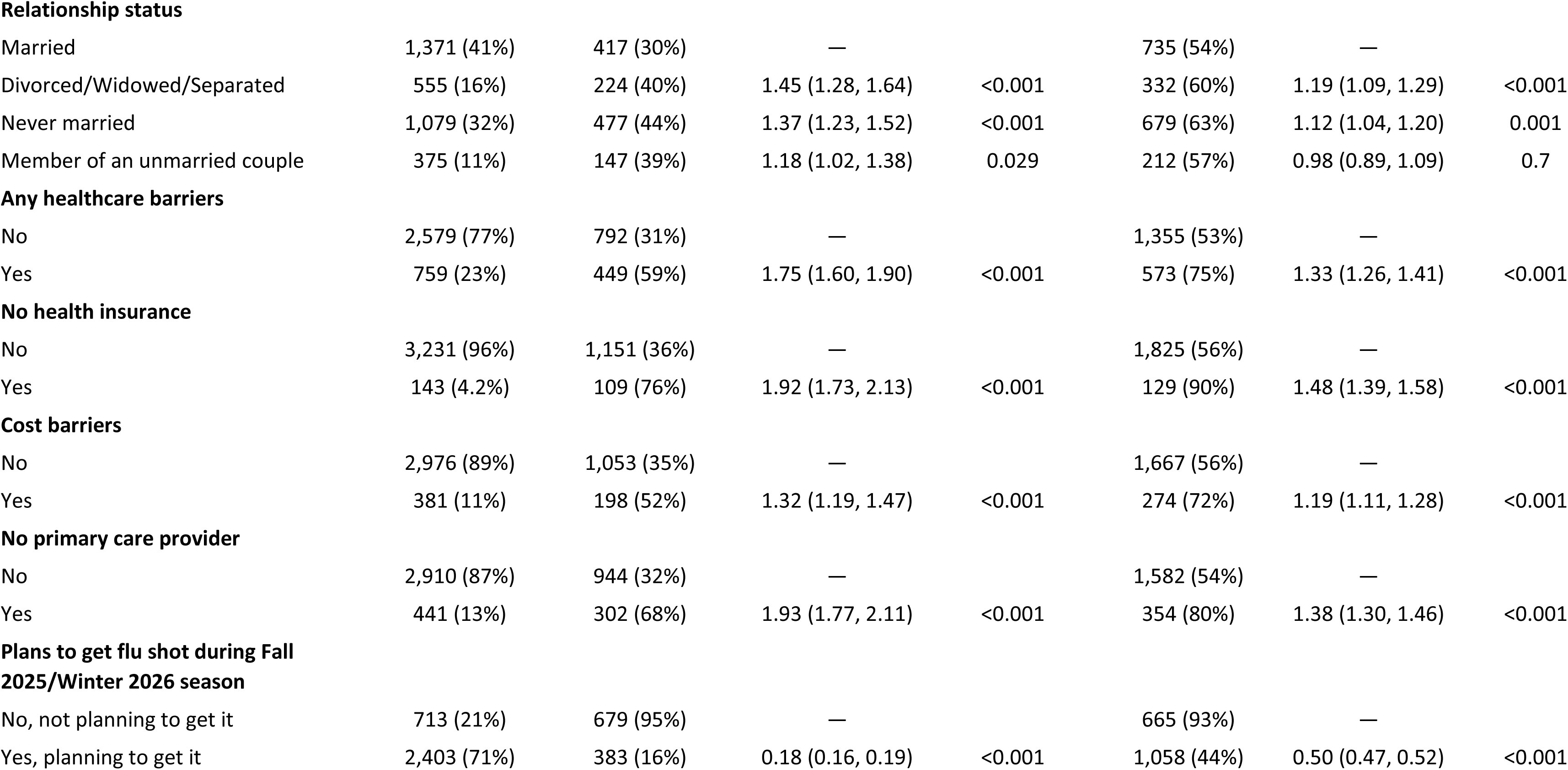
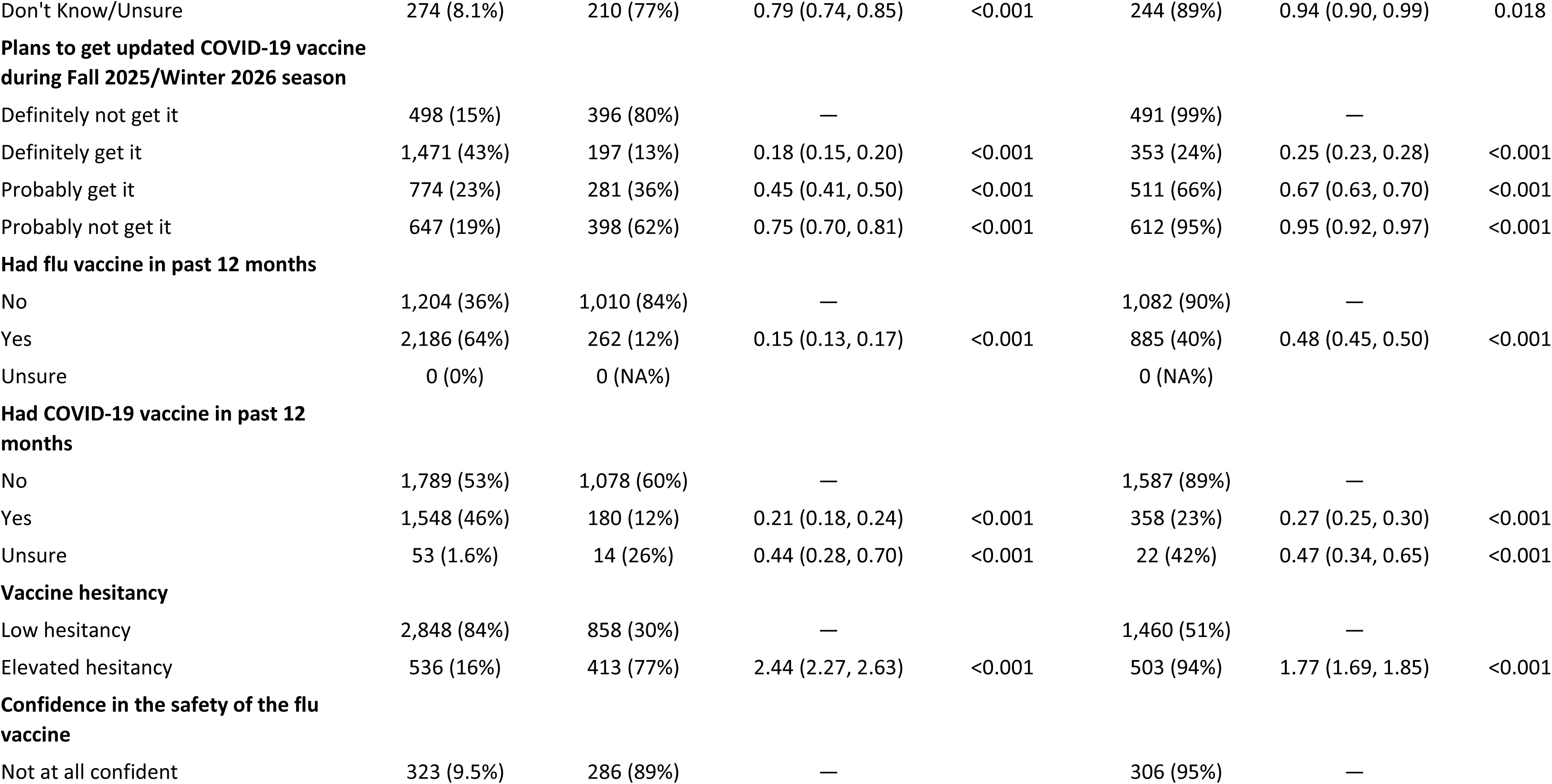
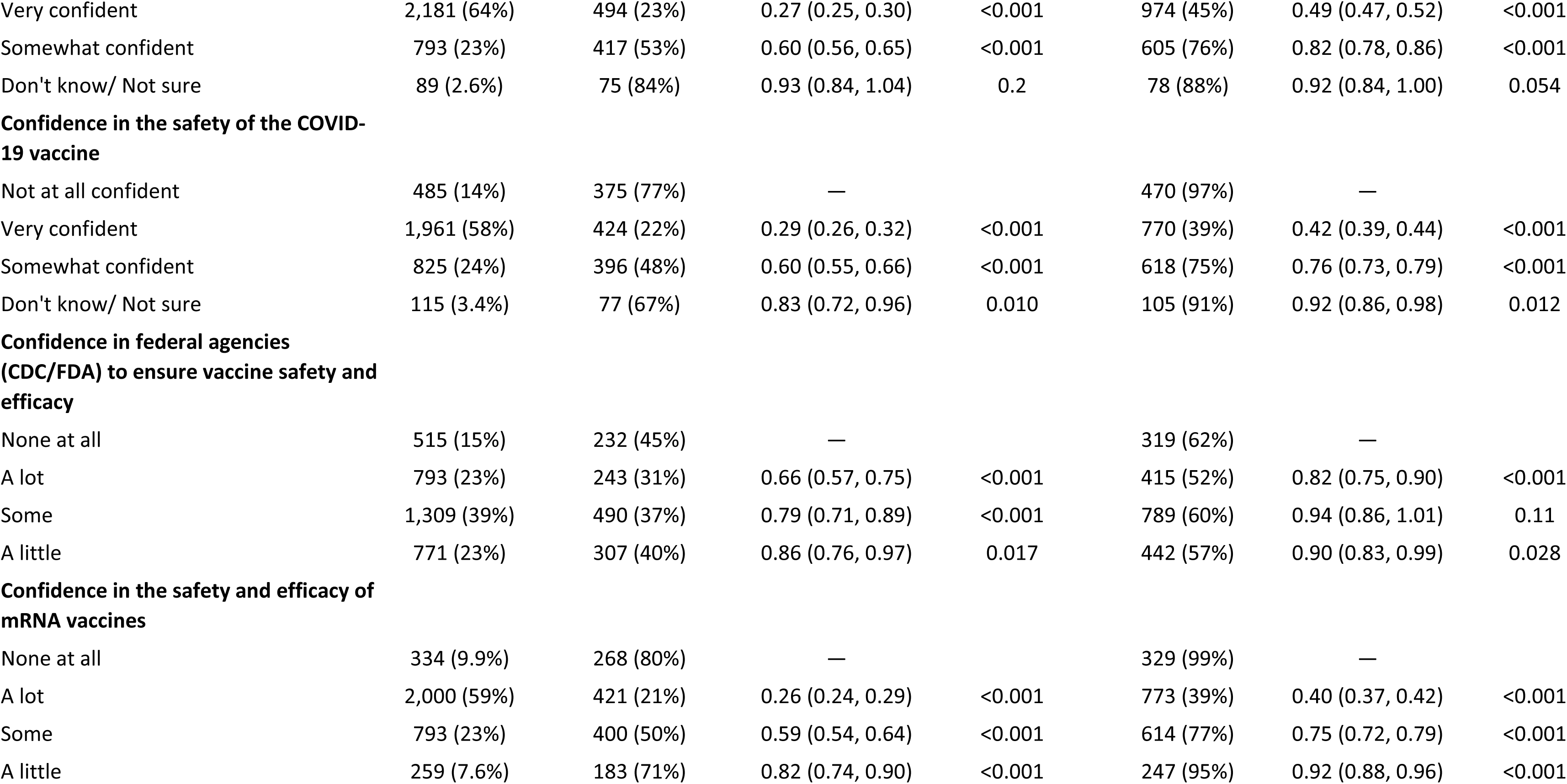
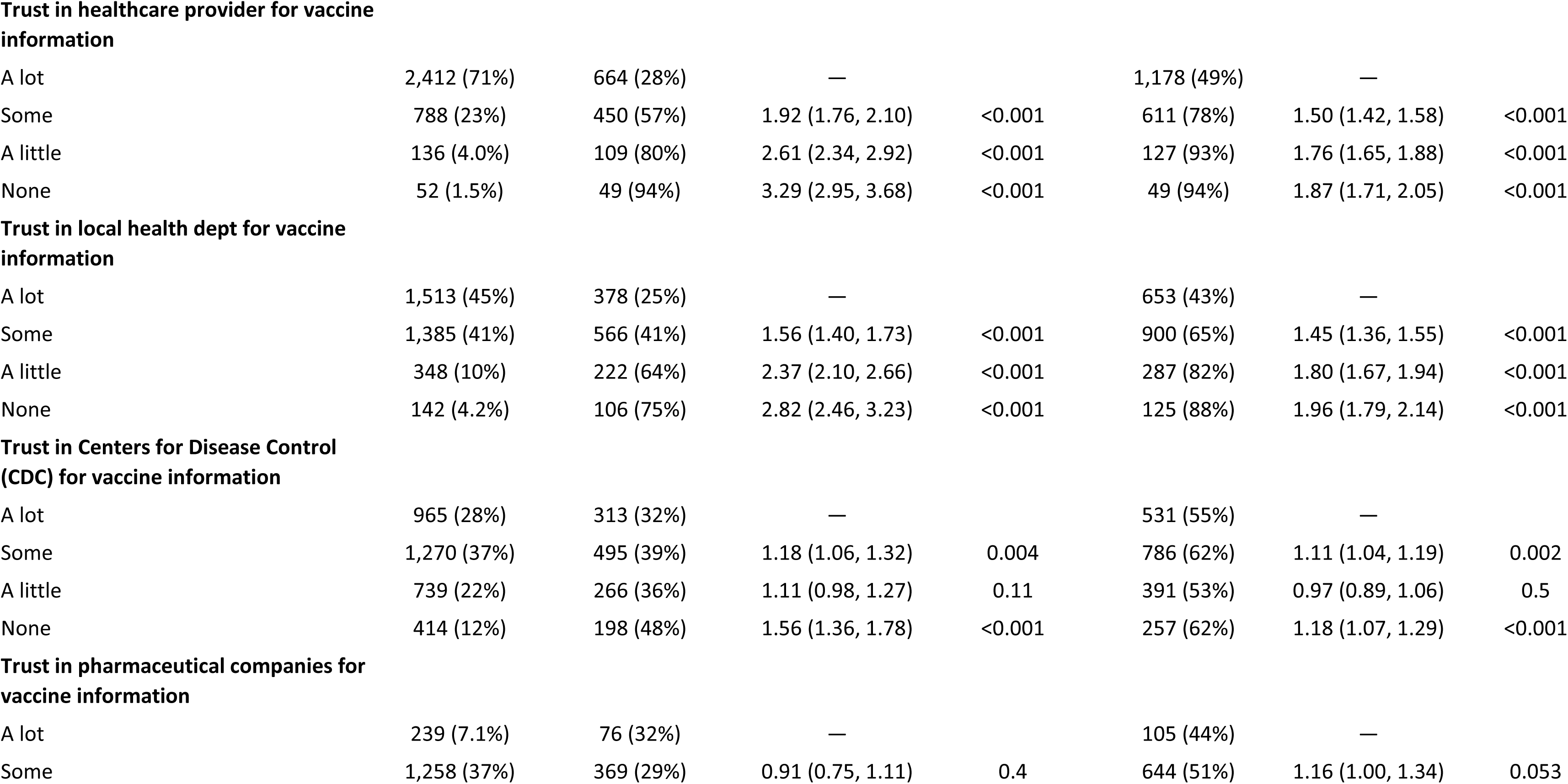
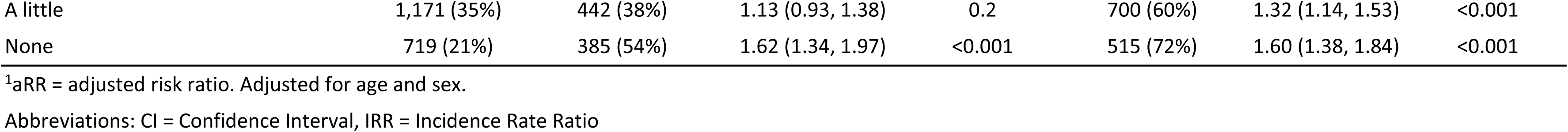
Predictors of influenza and COVID-19 non-vaccination during the Fall 2025/Winter 2026 respiratory virus season in the full analytic sample, CHASING COVID Cohort (N=3,390)

**Supplementary Table S2.**
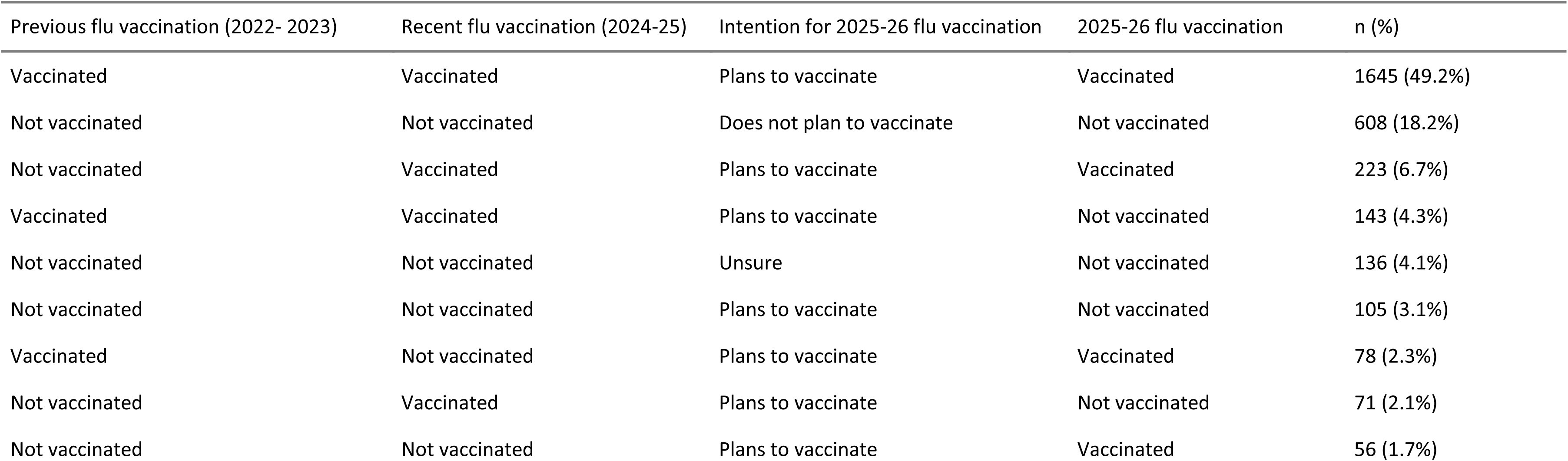
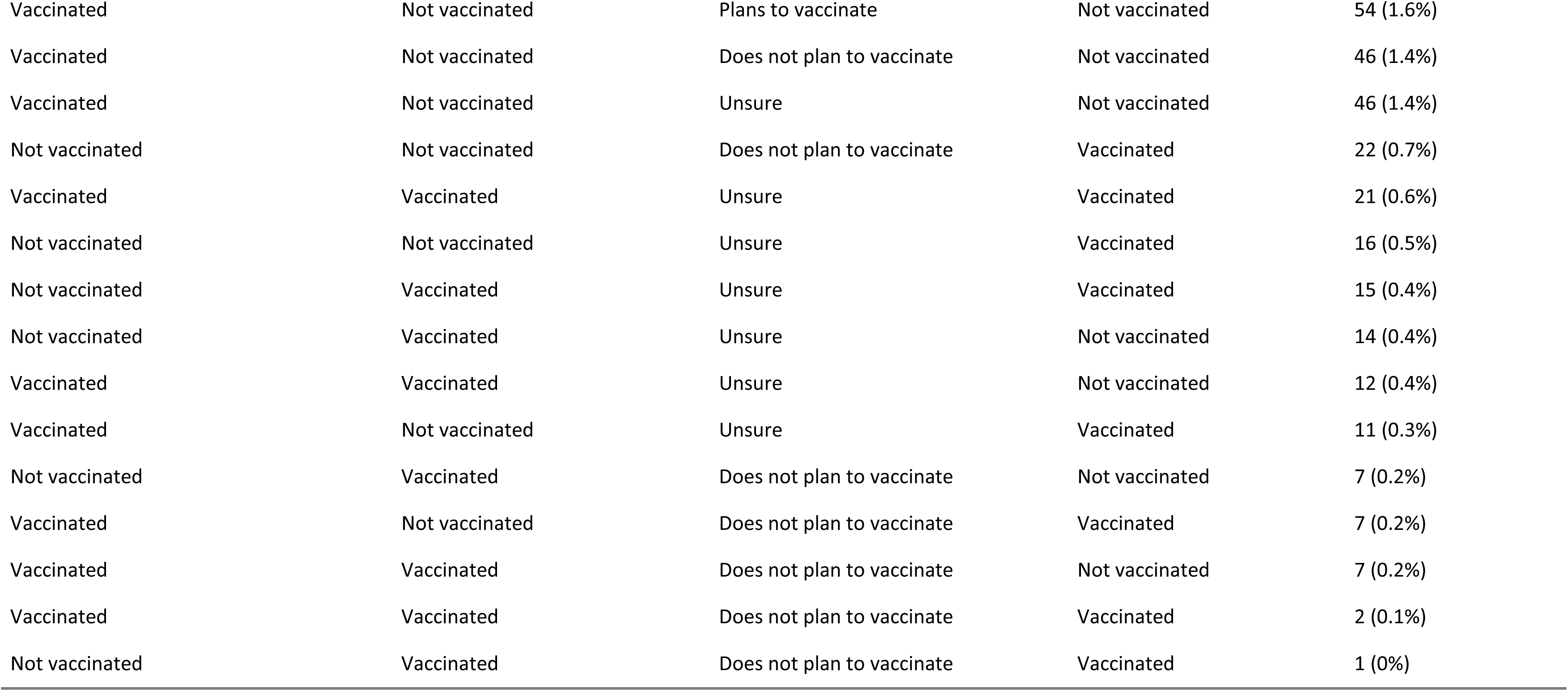
Influenza vaccination trajectories across prior vaccination, recent vaccination, intention, and 2025-26 behavior, CHASING COVID Cohort (N=3,346)

**Supplementary Table S3.**
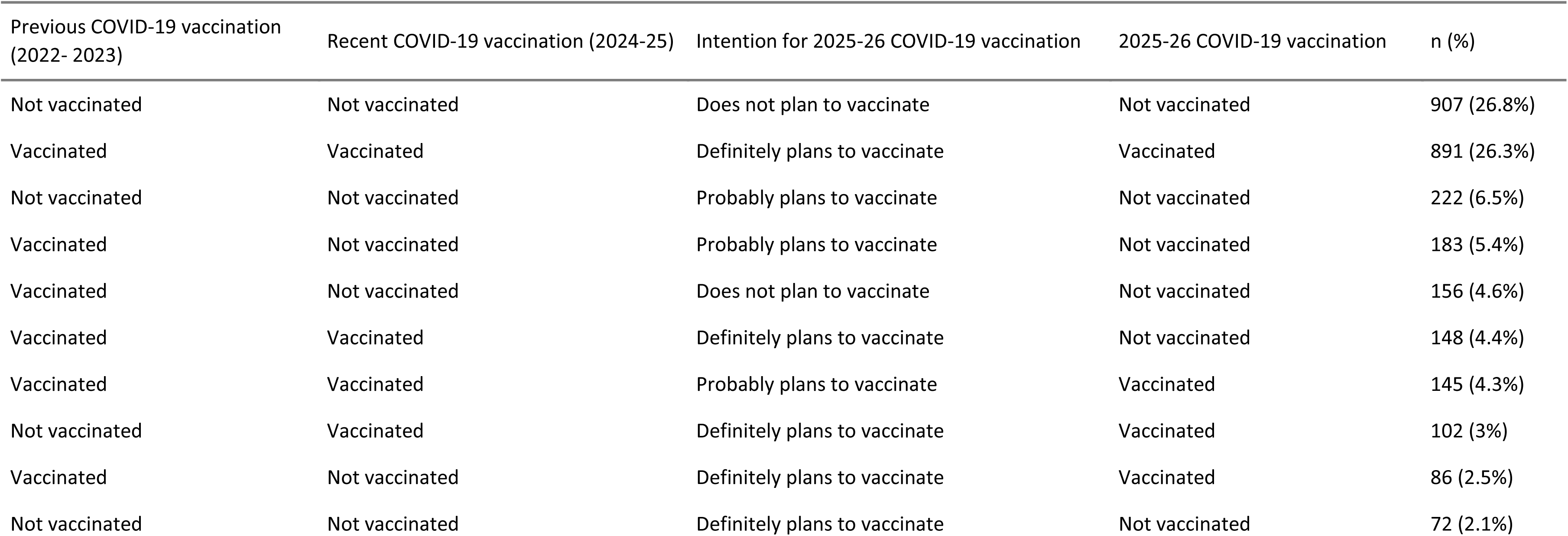
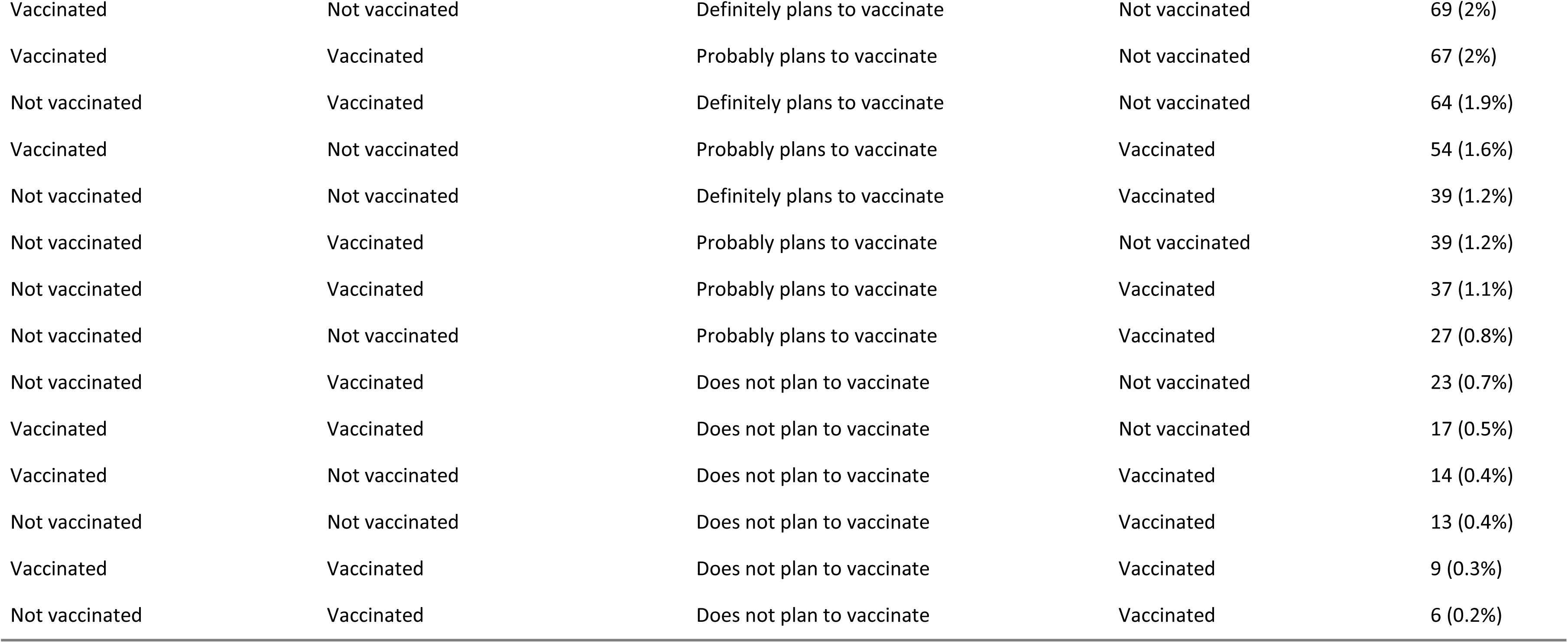
COVID-19 vaccination trajectories across prior vaccination, recent vaccination, intention, and 2025-26 behavior, CHASING COVID Cohort (N=3,390)

**Supplementary Figure S2.**
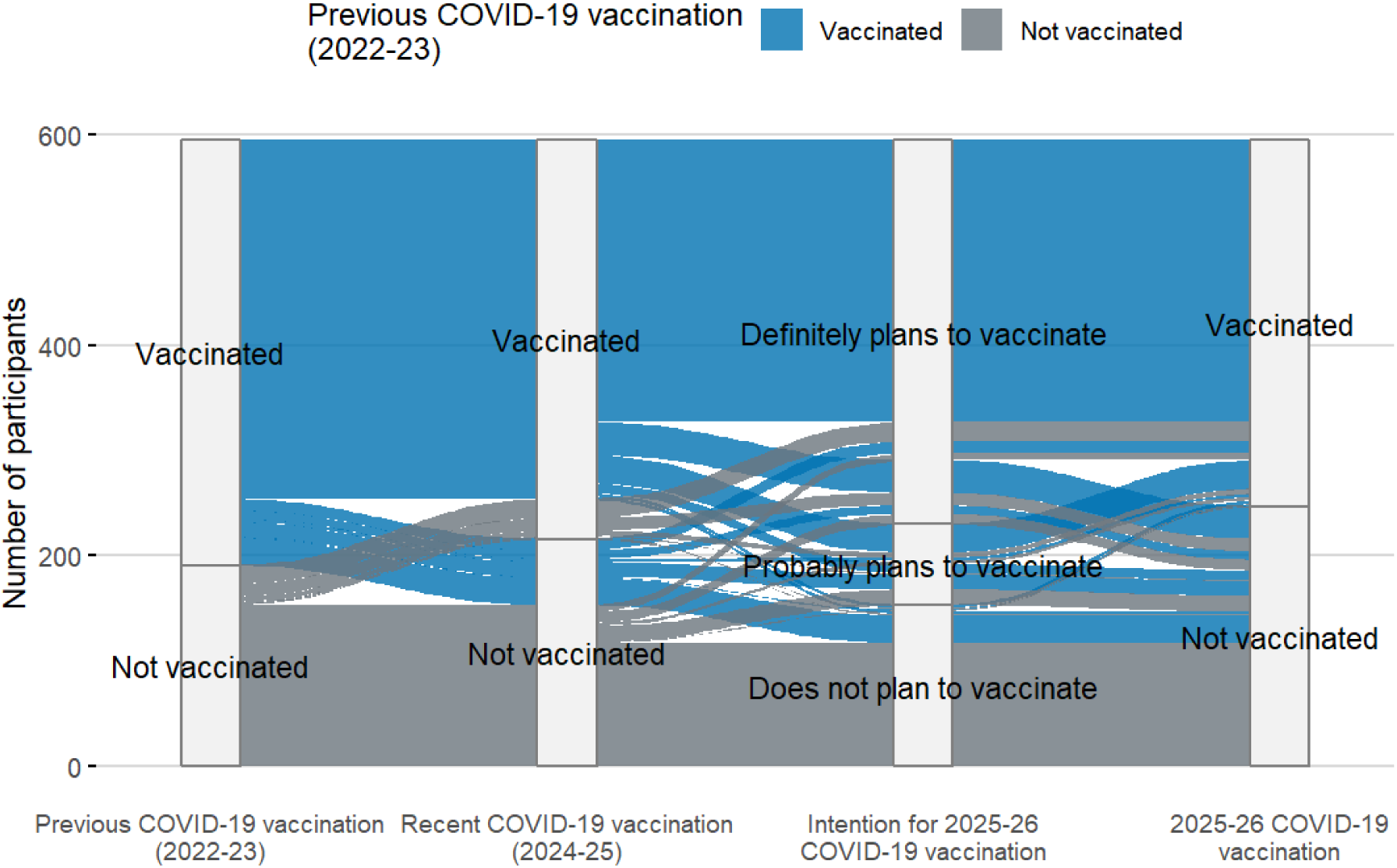
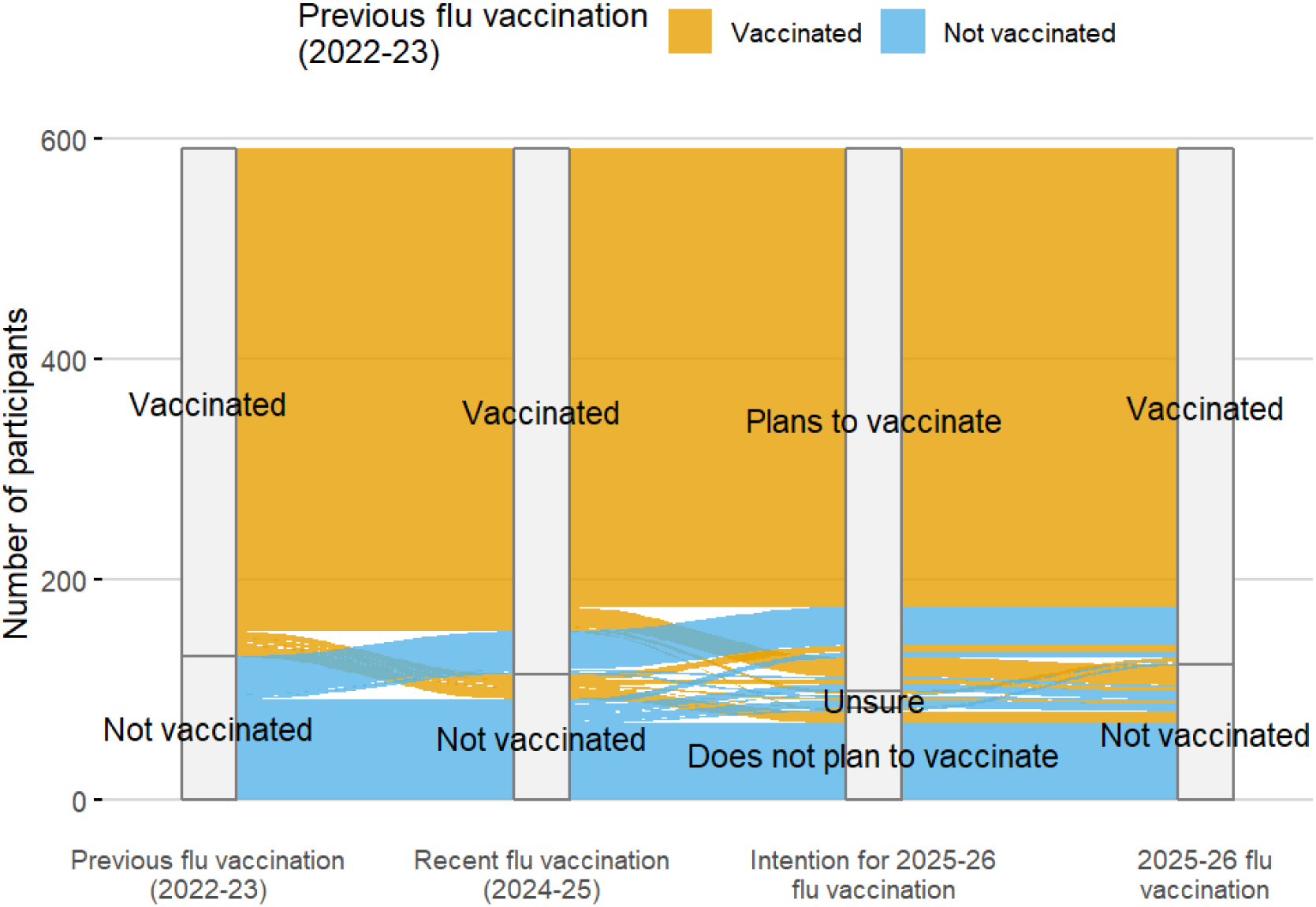
Participant trajectories across vaccination history, intention, and Fall 2025-Winter 2026 vaccination among adults aged ≥65 years, CHASING COVID Cohort (N=595)

**Supplementary Table S4.**
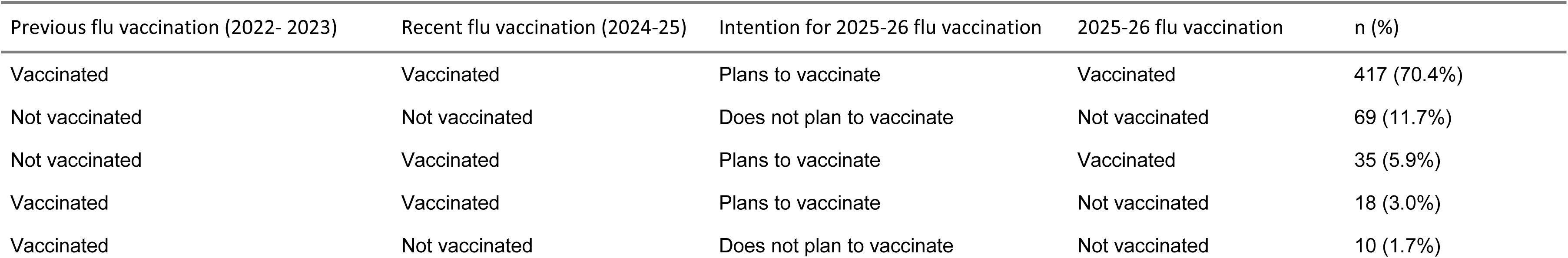
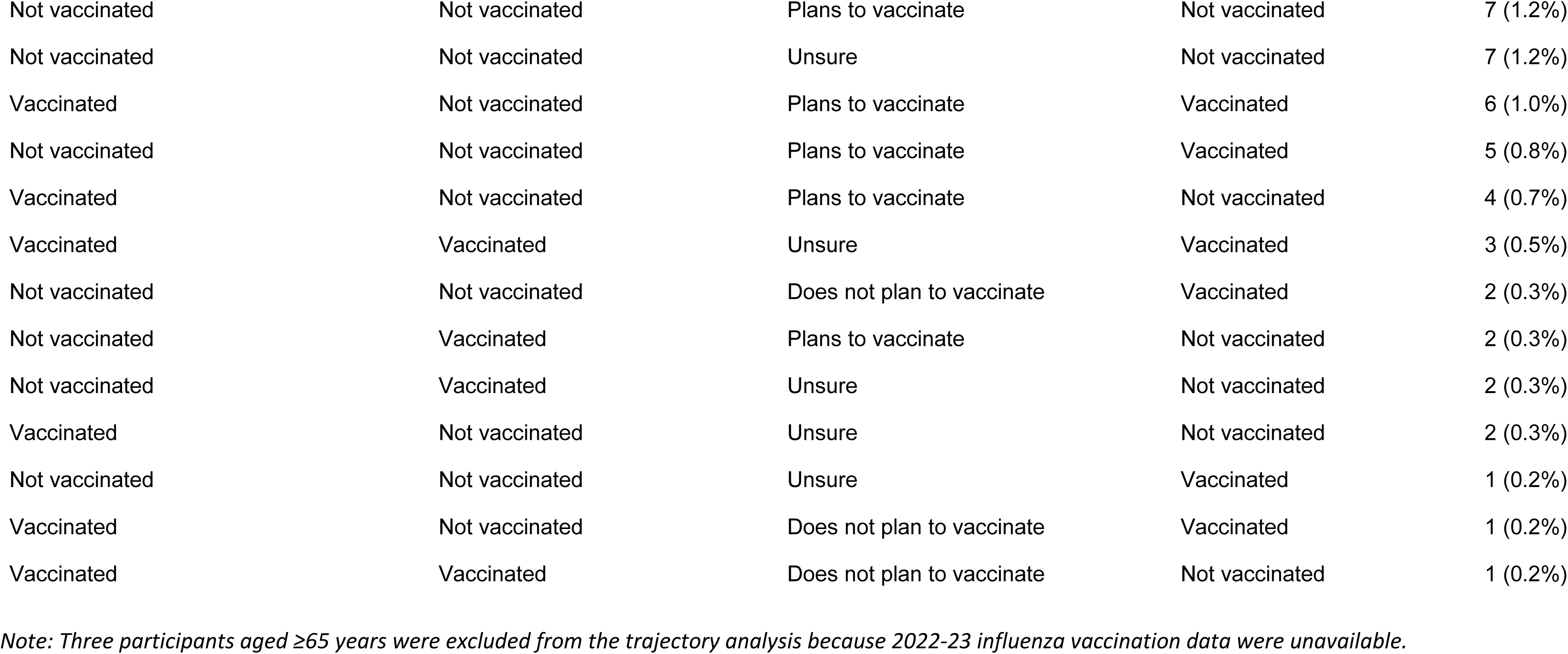
Influenza vaccination trajectories across prior vaccination, recent vaccination, intention, and 2025-26 behavior among adults aged ≥65 years, CHASING COVID Cohort (N=592)

**Supplementary Table S5.**
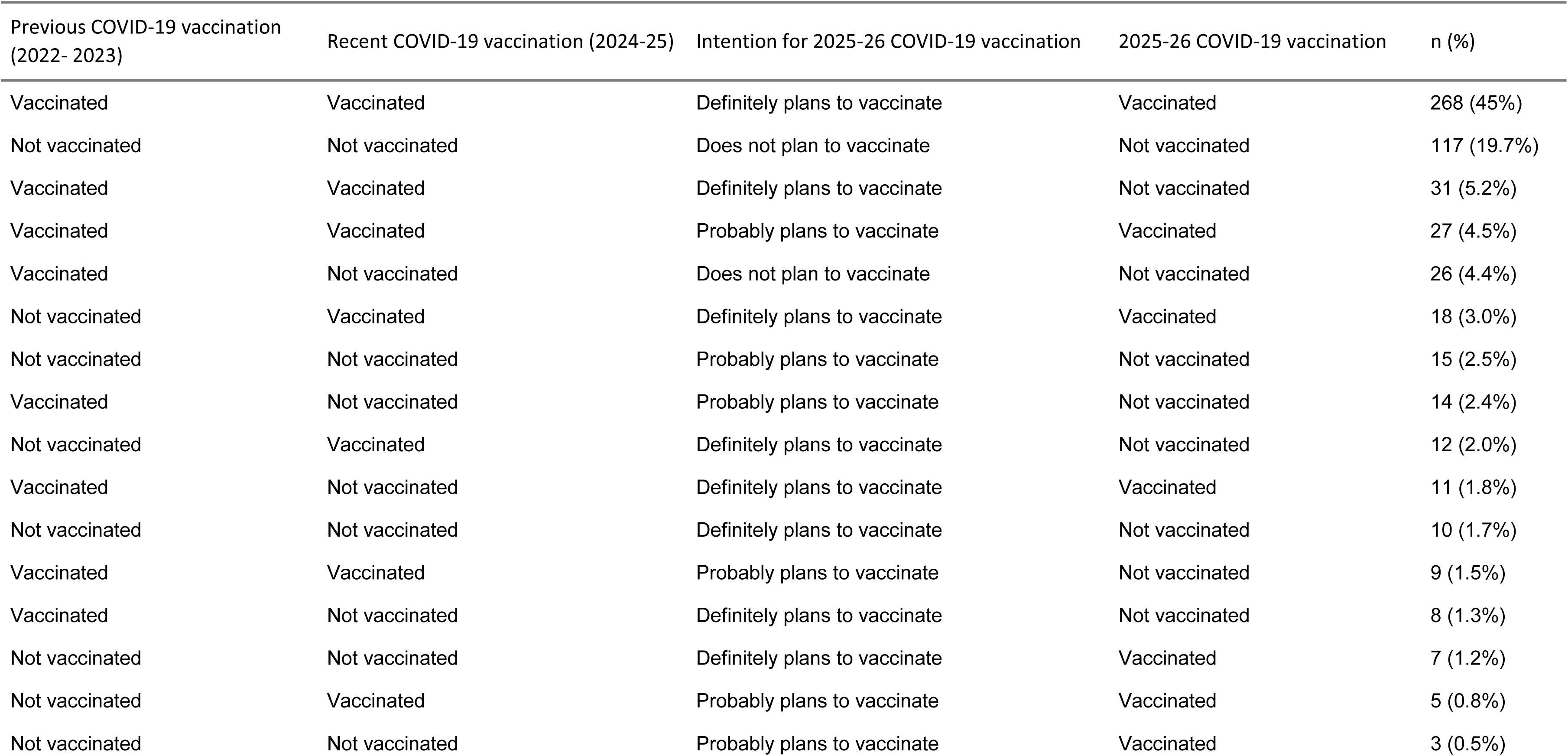
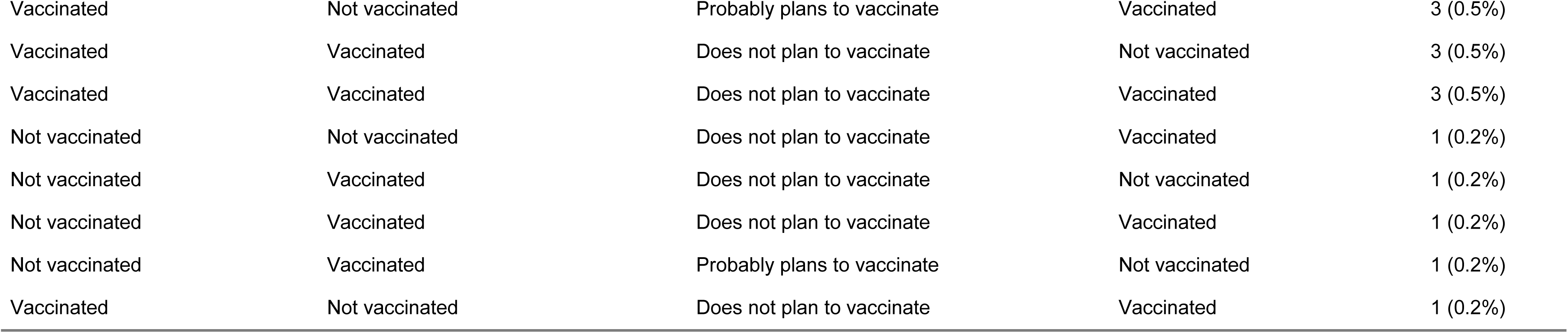
COVID-19 vaccination trajectories across prior vaccination, recent vaccination, intention, and 2025-26 behavior among adults aged ≥65 years, CHASING COVID Cohort (N=595)

## References

1. Debbag R, Rudin D, Ceddia F, Watkins J. The Impact of Vaccination on COVID-19, Influenza, and Respiratory Syncytial Virus-Related Outcomes: A Narrative Review. Infect Dis Ther. 2025;14(Suppl 1):63–97. doi:10.1007/s40121-024-01079-x

2. Qasmieh SA, Penrose K, Robertson MM, et al. COVID-19 and Influenza vaccine co-administration, and missed opportunities to increase COVID-19 vaccination coverage among U.S. adults, 2023-2024. medRxiv. Preprint posted online April 16, 2025:2025.04.14.25325480. doi:10.1101/2025.04.14.25325480

3. CDC. Flu Vaccination Coverage, United States, 2022–2023 Influenza Season. FluVaxView. December 18, 2025. Accessed May 13, 2026. https://www.cdc.gov/fluvaxview/coverage-by-season/2022-2023.html

4. Lu PJ, Zhou T, Santibanez TA, et al. COVID-19 Bivalent Booster Vaccination Coverage and Intent to Receive Booster Vaccination Among Adolescents and Adults - United States, November-December 2022. MMWR Morb Mortal Wkly Rep. 2023;72(7):190–198. doi:10.15585/mmwr.mm7207a5

5. Black CL, Kriss JL, Razzaghi H, et al. Influenza, Updated COVID-19, and Respiratory Syncytial Virus Vaccination Coverage Among Adults - United States, Fall 2023. MMWR Morb Mortal Wkly Rep. 2023;72(51):1377–1382. doi:10.15585/mmwr.mm7251a4

6. CDC. Vaccination Uptake, Intent, and Confidence. RespVaxView. March 11, 2026. Accessed May 13, 2026. https://www.cdc.gov/respvaxview/dashboards/vaccination-behavioral-social-drivers.html

7. Gómez E. Are we having the wrong conversation about vaccine hesitancy? Leading research projects weigh in with recommendations. RIVER EU. September 28, 2023. Accessed April 30, 2026. https://river-eu.org/are-we-having-the-wrong-conversation-about-vaccine-hesitancy-leading-research-projects-weigh-in-with-recommendations/

8. Wood S, Schulman K. When Vaccine Apathy, Not Hesitancy, Drives Vaccine Disinterest. JAMA. 2021;325(24):2435–2436. doi:10.1001/jama.2021.7707

9. Cummings PJ de-Winton, Bravo CG, Dukes KC, et al. Modifiable social and structural factors influence COVID-19 vaccine intention among frontline workers in the Midwestern USA: a community-engaged survey study. bmjph. 2025;3(1). doi:10.1136/bmjph-2023-000859

10. UCHealth KKM. Protect yourself from COVID-19 and flu this fall and winter. Everything you need to know about new flu shots and the updated 2025-26 COVID-19 vaccine. UCHealth Today. August 29, 2025. Accessed July 6, 2026. https://www.uchealth.org/today/everything-you-need-to-know-about-the-2025-26-covid-19-vaccine-and-flu-shots/

11. Richman AR, Schwartz AJ, Maness SB, Sanchez L, Torres E. Exploring Vaccine Hesitancy, Structural Barriers, and Trust in Vaccine Information Among Populations Living in the Rural Southern United States. Vaccines (Basel). 2025;13(7):699. doi:10.3390/vaccines13070699

12. Ortiz-Prado E, Suárez-Sangucho IA, Vasconez-Gonzalez J, et al. Pandemic paradox: How the COVID-19 crisis transformed vaccine hesitancy into a two-edged sword. Hum Vaccin Immunother. 21(1):2543167. doi:10.1080/21645515.2025.2543167

13. Lee SK, Sun J, Jang S, Connelly S. Misinformation of COVID-19 vaccines and vaccine hesitancy. Sci Rep. 2022;12(1):13681. doi:10.1038/s41598-022-17430-6

14. Neely SR, Eldredge C, Ersing R, Remington C. Vaccine Hesitancy and Exposure to Misinformation: a Survey Analysis. J Gen Intern Med. 2022;37(1):179–187. doi:10.1007/s11606-021-07171-z

15. Piltch-Loeb R, Penrose K, Stanton E, et al. Safety, Efficacy, and Ill Intent: Examining COVID-19 Vaccine Perceptions among the New Undervaccinated Moveable Middle in a U.S. Cohort, October 2022. Vaccines (Basel). 2023;11(11):1665. doi:10.3390/vaccines11111665

16. Public Health in 2025: Year in Review | Johns Hopkins | Bloomberg School of Public Health. December 17, 2025. Accessed April 30, 2026. https://publichealth.jhu.edu/2025/public-health-in-2025-year-in-review

17. One Big Beautiful Bill Law Summary. Accessed April 30, 2026. https://www.astho.org/advocacy/federal-government-affairs/leg-alerts/2025/one-big-beautiful-bill-law-summary/

18. Simmons-Duffin S. The Trump administration restructures federal health agencies, cuts 20,000 jobs. NPR. March 27, 2025. Accessed April 30, 2026. https://www.npr.org/sections/shots-health-news/2025/03/27/nx-s1-5342414/hhs-doge-rif-rfk-job-cuts

19. Public Health Infrastructure in Crisis: HHS Workforce Cuts, Reorganizations, and Funding Reductions: Impacts and Solutions. TFAH. Accessed April 30, 2026. https://www.tfah.org/report-details/funding-report-2025/

20. IDSA. More Than 130 Organizations Express Concern Over Changes to CDC’s Advisory Committee on Immunization Practices Charter. Accessed April 30, 2026. https://www.idsociety.org/news--publications-new/articles/2026/more-than-130-organizations-express-concern-over-changes-to-cdcs-advisory-committee-on-immunization-practices-charter/

21. ccaplan7. Upheaval at ACIP: A Timeline of U.S. Vaccine Policy and Leadership Shifts. PFID/VacciNATION. March 25, 2026. Accessed April 30, 2026. https://www.fightinfectiousdisease.org/post/upheaval-at-acip-a-timeline-of-u-s-vaccine-policy-and-leadership-shifts

22. Prasad V, Makary MA. An Evidence-Based Approach to Covid-19 Vaccination. New England Journal of Medicine. 2025;392(24):2484–2486. doi:10.1056/NEJMsb2506929

23. Stein R. A stricter FDA policy for COVID vaccines could limit future access. NPR. May 20, 2025. Accessed April 30, 2026. https://www.npr.org/sections/shots-health-news/2025/05/20/nx-s1-5405013/fda-covid-vaccine-limits

24. Mandavilli A. C.D.C. Cancels Publication of Study Showing Benefits of Covid Vaccines. The New York Times. April 22, 2026. Accessed April 30, 2026. https://www.nytimes.com/2026/04/22/us/politics/cdc-covid-vaccine-study.html

25. A CDC Website Change May Contribute to the Public’s Uncertainty on Vaccines and Autism. KFF. Accessed April 30, 2026. https://www.kff.org/quick-take/a-cdc-website-change-may-contribute-to-the-publics-uncertainty-on-vaccines-and-autism/

26. Sept. 26, 2025: National Advocacy Update. American Medical Association. September 26, 2025. Accessed July 6, 2026. https://www.ama-assn.org/health-care-advocacy/advocacy-update/sept-26-2025-national-advocacy-update

27. Carter SJ, Lauderdale J, Stollings JL, et al. Factors Influencing Influenza and COVID-19 Vaccine Decision-Making in the Post-ICU Period. CHEST Crit Care. 2023;1(3):100027. doi:10.1016/j.chstcc.2023.100027

28. Robertson MM, Kulkarni SG, Rane M, et al. Cohort profile: a national, community-based prospective cohort study of SARS-CoV-2 pandemic outcomes in the USA-the CHASING COVID Cohort study. BMJ Open. 2021;11(9):e048778. doi:10.1136/bmjopen-2021-048778

29. CHASING COVID Cohort Study. CUNY ISPH. Accessed March 11, 2024. https://cunyisph.org/chasing-covid/

30. CDC. People at Increased Risk for Severe Respiratory Illnesses. Respiratory Illnesses. August 14, 2025. Accessed May 15, 2026. https://www.cdc.gov/respiratory-viruses/risk-factors/index.html

31. SteelFisher GK, Findling MG, Caporello HL, McGowan E, Espino L, Sutton J. Divergent Attitudes Toward COVID-19 Vaccine vs Influenza Vaccine. JAMA Netw Open. 2023;6(12):e2349881. doi:10.1001/jamanetworkopen.2023.49881

32. Yuan J, Dong M, Ip DKM, So HC, Liao Q. Dual decision-making routes for COVID-19 and influenza vaccines uptake in parents: A mixed-methods study. Br J Health Psychol. 2025;30(2):e12789. doi:10.1111/bjhp.12789

33. CDC. Flu and People 65 Years and Older. Influenza (Flu). February 14, 2025. Accessed June 15, 2026. https://www.cdc.gov/flu/highrisk/65over.htm

34. CDC. Staying Up to Date with COVID-19 Vaccines. Covid. November 21, 2025. Accessed June 15, 2026. https://www.cdc.gov/covid/vaccines/stay-up-to-date.html

35. Lin C, Mullen J, Smith D, Kotarba M, Kaplan SJ, Tu P. Healthcare Providers’ Vaccine Perceptions, Hesitancy, and Recommendation to Patients: A Systematic Review. Vaccines. 2021;9(7):713. doi:10.3390/vaccines9070713

36. McElfish PA, Selig JP, Scott AJ, et al. Associations Between General Vaccine Hesitancy and Healthcare Access Among Arkansans. J Gen Intern Med. 2023;38(4):841–847. doi:10.1007/s11606-022-07859-w

37. Zha P, Qureshi R, Mahat G, Gao L, Garcia C, Wei Z. Trust in Information Sources and COVID-19 Vaccine Uptake. Public Health Chall. 2025;4(4):e70145. doi:10.1002/puh2.70145

38. Nguyen KH, Yankey D, Lu P jun, et al. Report of Health Care Provider Recommendation for COVID-19 Vaccination Among Adults, by Recipient COVID-19 Vaccination Status and Attitudes — United States, April–September 2021. MMWR Morb Mortal Wkly Rep. 2021;70(50):1723–1730. doi:10.15585/mmwr.mm7050a1

39. Kolobova I, Nyaku MK, Karakusevic A, Bridge D, Fotheringham I, O’Brien M. Vaccine uptake and barriers to vaccination among at-risk adult populations in the US. Hum Vaccin Immunother. 18(5):2055422. doi:10.1080/21645515.2022.2055422

40. Robinson R, Nguyen E, Wright M, et al. Factors contributing to vaccine hesitancy and reduced vaccine confidence in rural underserved populations. Humanit Soc Sci Commun. 2022;9(1):416. doi:10.1057/s41599-022-01439-3

41. Malik AA, Ahmed N, Shafiq M, et al. Behavioral interventions for vaccination uptake: A systematic review and meta-analysis. Health Policy. 2023;137:104894. doi:10.1016/j.healthpol.2023.104894

42. Shen Y, Wang J, zhao Q, et al. Predicting future vaccination habits: The link between influenza vaccination patterns and future vaccination decisions among old aged adults in China. Journal of Infection and Public Health. 2024;17(6):1079–1085. doi:10.1016/j.jiph.2024.04.017

43. Mahameed H, Al-Mahzoum K, AlRaie LA, et al. Previous Vaccination History and Psychological Factors as Significant Predictors of Willingness to Receive Mpox Vaccination and a Favorable Attitude towards Compulsory Vaccination. Vaccines (Basel). 2023;11(5):897. doi:10.3390/vaccines11050897

44. Walsh MM, Parker AM, Vardavas R, Nowak SA, Kennedy DP, Gidengil CA. The Stability of Influenza Vaccination Behavior Over Time: A Longitudinal Analysis of Individuals Across 8 Years. Ann Behav Med. 2020;54(10):783–793. doi:10.1093/abm/kaaa017

45. Mirpuri P, Rovin RA. COVID-19 and Historic Influenza Vaccinations in the United States: A Comparative Analysis. Vaccines (Basel). 2021;9(11):1284. doi:10.3390/vaccines9111284

46. Fuller HR, Huseth-Zosel A, Van Vleet B, Carson PJ. Barriers to vaccination among older adults: Demographic variation and links to vaccine acceptance. Aging and Health Research. 2024;4(1):100176. doi:10.1016/j.ahr.2023.100176

47. CDC. Vaccination Coverage among Adults in the United States, National Health Interview Survey, 2022. AdultVaxView. April 15, 2025. Accessed June 11, 2026. https://www.cdc.gov/adultvaxview/publications-resources/adult-vaccination-coverage-2022.html

48. Miller NS. Racial disparities in uptake rates for recommended adult vaccines. The Journalist’s Resource. September 22, 2021. Accessed June 11, 2026. https://live-journalists-resource.pantheonsite.io/home/racial-disparities-adult-vaccines-rates/

49. Kim D. Associations of race/ethnicity and socioeconomic factors with vaccination among US adults during the COVID-19 pandemic, January to March 2021. Prev Med Rep. 2022;31:102021. doi:10.1016/j.pmedr.2022.102021

50. Galagali PM, Kinikar AA, Kumar VS. Vaccine Hesitancy: Obstacles and Challenges. Curr Pediatr Rep. 2022;10(4):241–248. doi:10.1007/s40124-022-00278-9

51. Welch VL, Metcalf T, Macey R, et al. Understanding the Barriers and Attitudes toward Influenza Vaccine Uptake in the Adult General Population: A Rapid Review. Vaccines (Basel). 2023;11(1):180. doi:10.3390/vaccines11010180

52. CDC. Strategies for Increasing Adult Vaccination Rates. Vaccine Information for Adults. December 17, 2025. Accessed July 7, 2026. https://www.cdc.gov/vaccines-adults/hcp/vaccination-guidelines/index.html

53. Jaca A, Mathebula L, Malinga T, et al. Interventions to Improve Vaccination Uptake Among Adults: A Systematic Review and Meta-Analysis. Vaccines (Basel). 2025;13(8):811. doi:10.3390/vaccines13080811

54. Burson RC, Buttenheim AM, Armstrong A, Feemster KA. Community pharmacies as sites of adult vaccination: A systematic review. Hum Vaccin Immunother. 2016;12(12):3146–3159. doi:10.1080/21645515.2016.1215393

