## Supplementary Material for "Gaps Between Willingness and Uptake of Influenza and COVID-19 Vaccines During the 2025-26 Respiratory Virus Season in a U.S. Adult Cohort"

**SUPPLEMENTAL MATERIALS**

### **Supplemental Table S1. Vaccine-Related Survey Measures, Response Options, and Variable Definitions**

| Construct | Survey Measure | Response Options | Coding / Categorization |
| --- | --- | --- | --- |
| Employment status | Participants were asked about their current employment status.^*^ | Employed for wages; Self-employed; Out of work ≥1 year; Out of work <1 year; Homemaker; Student; Retired; Unable to work | Categorized according to participant response. |
| Parent/guardian status | Participants were asked whether they were the parent or guardian of a child under age 18. | Yes; No | Participants reporting one or more children under age 18 were classified as parents/guardians. |
| Pregnancy status | Participants assigned female sex at birth were asked whether they were currently pregnant.^*^ | Yes; No; Don’t know/Not sure | Categorized according to participant response. |
| Relationship status | Participants were asked about their current marital or relationship status.^*^ | Married; Divorced; Widowed; Separated; Never married; Member of an unmarried couple; Don’t know/Not sure | Categorized according to participant response. |
| Annual household income | Participants were asked to report annual household income from all sources.^*^ | Less than $10,000?; $10,000 to less than $15,000; $15,000 to less than $20,000;  $20,000 to less than $25,000; $25,000 to less than $35,000; $35,000 to less than $50,000; $50,000 to less than $75,000; $75,000 to less than $100,000; $100,000 to less than $150,000; $150,000 to less than $200,000; $200,000 or more | Less than $25,000; $25,000 to less than $50,000; $50,000 to $75,000; $75000 to $100,000; $100,000 to less than $200,000; $200,000 or more |
| Food insecurity | Food insecurity was measured using a validated two-item screener assessing concerns about food running out and food affordability during the prior 12 months.^1,2^ | Often true; Sometimes true; Never true | Participants were classified as food insecure if they responded “often true” or “sometimes true” to either item. |
| Housing insecurity | Housing insecurity was assessed by asking whether participants had been unable to pay rent, mortgage, or utility bills during the prior 12 months.^*^ | Yes; No; Don’t know/Not sure | Participants responding “yes” were classified as housing insecure. |
| Disability status | Disability status was assessed using the six-item American Community Survey disability measure evaluating hearing, vision, cognition, mobility, self-care, and independent living difficulties. | Yes; No | Participants endorsing any disability item were classified as having a disability. |
| Underlying health conditions | Participants were asked whether a healthcare professional had ever diagnosed them with selected chronic conditions identified by the CDC in March 2020 as increasing risk for severe COVID-19. | Yes; No; Don’t know/Not sure | Conditions included chronic obstructive pulmonary disease (COPD), emphysema, chronic bronchitis, angina/coronary heart disease, high blood pressure, history of myocardial infarction, current asthma, type 2 diabetes, kidney disease, immunocompromising condition, and HIV-positive status. Participants reporting one or more conditions were classified as having an underlying health condition. |
| Health insurance coverage | Participants reported their current source of health insurance coverage.^*^ | Employer-sponsored; Private; Medicare; Medicaid; Military-related; No coverage; Other | Participants reporting no coverage were classified as uninsured. |
| Regular healthcare provider | Participants were asked whether they had one or more personal healthcare providers.^*^ | Yes, one; More than one; No; Don’t know/Not sure | Participants reporting “no” were classified as lacking a regular provider. |
| Cost-related barriers to care | Participants were asked whether there was a time in the prior 12 months when they needed to see a doctor but could not because of cost.^*^ | Yes; No; Don’t know/Not sure | Participants responding “yes” were classified as having a cost-related healthcare barrier. |
| Composite healthcare access barriers | Composite measure of barriers to healthcare access. Similar composite measures have been used previously to identify individuals with reduced access to healthcare services.^3^ | Derived variable | Participants were classified as having a healthcare access barrier if they lacked health insurance, lacked a regular healthcare provider, or reported being unable to see a doctor due to cost. |
| Trusted sources of vaccine information | Participants rated their level of trust in various individuals and organizations to provide reliable information about vaccines, including healthcare providers, local health departments, the Centers for Disease Control and Prevention (CDC), and pharmaceutical companies.^**^ | A lot; Some; A little; None at all | Categorized according to participant response. |
| Vaccine hesitancy | Vaccine hesitancy was assessed using the 10-item Vaccine Hesitancy Scale (VHS), which measures attitudes regarding vaccine importance, effectiveness, community benefit, and safety concerns. Example items included: “Childhood vaccines are important for children’s health,” “Childhood vaccines are effective,” “Having children vaccinated is important for the health of others in my community,” “I am concerned about serious adverse effects of vaccines,” and “Children do not need vaccines for diseases that are not common anymore.” | Strongly disagree (1); Disagree (2); Neither agree nor disagree (3); Agree (4); Strongly agree (5) | Items reflecting lower hesitancy were reverse-coded such that higher scores consistently reflected greater vaccine hesitancy. Item scores were summed to create a total score ranging from 10–50. Participants were categorized as having low (≤25), medium (26–43), or high (≥44) vaccine hesitancy. |
| Confidence in influenza vaccine safety | Participants reported their confidence in the safety of influenza vaccines.^**^ | Very confident; Somewhat confident; Not at all confident; Don’t know/Not sure | Categorized according to participant response. |
| Confidence in COVID-19 vaccine safety | Participants reported their confidence in the safety of COVID-19 vaccines.^**^ | Very confident; Somewhat confident; Not at all confident; Don’t know/Not sure | Categorized according to participant response. |
| Confidence in federal agencies | Participants reported how much confidence they had in federal agencies to ensure vaccine safety and effectiveness.^**^ | A lot; Some; A little; None at all | Categorized according to participant response. |
| Confidence in mRNA vaccines | Participants reported how much confidence they had in the safety and effectiveness of mRNA vaccines, such as the Moderna or Pfizer COVID-19 vaccines. ^**^ | A lot; Some; A little; None at all | Categorized according to participant response. |
| Influenza vaccination intentions | Participants were asked whether they planned to receive a flu vaccine during the upcoming 2025-2026 respiratory virus season. | Yes; No; Don’t know/Not sure | Participants responding “yes” were classified as intending to vaccinate. |
| COVID-19 vaccination intentions | Participants were asked whether, if an updated COVID-19 vaccine were available to them that fall, they would “definitely get it,” “probably get it,” “probably not get it,” or “definitely not get it.” ^*^ | Definitely get it; Probably get it; Probably not get it; Definitely not get it | Participants responding “definitely get it” were classified as intending to vaccinate. |
| Recent influenza vaccination | Participants were asked whether they had received either an intranasal influenza vaccine or injectable influenza vaccine during the previous 12 months. ^*^ | Yes; No; Don’t know/Not sure | Participants responding “yes” were classified as recently vaccinated. |
| Recent COVID-19 vaccination | Participants were asked whether they had received a COVID-19 vaccine dose during the previous 12 months. | Yes; No; Don’t know/Not sure | Participants responding “yes” were classified as recently vaccinated. |
| Influenza vaccination follow-through | Participants were asked in March 2026 whether they had received a flu vaccine (nasal spray or injection) since September 1, 2025. | Yes; No; Don’t know/Not sure | Participants responding “yes” were classified as vaccinated during the follow-up period. |
| COVID-19 vaccination follow-through | Participants were asked in March 2026 whether they had received a COVID-19 vaccine since September 1, 2025. | Yes; No; Don’t know/Not sure | Participants responding “yes” were classified as vaccinated during the follow-up period. |
| Reasons for non-vaccination | Participants who remained unvaccinated during follow-up were asked to select reasons for delaying or skipping influenza or COVID-19 vaccination. | Concern about side effects; Lack of trust in vaccine safety; Perceived lack of need; Health condition; Lack of healthcare provider recommendation; Inability to find a convenient time/place/appointment; Cost or insurance barriers; Belief that they could stay healthy in other ways; Other (write-in); None of the above | Participants could select multiple responses. Open-ended “other” responses were qualitatively reviewed and grouped into broad thematic categories. |

*^1^Hager ER, Quigg AM, Black MM, et al. Development and validity of a 2-item screen to identify families at risk for food insecurity. Pediatrics. 2010;126(1):e26-32. doi:10.1542/peds.2009-3146*

*^2^Radandt NE, Corbridge T, Johnson DB, Kim AS, Scott JM, Coldwell SE. Validation of a Two-Item Food Security Screening Tool in a Dental Setting. J Dent Child (Chic). 2018;85(3):114-119.*

*^3^Robertson MM, Shamsunder MG, Brazier E, et al. Racial/Ethnic Disparities in Exposure, Disease Susceptibility, and Clinical Outcomes during COVID-19 Pandemic in National Cohort of Adults, United States. Emerg Infect Dis. 2022;28(11):2171-2180. doi:10.3201/eid2811.220072*

**Question source from the 2023 Behavioral Risk Factor Surveillance System questionnaire*

***Question sourced from the KFF Health Information and Tracking Poll*

**Supplemental Table S2. Predictors of influenza and COVID-19 non-vaccination during the Fall 2025/Winter 2026 respiratory virus season in the full analytic sample, CHASING COVID Cohort (N=3,390)**

| **Characteristic**^1^ | | **Influenza non-vaccination** | | | **COVID-19 non-vaccination** | | |
| --- | --- | --- | --- | --- | --- | --- | --- |
|  | **Total N (%)**^1^ | **N (row %)**^1^ | **aRR (95% CI)**^1^ | **p-value**^1^ | **N (row %)**^1^ | **aRR (95% CI)**^1^ | **p-value**^1^ |
| Total | 3390 (100%) | 1272 (38%) |  |  | 1967 (58%) |  |  |
| **Age** |  |  |  |  |  |  |  |
| 18–29 | 365 (11%) | 167 (46%) | — |  | 252 (69%) | — |  |
| 30–39 | 878 (26%) | 387 (44%) | 0.99 (0.86, 1.13) | 0.9 | 573 (65%) | 0.96 (0.88, 1.04) | 0.4 |
| 40–49 | 778 (23%) | 330 (42%) | 0.97 (0.85, 1.12) | 0.7 | 478 (61%) | 0.92 (0.84, 1.00) | 0.058 |
| 50–59 | 541 (16%) | 201 (37%) | 0.84 (0.72, 0.98) | 0.031 | 311 (57%) | 0.85 (0.77, 0.94) | 0.001 |
| 60+ | 828 (24%) | 187 (23%) | 0.50 (0.42, 0.59) | <0.001 | 353 (43%) | 0.62 (0.56, 0.69) | <0.001 |
| **Gender** |  |  |  |  |  |  |  |
| Female | 1,848 (55%) | 773 (42%) | — |  | 1,175 (64%) | — |  |
| Male | 1,447 (43%) | 471 (33%) | 0.77 (0.70, 0.85) | <0.001 | 753 (52%) | 0.82 (0.77, 0.87) | <0.001 |
| Non-binary | 95 (2.8%) | 28 (29%) | 0.61 (0.45, 0.84) | 0.003 | 39 (41%) | 0.58 (0.46, 0.75) | <0.001 |
| **Race/ethnicity** |  |  |  |  |  |  |  |
| White NH | 2,116 (62%) | 677 (32%) | — |  | 1,125 (53%) | — |  |
| Hispanic | 553 (16%) | 271 (49%) | 1.35 (1.22, 1.50) | <0.001 | 368 (67%) | 1.15 (1.07, 1.23) | <0.001 |
| Black NH | 350 (10%) | 184 (53%) | 1.49 (1.33, 1.67) | <0.001 | 241 (69%) | 1.21 (1.12, 1.31) | <0.001 |
| Asian/Pacific Islander | 259 (7.6%) | 80 (31%) | 0.83 (0.69, 1.01) | 0.069 | 159 (61%) | 1.04 (0.93, 1.15) | 0.5 |
| Other/Multiracial | 110 (3.2%) | 59 (54%) | 1.59 (1.33, 1.92) | <0.001 | 73 (66%) | 1.22 (1.07, 1.40) | 0.004 |
| **Education** |  |  |  |  |  |  |  |
| HS graduate or less | 376 (11%) | 226 (60%) | — |  | 299 (80%) | — |  |
| Some college | 888 (26%) | 444 (50%) | 0.88 (0.79, 0.97) | 0.011 | 623 (70%) | 0.92 (0.86, 0.98) | 0.011 |
| College graduate | 2,126 (63%) | 602 (28%) | 0.50 (0.45, 0.56) | <0.001 | 1,045 (49%) | 0.65 (0.61, 0.70) | <0.001 |
| **Annual Household Income** |  |  |  |  |  |  |  |
| <$25,000 | 425 (13%) | 232 (55%) | — |  | 311 (73%) | — |  |
| $25,000-$49,999 | 548 (16%) | 275 (50%) | 0.95 (0.84, 1.06) | 0.4 | 385 (70%) | 0.98 (0.90, 1.06) | 0.6 |
| $50,000-$74,999 | 548 (16%) | 234 (43%) | 0.78 (0.69, 0.88) | <0.001 | 333 (61%) | 0.83 (0.76, 0.90) | <0.001 |
| $75,000-$99,999 | 499 (15%) | 181 (36%) | 0.68 (0.59, 0.79) | <0.001 | 284 (57%) | 0.79 (0.72, 0.87) | <0.001 |
| $100,000-$199,999 | 932 (28%) | 267 (29%) | 0.52 (0.45, 0.59) | <0.001 | 484 (52%) | 0.70 (0.65, 0.77) | <0.001 |
| $200,000+ | 428 (13%) | 80 (19%) | 0.34 (0.27, 0.42) | <0.001 | 167 (39%) | 0.53 (0.47, 0.61) | <0.001 |
| **Employment status** |  |  |  |  |  |  |  |
| Employed | 2,356 (69%) | 911 (39%) | — |  | 1,415 (60%) | — |  |
| Unemployed | 178 (5.3%) | 81 (46%) | 1.20 (1.02, 1.42) | 0.030 | 117 (66%) | 1.12 (1.00, 1.25) | 0.045 |
| Not in Labor Force | 856 (25%) | 280 (33%) | 1.11 (0.99, 1.25) | 0.077 | 435 (51%) | 0.99 (0.92, 1.07) | 0.8 |
| **Food insecurity** |  |  |  |  |  |  |  |
| No | 2,625 (78%) | 852 (32%) | — |  | 1,406 (54%) | — |  |
| Yes | 750 (22%) | 415 (55%) | 1.55 (1.42, 1.69) | <0.001 | 548 (73%) | 1.28 (1.21, 1.35) | <0.001 |
| **Housing insecurity** |  |  |  |  |  |  |  |
| No | 2,900 (86%) | 1,010 (35%) | — |  | 1,606 (55%) | — |  |
| Yes | 452 (13%) | 241 (53%) | 1.41 (1.28, 1.56) | <0.001 | 328 (73%) | 1.24 (1.16, 1.32) | <0.001 |
| Don't know/ Not sure | 23 (0.7%) | 15 (65%) | 1.71 (1.24, 2.37) | 0.001 | 20 (87%) | 1.48 (1.28, 1.72) | <0.001 |
| **Parent/guardian of child under 18** |  |  |  |  |  |  |  |
| No | 2,589 (76%) | 895 (35%) | — |  | 1,415 (55%) | — |  |
| Yes | 797 (24%) | 375 (47%) | 1.12 (1.02, 1.23) | 0.023 | 550 (69%) | 1.11 (1.04, 1.18) | 0.002 |
| **Disability** |  |  |  |  |  |  |  |
| No | 2,689 (79%) | 970 (36%) | — |  | 1,533 (57%) | — |  |
| Yes | 697 (21%) | 300 (43%) | 1.23 (1.11, 1.35) | <0.001 | 430 (62%) | 1.11 (1.04, 1.18) | 0.003 |
| **Pregnancy** |  |  |  |  |  |  |  |
| No | 1,891 (98%) | 784 (41%) | — |  | 1,184 (63%) | — |  |
| Yes | 29 (1.5%) | 9 (31%) | 0.67 (0.39, 1.15) | 0.14 | 17 (59%) | 0.85 (0.62, 1.15) | 0.3 |
| **Underlying health conditions** |  |  |  |  |  |  |  |
| No | 1,301 (38%) | 550 (42%) | — |  | 800 (61%) | — |  |
| Yes | 2,089 (62%) | 722 (35%) | 0.93 (0.85, 1.02) | 0.11 | 1,167 (56%) | 1.00 (0.94, 1.06) | >0.9 |
| **Relationship status** |  |  |  |  |  |  |  |
| Married | 1,371 (41%) | 417 (30%) | — |  | 735 (54%) | — |  |
| Divorced/Widowed/Separated | 555 (16%) | 224 (40%) | 1.45 (1.28, 1.64) | <0.001 | 332 (60%) | 1.19 (1.09, 1.29) | <0.001 |
| Never married | 1,079 (32%) | 477 (44%) | 1.37 (1.23, 1.52) | <0.001 | 679 (63%) | 1.12 (1.04, 1.20) | 0.001 |
| Member of an unmarried couple | 375 (11%) | 147 (39%) | 1.18 (1.02, 1.38) | 0.029 | 212 (57%) | 0.98 (0.89, 1.09) | 0.7 |
| **Any healthcare barriers** |  |  |  |  |  |  |  |
| No | 2,579 (77%) | 792 (31%) | — |  | 1,355 (53%) | — |  |
| Yes | 759 (23%) | 449 (59%) | 1.75 (1.60, 1.90) | <0.001 | 573 (75%) | 1.33 (1.26, 1.41) | <0.001 |
| **No health insurance** |  |  |  |  |  |  |  |
| No | 3,231 (96%) | 1,151 (36%) | — |  | 1,825 (56%) | — |  |
| Yes | 143 (4.2%) | 109 (76%) | 1.92 (1.73, 2.13) | <0.001 | 129 (90%) | 1.48 (1.39, 1.58) | <0.001 |
| **Cost barriers** |  |  |  |  |  |  |  |
| No | 2,976 (89%) | 1,053 (35%) | — |  | 1,667 (56%) | — |  |
| Yes | 381 (11%) | 198 (52%) | 1.32 (1.19, 1.47) | <0.001 | 274 (72%) | 1.19 (1.11, 1.28) | <0.001 |
| **No primary care provider** |  |  |  |  |  |  |  |
| No | 2,910 (87%) | 944 (32%) | — |  | 1,582 (54%) | — |  |
| Yes | 441 (13%) | 302 (68%) | 1.93 (1.77, 2.11) | <0.001 | 354 (80%) | 1.38 (1.30, 1.46) | <0.001 |
| **Plans to get flu shot during Fall 2025/Winter 2026 season** |  |  |  |  |  |  |  |
| No, not planning to get it | 713 (21%) | 679 (95%) | — |  | 665 (93%) | — |  |
| Yes, planning to get it | 2,403 (71%) | 383 (16%) | 0.18 (0.16, 0.19) | <0.001 | 1,058 (44%) | 0.50 (0.47, 0.52) | <0.001 |
| Don't Know/Unsure | 274 (8.1%) | 210 (77%) | 0.79 (0.74, 0.85) | <0.001 | 244 (89%) | 0.94 (0.90, 0.99) | 0.018 |
| **Plans to get updated COVID-19 vaccine during Fall 2025/Winter 2026 season** |  |  |  |  |  |  |  |
| Definitely not get it | 498 (15%) | 396 (80%) | — |  | 491 (99%) | — |  |
| Definitely get it | 1,471 (43%) | 197 (13%) | 0.18 (0.15, 0.20) | <0.001 | 353 (24%) | 0.25 (0.23, 0.28) | <0.001 |
| Probably get it | 774 (23%) | 281 (36%) | 0.45 (0.41, 0.50) | <0.001 | 511 (66%) | 0.67 (0.63, 0.70) | <0.001 |
| Probably not get it | 647 (19%) | 398 (62%) | 0.75 (0.70, 0.81) | <0.001 | 612 (95%) | 0.95 (0.92, 0.97) | <0.001 |
| **Had flu vaccine in past 12 months** |  |  |  |  |  |  |  |
| No | 1,204 (36%) | 1,010 (84%) | — |  | 1,082 (90%) | — |  |
| Yes | 2,186 (64%) | 262 (12%) | 0.15 (0.13, 0.17) | <0.001 | 885 (40%) | 0.48 (0.45, 0.50) | <0.001 |
| Unsure | 0 (0%) | 0 (NA%) |  |  | 0 (NA%) |  |  |
| **Had COVID-19 vaccine in past 12 months** |  |  |  |  |  |  |  |
| No | 1,789 (53%) | 1,078 (60%) | — |  | 1,587 (89%) | — |  |
| Yes | 1,548 (46%) | 180 (12%) | 0.21 (0.18, 0.24) | <0.001 | 358 (23%) | 0.27 (0.25, 0.30) | <0.001 |
| Unsure | 53 (1.6%) | 14 (26%) | 0.44 (0.28, 0.70) | <0.001 | 22 (42%) | 0.47 (0.34, 0.65) | <0.001 |
| **Vaccine hesitancy** |  |  |  |  |  |  |  |
| Low hesitancy | 2,848 (84%) | 858 (30%) | — |  | 1,460 (51%) | — |  |
| Elevated hesitancy | 536 (16%) | 413 (77%) | 2.44 (2.27, 2.63) | <0.001 | 503 (94%) | 1.77 (1.69, 1.85) | <0.001 |
| **Confidence in the safety of the flu vaccine** |  |  |  |  |  |  |  |
| Not at all confident | 323 (9.5%) | 286 (89%) | — |  | 306 (95%) | — |  |
| Very confident | 2,181 (64%) | 494 (23%) | 0.27 (0.25, 0.30) | <0.001 | 974 (45%) | 0.49 (0.47, 0.52) | <0.001 |
| Somewhat confident | 793 (23%) | 417 (53%) | 0.60 (0.56, 0.65) | <0.001 | 605 (76%) | 0.82 (0.78, 0.86) | <0.001 |
| Don't know/ Not sure | 89 (2.6%) | 75 (84%) | 0.93 (0.84, 1.04) | 0.2 | 78 (88%) | 0.92 (0.84, 1.00) | 0.054 |
| **Confidence in the safety of the COVID-19 vaccine** |  |  |  |  |  |  |  |
| Not at all confident | 485 (14%) | 375 (77%) | — |  | 470 (97%) | — |  |
| Very confident | 1,961 (58%) | 424 (22%) | 0.29 (0.26, 0.32) | <0.001 | 770 (39%) | 0.42 (0.39, 0.44) | <0.001 |
| Somewhat confident | 825 (24%) | 396 (48%) | 0.60 (0.55, 0.66) | <0.001 | 618 (75%) | 0.76 (0.73, 0.79) | <0.001 |
| Don't know/ Not sure | 115 (3.4%) | 77 (67%) | 0.83 (0.72, 0.96) | 0.010 | 105 (91%) | 0.92 (0.86, 0.98) | 0.012 |
| **Confidence in federal agencies (CDC/FDA) to ensure vaccine safety and efficacy** |  |  |  |  |  |  |  |
| None at all | 515 (15%) | 232 (45%) | — |  | 319 (62%) | — |  |
| A lot | 793 (23%) | 243 (31%) | 0.66 (0.57, 0.75) | <0.001 | 415 (52%) | 0.82 (0.75, 0.90) | <0.001 |
| Some | 1,309 (39%) | 490 (37%) | 0.79 (0.71, 0.89) | <0.001 | 789 (60%) | 0.94 (0.86, 1.01) | 0.11 |
| A little | 771 (23%) | 307 (40%) | 0.86 (0.76, 0.97) | 0.017 | 442 (57%) | 0.90 (0.83, 0.99) | 0.028 |
| **Confidence in the safety and efficacy of mRNA vaccines** |  |  |  |  |  |  |  |
| None at all | 334 (9.9%) | 268 (80%) | — |  | 329 (99%) | — |  |
| A lot | 2,000 (59%) | 421 (21%) | 0.26 (0.24, 0.29) | <0.001 | 773 (39%) | 0.40 (0.37, 0.42) | <0.001 |
| Some | 793 (23%) | 400 (50%) | 0.59 (0.54, 0.64) | <0.001 | 614 (77%) | 0.75 (0.72, 0.79) | <0.001 |
| A little | 259 (7.6%) | 183 (71%) | 0.82 (0.74, 0.90) | <0.001 | 247 (95%) | 0.92 (0.88, 0.96) | <0.001 |
| **Trust in healthcare provider for vaccine information** |  |  |  |  |  |  |  |
| A lot | 2,412 (71%) | 664 (28%) | — |  | 1,178 (49%) | — |  |
| Some | 788 (23%) | 450 (57%) | 1.92 (1.76, 2.10) | <0.001 | 611 (78%) | 1.50 (1.42, 1.58) | <0.001 |
| A little | 136 (4.0%) | 109 (80%) | 2.61 (2.34, 2.92) | <0.001 | 127 (93%) | 1.76 (1.65, 1.88) | <0.001 |
| None | 52 (1.5%) | 49 (94%) | 3.29 (2.95, 3.68) | <0.001 | 49 (94%) | 1.87 (1.71, 2.05) | <0.001 |
| **Trust in local health dept for vaccine information** |  |  |  |  |  |  |  |
| A lot | 1,513 (45%) | 378 (25%) | — |  | 653 (43%) | — |  |
| Some | 1,385 (41%) | 566 (41%) | 1.56 (1.40, 1.73) | <0.001 | 900 (65%) | 1.45 (1.36, 1.55) | <0.001 |
| A little | 348 (10%) | 222 (64%) | 2.37 (2.10, 2.66) | <0.001 | 287 (82%) | 1.80 (1.67, 1.94) | <0.001 |
| None | 142 (4.2%) | 106 (75%) | 2.82 (2.46, 3.23) | <0.001 | 125 (88%) | 1.96 (1.79, 2.14) | <0.001 |
| **Trust in Centers for Disease Control (CDC) for vaccine information** |  |  |  |  |  |  |  |
| A lot | 965 (28%) | 313 (32%) | — |  | 531 (55%) | — |  |
| Some | 1,270 (37%) | 495 (39%) | 1.18 (1.06, 1.32) | 0.004 | 786 (62%) | 1.11 (1.04, 1.19) | 0.002 |
| A little | 739 (22%) | 266 (36%) | 1.11 (0.98, 1.27) | 0.11 | 391 (53%) | 0.97 (0.89, 1.06) | 0.5 |
| None | 414 (12%) | 198 (48%) | 1.56 (1.36, 1.78) | <0.001 | 257 (62%) | 1.18 (1.07, 1.29) | <0.001 |
| **Trust in pharmaceutical companies for vaccine information** |  |  |  |  |  |  |  |
| A lot | 239 (7.1%) | 76 (32%) | — |  | 105 (44%) | — |  |
| Some | 1,258 (37%) | 369 (29%) | 0.91 (0.75, 1.11) | 0.4 | 644 (51%) | 1.16 (1.00, 1.34) | 0.053 |
| A little | 1,171 (35%) | 442 (38%) | 1.13 (0.93, 1.38) | 0.2 | 700 (60%) | 1.32 (1.14, 1.53) | <0.001 |
| None | 719 (21%) | 385 (54%) | 1.62 (1.34, 1.97) | <0.001 | 515 (72%) | 1.60 (1.38, 1.84) | <0.001 |
| ^1^aRR = adjusted risk ratio. Adjusted for age and sex. | | | | | | | |
| Abbreviations: CI = Confidence Interval, IRR = Incidence Rate Ratio | | | | | | | |

#### **Supplementary Table S2. Influenza vaccination trajectories across prior vaccination, recent vaccination, intention, and 2025-26 behavior, CHASING COVID Cohort (N=3,346)**

| Previous flu vaccination (2022- 2023) | Recent flu vaccination (2024-25) | Intention for 2025-26 flu vaccination | 2025-26 flu vaccination | n (%) |
| --- | --- | --- | --- | --- |
| Vaccinated | Vaccinated | Plans to vaccinate | Vaccinated | 1645 (49.2%) |
| Not vaccinated | Not vaccinated | Does not plan to vaccinate | Not vaccinated | 608 (18.2%) |
| Not vaccinated | Vaccinated | Plans to vaccinate | Vaccinated | 223 (6.7%) |
| Vaccinated | Vaccinated | Plans to vaccinate | Not vaccinated | 143 (4.3%) |
| Not vaccinated | Not vaccinated | Unsure | Not vaccinated | 136 (4.1%) |
| Not vaccinated | Not vaccinated | Plans to vaccinate | Not vaccinated | 105 (3.1%) |
| Vaccinated | Not vaccinated | Plans to vaccinate | Vaccinated | 78 (2.3%) |
| Not vaccinated | Vaccinated | Plans to vaccinate | Not vaccinated | 71 (2.1%) |
| Not vaccinated | Not vaccinated | Plans to vaccinate | Vaccinated | 56 (1.7%) |
| Vaccinated | Not vaccinated | Plans to vaccinate | Not vaccinated | 54 (1.6%) |
| Vaccinated | Not vaccinated | Does not plan to vaccinate | Not vaccinated | 46 (1.4%) |
| Vaccinated | Not vaccinated | Unsure | Not vaccinated | 46 (1.4%) |
| Not vaccinated | Not vaccinated | Does not plan to vaccinate | Vaccinated | 22 (0.7%) |
| Vaccinated | Vaccinated | Unsure | Vaccinated | 21 (0.6%) |
| Not vaccinated | Not vaccinated | Unsure | Vaccinated | 16 (0.5%) |
| Not vaccinated | Vaccinated | Unsure | Vaccinated | 15 (0.4%) |
| Not vaccinated | Vaccinated | Unsure | Not vaccinated | 14 (0.4%) |
| Vaccinated | Vaccinated | Unsure | Not vaccinated | 12 (0.4%) |
| Vaccinated | Not vaccinated | Unsure | Vaccinated | 11 (0.3%) |
| Not vaccinated | Vaccinated | Does not plan to vaccinate | Not vaccinated | 7 (0.2%) |
| Vaccinated | Not vaccinated | Does not plan to vaccinate | Vaccinated | 7 (0.2%) |
| Vaccinated | Vaccinated | Does not plan to vaccinate | Not vaccinated | 7 (0.2%) |
| Vaccinated | Vaccinated | Does not plan to vaccinate | Vaccinated | 2 (0.1%) |
| Not vaccinated | Vaccinated | Does not plan to vaccinate | Vaccinated | 1 (0%) |

#### **Supplementary Table S3. COVID-19 vaccination trajectories across prior vaccination, recent vaccination, intention, and 2025-26 behavior, CHASING COVID Cohort (N=3,390)**

| Previous COVID-19 vaccination (2022- 2023) | Recent COVID-19 vaccination (2024-25) | Intention for 2025-26 COVID-19 vaccination | 2025-26 COVID-19 vaccination | n (%) |
| --- | --- | --- | --- | --- |
| Not vaccinated | Not vaccinated | Does not plan to vaccinate | Not vaccinated | 907 (26.8%) |
| Vaccinated | Vaccinated | Definitely plans to vaccinate | Vaccinated | 891 (26.3%) |
| Not vaccinated | Not vaccinated | Probably plans to vaccinate | Not vaccinated | 222 (6.5%) |
| Vaccinated | Not vaccinated | Probably plans to vaccinate | Not vaccinated | 183 (5.4%) |
| Vaccinated | Not vaccinated | Does not plan to vaccinate | Not vaccinated | 156 (4.6%) |
| Vaccinated | Vaccinated | Definitely plans to vaccinate | Not vaccinated | 148 (4.4%) |
| Vaccinated | Vaccinated | Probably plans to vaccinate | Vaccinated | 145 (4.3%) |
| Not vaccinated | Vaccinated | Definitely plans to vaccinate | Vaccinated | 102 (3%) |
| Vaccinated | Not vaccinated | Definitely plans to vaccinate | Vaccinated | 86 (2.5%) |
| Not vaccinated | Not vaccinated | Definitely plans to vaccinate | Not vaccinated | 72 (2.1%) |
| Vaccinated | Not vaccinated | Definitely plans to vaccinate | Not vaccinated | 69 (2%) |
| Vaccinated | Vaccinated | Probably plans to vaccinate | Not vaccinated | 67 (2%) |
| Not vaccinated | Vaccinated | Definitely plans to vaccinate | Not vaccinated | 64 (1.9%) |
| Vaccinated | Not vaccinated | Probably plans to vaccinate | Vaccinated | 54 (1.6%) |
| Not vaccinated | Not vaccinated | Definitely plans to vaccinate | Vaccinated | 39 (1.2%) |
| Not vaccinated | Vaccinated | Probably plans to vaccinate | Not vaccinated | 39 (1.2%) |
| Not vaccinated | Vaccinated | Probably plans to vaccinate | Vaccinated | 37 (1.1%) |
| Not vaccinated | Not vaccinated | Probably plans to vaccinate | Vaccinated | 27 (0.8%) |
| Not vaccinated | Vaccinated | Does not plan to vaccinate | Not vaccinated | 23 (0.7%) |
| Vaccinated | Vaccinated | Does not plan to vaccinate | Not vaccinated | 17 (0.5%) |
| Vaccinated | Not vaccinated | Does not plan to vaccinate | Vaccinated | 14 (0.4%) |
| Not vaccinated | Not vaccinated | Does not plan to vaccinate | Vaccinated | 13 (0.4%) |
| Vaccinated | Vaccinated | Does not plan to vaccinate | Vaccinated | 9 (0.3%) |
| Not vaccinated | Vaccinated | Does not plan to vaccinate | Vaccinated | 6 (0.2%) |

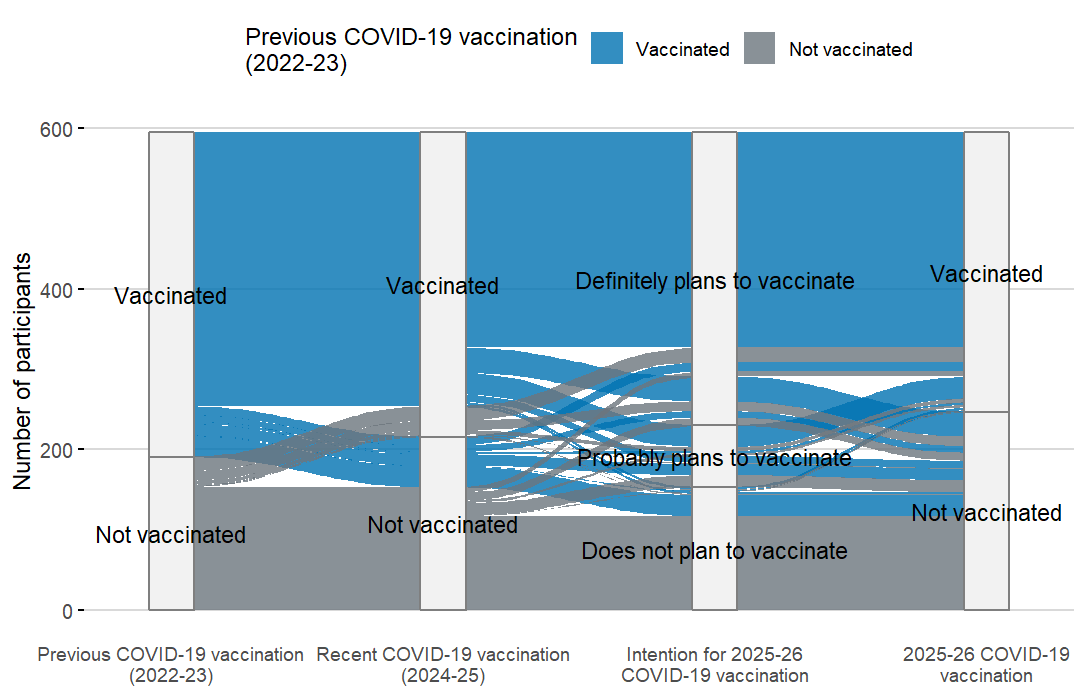
**Supplementary Figure S2.** **Participant trajectories across vaccination history, intention, and Fall 2025-Winter 2026 vaccination among adults aged ≥65 years, CHASING COVID Cohort (N=595)**

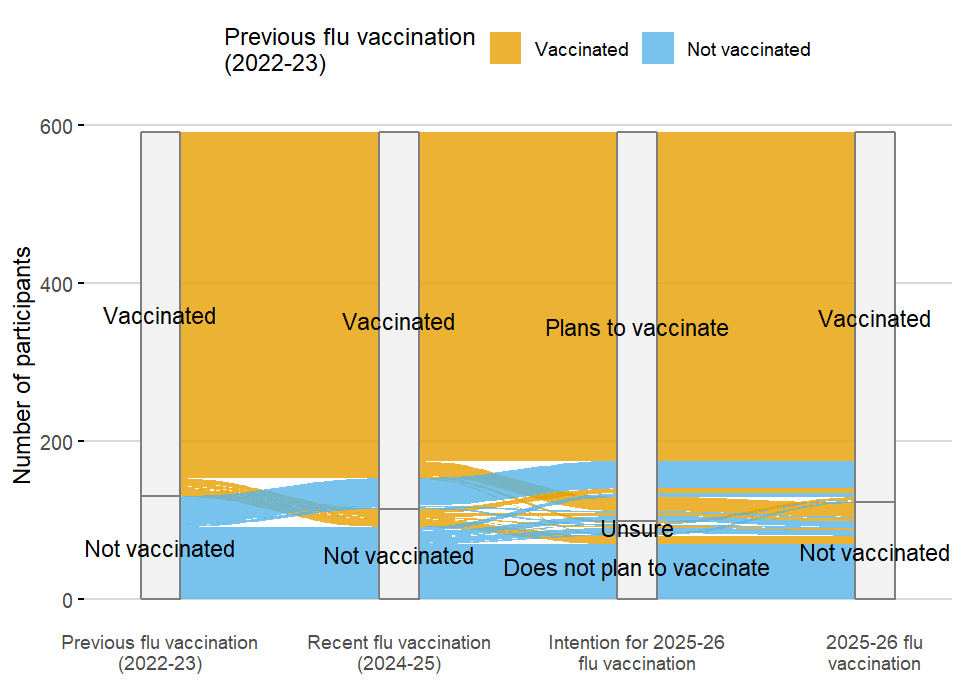

**Supplementary Table S4. Influenza vaccination trajectories across prior vaccination, recent vaccination, intention, and 2025-26 behavior among adults aged ≥65 years, CHASING COVID Cohort (N=592)**

| Previous flu vaccination (2022- 2023) | Recent flu vaccination (2024-25) | Intention for 2025-26 flu vaccination | 2025-26 flu vaccination | n (%) |
| --- | --- | --- | --- | --- |
| Vaccinated | Vaccinated | Plans to vaccinate | Vaccinated | 417 (70.4%) |
| Not vaccinated | Not vaccinated | Does not plan to vaccinate | Not vaccinated | 69 (11.7%) |
| Not vaccinated | Vaccinated | Plans to vaccinate | Vaccinated | 35 (5.9%) |
| Vaccinated | Vaccinated | Plans to vaccinate | Not vaccinated | 18 (3.0%) |
| Vaccinated | Not vaccinated | Does not plan to vaccinate | Not vaccinated | 10 (1.7%) |
| Not vaccinated | Not vaccinated | Plans to vaccinate | Not vaccinated | 7 (1.2%) |
| Not vaccinated | Not vaccinated | Unsure | Not vaccinated | 7 (1.2%) |
| Vaccinated | Not vaccinated | Plans to vaccinate | Vaccinated | 6 (1.0%) |
| Not vaccinated | Not vaccinated | Plans to vaccinate | Vaccinated | 5 (0.8%) |
| Vaccinated | Not vaccinated | Plans to vaccinate | Not vaccinated | 4 (0.7%) |
| Vaccinated | Vaccinated | Unsure | Vaccinated | 3 (0.5%) |
| Not vaccinated | Not vaccinated | Does not plan to vaccinate | Vaccinated | 2 (0.3%) |
| Not vaccinated | Vaccinated | Plans to vaccinate | Not vaccinated | 2 (0.3%) |
| Not vaccinated | Vaccinated | Unsure | Not vaccinated | 2 (0.3%) |
| Vaccinated | Not vaccinated | Unsure | Not vaccinated | 2 (0.3%) |
| Not vaccinated | Not vaccinated | Unsure | Vaccinated | 1 (0.2%) |
| Vaccinated | Not vaccinated | Does not plan to vaccinate | Vaccinated | 1 (0.2%) |
| Vaccinated | Vaccinated | Does not plan to vaccinate | Not vaccinated | 1 (0.2%) |

*Note: Three participants aged ≥65 years were excluded from the trajectory analysis because 2022-23 influenza vaccination data were unavailable.*

#### **Supplementary Table S5. COVID-19 vaccination trajectories across prior vaccination, recent vaccination, intention, and 2025-26 behavior among adults aged ≥65 years, CHASING COVID Cohort (N=595)**

| Previous COVID-19 vaccination (2022- 2023) | Recent COVID-19 vaccination (2024-25) | Intention for 2025-26 COVID-19 vaccination | 2025-26 COVID-19 vaccination | n (%) |
| --- | --- | --- | --- | --- |
| Vaccinated | Vaccinated | Definitely plans to vaccinate | Vaccinated | 268 (45%) |
| Not vaccinated | Not vaccinated | Does not plan to vaccinate | Not vaccinated | 117 (19.7%) |
| Vaccinated | Vaccinated | Definitely plans to vaccinate | Not vaccinated | 31 (5.2%) |
| Vaccinated | Vaccinated | Probably plans to vaccinate | Vaccinated | 27 (4.5%) |
| Vaccinated | Not vaccinated | Does not plan to vaccinate | Not vaccinated | 26 (4.4%) |
| Not vaccinated | Vaccinated | Definitely plans to vaccinate | Vaccinated | 18 (3.0%) |
| Not vaccinated | Not vaccinated | Probably plans to vaccinate | Not vaccinated | 15 (2.5%) |
| Vaccinated | Not vaccinated | Probably plans to vaccinate | Not vaccinated | 14 (2.4%) |
| Not vaccinated | Vaccinated | Definitely plans to vaccinate | Not vaccinated | 12 (2.0%) |
| Vaccinated | Not vaccinated | Definitely plans to vaccinate | Vaccinated | 11 (1.8%) |
| Not vaccinated | Not vaccinated | Definitely plans to vaccinate | Not vaccinated | 10 (1.7%) |
| Vaccinated | Vaccinated | Probably plans to vaccinate | Not vaccinated | 9 (1.5%) |
| Vaccinated | Not vaccinated | Definitely plans to vaccinate | Not vaccinated | 8 (1.3%) |
| Not vaccinated | Not vaccinated | Definitely plans to vaccinate | Vaccinated | 7 (1.2%) |
| Not vaccinated | Vaccinated | Probably plans to vaccinate | Vaccinated | 5 (0.8%) |
| Not vaccinated | Not vaccinated | Probably plans to vaccinate | Vaccinated | 3 (0.5%) |
| Vaccinated | Not vaccinated | Probably plans to vaccinate | Vaccinated | 3 (0.5%) |
| Vaccinated | Vaccinated | Does not plan to vaccinate | Not vaccinated | 3 (0.5%) |
| Vaccinated | Vaccinated | Does not plan to vaccinate | Vaccinated | 3 (0.5%) |
| Not vaccinated | Not vaccinated | Does not plan to vaccinate | Vaccinated | 1 (0.2%) |
| Not vaccinated | Vaccinated | Does not plan to vaccinate | Not vaccinated | 1 (0.2%) |
| Not vaccinated | Vaccinated | Does not plan to vaccinate | Vaccinated | 1 (0.2%) |
| Not vaccinated | Vaccinated | Probably plans to vaccinate | Not vaccinated | 1 (0.2%) |
| Vaccinated | Not vaccinated | Does not plan to vaccinate | Vaccinated | 1 (0.2%) |
